# A non-local diffusion magnetic resonance imaging tract density biomarker to stratify, predict, and interpret survival rates in human glioblastoma

**DOI:** 10.1101/2025.06.16.25329669

**Authors:** Joan Falcó-Roget, Gianpaolo Antonio Basile, Anna Janus, Sara Lillo, Letterio Salvatore Politi, Jan K. Argasinski, Alberto Cacciola

## Abstract

**Background:** Glioblastoma (GBM) is a lethal tumor, actively growing and invading neighboring neural tissue. GBMs appear functionally connected to distributed and spatially distant regions rather than representing an isolated and passive lesion disrupting the brain circuitry. Moreover, increasing evidence suggests that white matter serves as the morphological substrate for GBM to progress and migrate to distant areas in the human brain.

**Methods:** We hypothesized that the subset of white matter tracts intersecting the tumors depict the physical substrate for large-scale neuron-glioma interactions and would therefore inform prognosis. Using normative models, we design, analyze, interpret, and test a Lesion-Tract Density Index (L-TDI) marker that considers the distributed white matter pathways interacting with the tumor in two independent cohorts of *N* = 367 and *N* = 496 patients, respectively.

**Results:** First, we show that the average tract density within this white matter map robustly stratifies survival rates due to widespread white matter involvement. Second, we demonstrate why tract density-based markers offer critical and necessary insights into the morphology, location, and evolution of human GBM by proving how the proposed L-TDI implicitly considers tumor volume, white matter density, and location. We provide further evidence that the non-uniform distribution of GBMs and their differential prognosis emerge from white matter morphology. Third, we validate the L-TDI marker with multiple Cox survival models and analyze its contribution in relation to other covariates of interest (e.g., MGMT promoter methylation). Lastly, by using a simple logistic model, we predict patient death at 12 months with balanced accuracies of 0.68 and 0.65, and areas under the curve of 0.74 and 0.73 when training and testing in separate and independent cohorts.

**Conclusions:** Overall, we offer a concrete implementation of the emerging paradigm that views GBM not as a focal lesion, but as a network disease shaped by its complex interactions with distant brain regions.

## 1. Introduction

Recent discoveries in cancer neuroscience have revealed that gliomas, particularly glioblastomas (GBMs), exhibit dynamic interactions with neuronal circuits that go far beyond passive infiltration (Sadanand, 2019; Venkataramani et al., 2020). Unlike stroke or traumatic brain injury, which manifest as structural disconnections, gliomas appear to functionally integrate within existing brain networks (Venkataramani et al., 2022). They leverage synaptic plasticity to reshape local and long-range circuitry (Krishna et al., 2023; Taylor et al., 2023), evade detection, and recur in anatomically unpredictable locations, ultimately contributing to reduced overall survival rates (Ostrom et al., 2014; Ostrom, Francis and Barnholtz-Sloan, 2021). Increasing evidence suggests the existence of a positive feedback loop between glioma-induced neuronal activity (Buckingham et al., 2011; Campbell, Buckingham and Sontheimer, 2012) and neuron-glioma connectivity (Venkatesh et al., 2015; Venkatesh et al., 2019), which collectively promote tumor growth and invasion (Venkataramani et al., 2019). This active and adaptive behavior supports the view of gliomas as complex systems that interact with, and co-opt, the host brain’s own connectome. These features hinder the existence of two interdependent networks, the brain and neuron-glioma networks, both capable of functioning and evolving in relative isolation while influencing one another, thus giving rise to critical and seemingly random system behaviors (Gao et al., 2011; Herbet and Duffau, 2020).

In this conceptual framework, three central questions arise: 1) how does the whole-brain connectivity changes in the presence of GBM (Sun et al., 2025); 2) what are the properties of the neuron-glioma networks (Tetzlaff et al., 2025), and 3) to what extent does the tumor’s spatial embedding within the brain connectome, rather than its size or spatial location alone, determine its clinical impact, including survival? The answers to these questions remain largely unexplored *in-vivo* and the present study seeks to specifically address the latter.

We explore how these emerging insights can be translated into the neuroimaging domain. Unlike cancer neuroscience, which primarily relies on *in-vitro*, animal, or organoid models (Monje et al., 2020; Shi et al., 2022), neuroimaging may offer rapid clinical applicability due to its non-invasive nature. Functional magnetic resonance imaging studies have mostly focused on the connectivity between the tumor and the rest of the brain (Stoecklein et al., 2020; Daniel et al., 2020; Falcó-Roget et al., 2024), and its potential association with survival and cognitive deficits (Daniel et al., 2021; Sprugnoli et al., 2022). In contrast, global alterations in functional connectivity profiles have shown less promising clinical implications (Derks et al., 2017; Nenning et al., 2020; Silvestri et al., 2022; Park et al., 2023). Diffusion magnetic resonance imaging studies have primarily focused on structural alterations or impairments in whole-brain connectivity (Fekonja et al., 2022; Wei et al., 2022), which appear more robustly associated with survival and cognitive outcomes (Wei et al., 2022; Herbet, Duffau and Mandonnet, 2024). Yet, these attempts often failed to conceptualize the tumor itself as a component of a broader network (Tetzlaff et al., 2025), ultimately determining disease progression.

To address this gap, we move towards a connectomics perspective (Duffau and Filippi, 2024; Salvalaggio et al., 2024), modeling GBM as a node embedded in and interacting with a white matter scaffold, rather than a focal lesion. Building on recent work that used local tract density as a proxy for infiltration (Salvalaggio et al., 2023), we introduce a novel approach, the lesion-tract density index (L-TDI), which incorporates both the lesion’s morphology and its spatial embedding within normative white matter tractograms. Herein, we apply this framework to two large, independent GBM cohorts, aiming at providing an imaging biomarker for GBM stratification and survival prediction.

## 2. Methods

In this study, two independent and publicly available GBM cohorts were employed. A comprehensive overview of the methodological framework is shown in Fig. 1, while a more detailed description of the conceptual, theoretical, and computational steps performed is available in the Supplementary Material.

**Figure 1.**
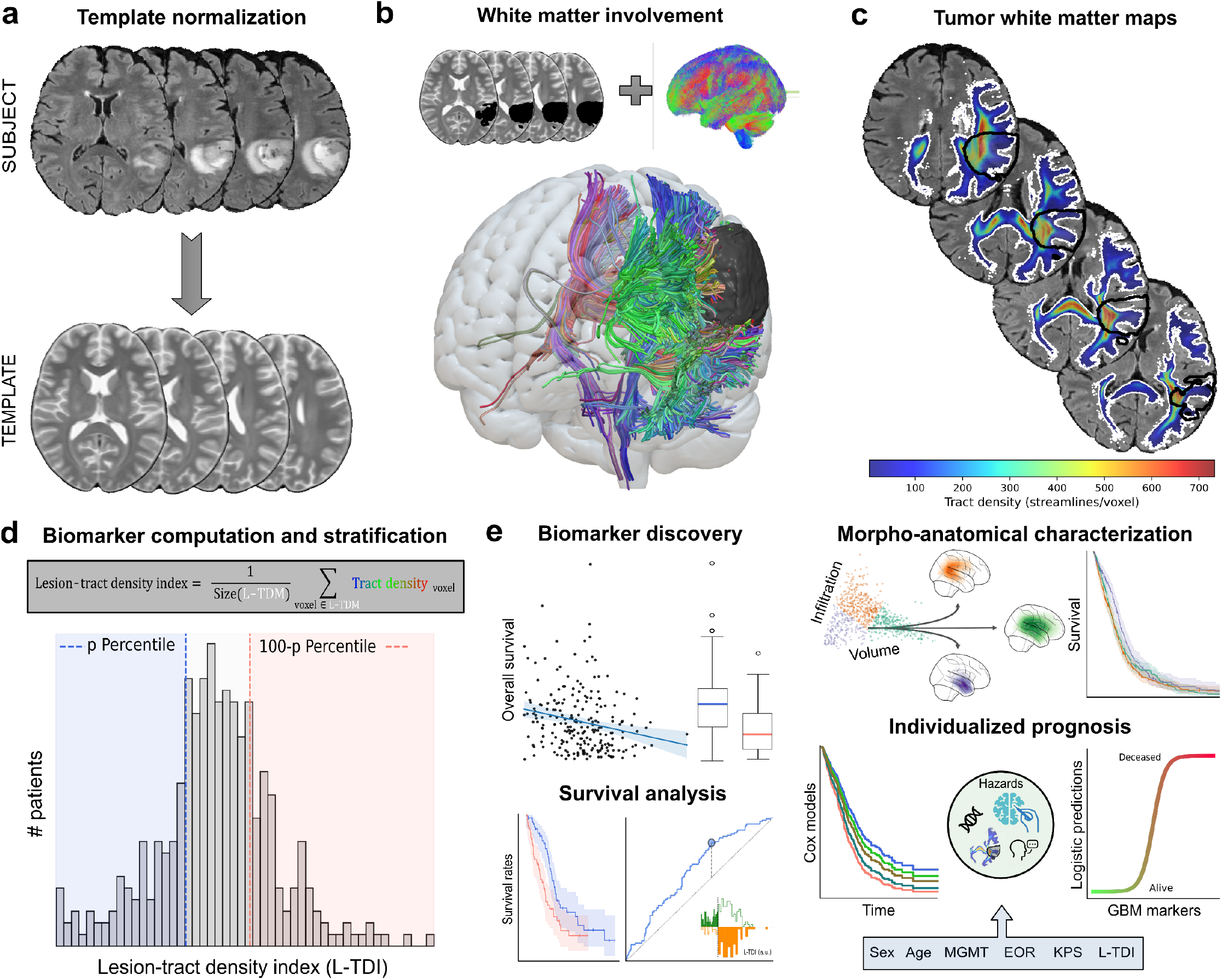
The lesion-tract density index (L-TDI): a survival marker in human GBM. **(a)** For every subject, the data was normalized to a common template. The T2-weighted image was chosen as the reference modality, and the GBM was inversely masked to discard distorted tissue during the optimization. **(b)** The normalized tumor mask (black volume) is used in combination with the whole brain normative tractogram to extract the set of streamlines that intersect the tumor in the same template space as in **(a). (c)** The lesion-tract density map (L-TDM,white contour) provides a unique white matter density map describing the tracts in **(b)** that interact with the GBM (black contour). **(d)** From the L-TDM, we can define the L-TDI marker by averaging the tract density within the L-TDM binary mask (white contour in **(c)**). For a given cohort, the sample can be stratified according to a given percentile. **(e)** Once a stratification threshold has been set, we conduct thorough tests to assess the discovery of the biomarker. Additional survival analyses on multiple stratification thresholds help determine whether the potential biomarker holds prognostic value. The L-TDI can also be used to characterize both morphological and anatomical landscapes in GBMs, which can be used to understand differences in the survival rates. Once a robust eMect has been identified, the L-TDI can be incorporated into standard risk analysis and predictive frameworks to improve patient care.

### 2.1. UCSF Cohort: The University of California San Francisco glioblastoma cohort

From the original 501 samples available in the University of California San Francisco dataset (Calabrese et al., 2022), we included the *N* = 367 patients with recorded survival data and confirmed grade 4 glioblastoma IDH-wildtype diagnosis as per the WHO 2021 classification (Louis et al., 2021). All the 6 follow-up entries from the original 501 were discarded. All the selected patients had available high-quality multimodal imaging data and tumor multi-tissue segmentations obtained from automated deep learning models and manually corrected if necessary (Calabrese et al., 2022).

Clinical data (Table 1) included biological sex, age at the time of the MRI scan, MGMT (O^6^-methylguanine-DNA methyltransferase) promoter status, MGMT index, and the extent of resection (EOR) based on the surgical and/or postoperative MRI reports. Overall survival (OS) was measured in days from the initial diagnosis to the date of death (status=1) or the last clinical follow-up (status=0). For this cohort, 39.24%of the patients were right-censored (i.e., lost to follow-up). Additionally, 41 patients did not undergo a full surgical procedure. While the prevalence of GBMs was slightly higher in males than females, we did not detect a significant sex-difference in prognosis. Instead, as expected, positive MGMT promoter status and gross total resection (GTR) as opposed to subtotal resection (STR) were associated with increased survival rates (Fig. S1).

**Table 1.** Clinical covariates of interest. Demographic information and absolute counts for each of the two analyzed cohorts. Age was measured in years [mean±SEM]. The average age in males was significantly lower after joining the cohorts (*t* = −2.1410, *p* = 0.0326; two-sided Welch’s t-test), but not in the UCSF (*t* = −1.2484, *p* = 0.2128; two-sided Welch’s t-test) nor the UPENN (*t* = −1.7679, *p* = 0.0779; two-sided Welch’s t-test) cohorts separately. The MGMT promoter categories were positive (P), negative (N), Indeterminate (I), and unavailable (Unv) methylation. The categories for the extent of resection were gross total resection (GTR), subtotal resection (STR), biopsy (bio; no major surgical intervention was performed), and unavailable (Unv). Karnofsky performance status (KPS) scores were only available for 75 patients from the UPENN cohort (see Fig. S2e).

| Cohort | Biological sex (M/F) | Age at MRI (M/F) | MGMT promoter status (P/N/I/Unv) | Extent of resection (GTR/STR/bio/Unv) | Total |
| --- | --- | --- | --- | --- | --- |
| UCSF | 218/149 | 61.15±0.82/62.74±0.98 | 246/104/5/12 | 214/112/41/0 | 367 |
| UPENN | 297/199 | 62.63±0.64/64.53±0.87 | 103/151/25/217 | 292/180/0/24 | 496 |
| Joint | 515/348 | 62.00±0.39/63.76±0.41 | 349/255/30/229 | 506/292/41/24 | 863 |

### 2.2. UPENN Cohort: The University of Pennsylvania glioblastoma cohort

From the original 671 samples available in the University of Pennsylvania dataset (Bakas et al., 2022), we included the *N* = 496 patients with recorded survival data and confirmed glioblastoma grade 4 IDH-wildtype diagnosis as per the WHO 2021 classification (Louis et al., 2021). All the 60 follow-up entries from the original 671 were discarded. All the selected patients had available high-quality multimodal imaging data and tumor multi-tissue segmentations obtained from automated deep learning models and manually corrected if necessary (Bakas et al., 2022).

Clinical data (Table 1) included biological sex, age at the time of the MRI scan, MGMT (O^6^-methylguanine-DNA methyltransferase) promoter status, MGMT index, and the extent of resection (EOR) representing excision of more or less than 90% of the tumor. Karnofsky performance status (KPS) scores were available for *N* = 53 patients. OS was measured in days from the first surgical intervention until the date of death (status=1) or the last clinical follow-up (status=0). For this cohort, 2.82%of the patients were right-censored (i.e., lost to follow-up). As per the UCSF cohort, the prevalence of GBMs was slightly higher in males than females, but we did not detect any significant differences in prognosis. Instead, as expected, positive MGMT promoter status, high KPS score, and a gross total resection (GTR), as opposed to subtotal resection (STR), were associated with increased survival rates (Fig. S2).

### 2.3. Image normalization to the MNI template

For both cohorts, the structural and multi-label lesion segmentations were already co-registered (i.e., aligned in subject space). We nonlinearly normalized all the necessary images to the Montreal Neurological Institute (MNI ICBM 2009b NLIN Asymmetric; hereafter referred to as “MNI template”) standard space (Elias et al., 2024). Before normalizing, for both cohorts, we binarized and inverted the multi-label lesion, ensuring that no pathological tissue was used during the registration process. Then, we nonlinearly mapped the T2 images to the T2 MNI template (Avants et al., 2008) and applied the transformation to the tumor masks. The normalized images were overlaid onto the MNI template to visually assess the alignment of anatomical landmarks (e.g., sulci and gyri). As a last step, the volume of the tumors was computed in cubic centimeters in MNI space (Fig. 1a).

### 2.4. Normative tractograms, tract density maps and indices

To evaluate the white matter pathways interacting with the tumor lesion, we sought to employ a publicly available normative structural tractogram constructed from 985 healthy subjects and 11,820,000 streamlines (Elias et al., 2024). To compute a patient-specific lesion-tract density map (L-TDM) and index (L-TDI), we selected the streamlines that intersected a given tumor mask (Fig. 1b-c) and obtained the corresponding L-TDM by counting the number of streamlines passing through every voxel (Calamante et al., 2010). The L-TDI was the average tract density within the L-TDM (Fig. 1d), thus encompassing the average white matter density of the whole brain-lesion normative circuit. For completeness, we also obtained the tract density map (TDM) and index (TDI) of every patient (Salvalaggio et al., 2023). For that, from the normative tractogram, we generated a voxel-wise density map, overlaid each tumor mask to create patient-specific TDMs, and computed the TDI as the average density within each tumor. In summary, the L-TDI modeled the GBM as a distributed white matter lesion, as opposed to the local (within tumor) density of potentially infiltrated white matter measured by the TDI.

### 2.5. Survival analysis and statistical procedures

We treated UCSF as the discovery and UPENN as the replication cohort to avoid dataset-specific bias. In the UCSF, we stratified patients into “Low” and “High” L-TDI groups using percentiles ranging from 20 to 50 (in steps of 5). Kaplan-Meier curves were compared using two-sided log-rank tests, with p-values False Discovery Rate (FDR)-corrected across several percentiles. We then assessed median OS for deceased patients using two-sided Mann-Whitney U-tests, also FDR-corrected. This confirmed robust, threshold-independent survival differences based on tract density-based stratification. We compared average L-TDI and TDI between patients who died or survived past specific timepoints (6–48 months) using two-sided Mann-Whitney U-tests, FDR-corrected. We also computed Pearson correlations (ρ) between (L-)TDIs and survival times within subgroups of patients who died before each threshold. Significance was assessed via two-sided permutation tests (*n* = 2500 resamples), with FDR correction applied across thresholds.

Lastly, we computed ROC curves using raw L-TDI values to classify survival at 12 and 24 months, reporting balanced accuracy via Youden’s J index (Youden, 1950). Additionally, we assessed the time-dependent discriminative power of tract density markers using cumulative and dynamic AUCs (Hung and Chiang, 2009). A repeated K-fold cross-validation (50 runs) estimated the censoring distribution, with time sampled from 6 to 36 months and K set to 2, 6, and 8. Dynamic AUCs were trained on (K–1) folds and tested on the held-out split. All the previous analyses were then repeated in the same way for the replicative cohort.

### 2.6. Dimensionality reduction and clustering of macroscopic features in GBMs

To understand the relationship between tract density indices and tumor volume, we performed linear principal component (PC) analyses with the TDI, L-TDI, and volume as features of interest. The 1^st^ and 2^nd^ PCs accounted for more than 99% of the variance, indicating that the L-TDI was linearly predictable from them. Then, we performed automatic *K*-means clustering of patients based on the TDI and tumor volume. We calculated the Kaplan-Meier curves of each of the 3 found groups and obtained the average overlap of every brain lobe delineated in the USCLobes brain atlas (Joshi et al., 2022) with the tumor masks. Similarly, to move beyond traditional lesion-symptom mappings, we computed the average overlap between the white matter tracts defined in the XTRACT atlas (Warrington et al., 2020) and the L-TDMs. The resulting overlaps were then sorted in descending order and compared between clusters using both Spearman correlation and Kendall’s τ coefficients.

### 2.7. Cox proportional hazard modeling

To evaluate the added clinical value of tract density-based markers, we fitted four Cox proportional hazard models per cohort:

1. a *naïve* model with classical covariates (sex, age, MGMT status, EOR),
2. an *L-TDI* model (naïve + L-TDI),
3. a *TDI* model (naïve + TDI),
4. a *TDI+Volume* model (naïve + TDI + lesion volume).

Models were compared using log-likelihood ratio tests. Hazard ratios were assessed via two-sided Wald tests with associated T-distributions, and 95% confidence intervals were reported.

To improve the estimates of the hazard rates, we also joined the survival data from the two cohorts, effectively doubling the sample size (see Supplementary Materials). However, since the survival was measured from days of diagnosis and from days of surgery in the UCSF and UPENN cohorts, respectively, we accounted for measurement-related site differences in survival rates. For each patient, we obtained the site-corrected OS,

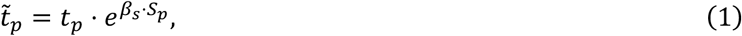

where *β*_*S*_ is estimated from a Cox model incorporating a variable *S*_*p*_ = {0,1} for the UCSF and UPENN cohorts, respectively. After applying the transformation in Eq. (1) to all the survival times, we fitted a second Cox multivariate model to evaluate the hazard ratios associated with each covariate. When comparing survival rates across different strata, the time must be measured using the same reference units. Crucially, changing the measurement units introduces a proportional shift in the underlying hazard rates (e.g., 365 days = 12 months). Since the covariate *S*_*p*_ complies with the assumption of proportional hazards (see Fig. S3), the transformation in Eq. (1) and the procedure described here can be reliably used to estimate the conversion factor between “days from diagnosis” and “days from surgery”. See the Supplementary Materials for details on the effects that this transformation causes on the survival and hazard rates.

Lastly, we computed Harrell’s concordance index (*C*^*H*^) to evaluate the discriminative power of a given feature (Harrell, Lee and Mark, 1996). For each feature (e.g., MGMT promoter status), we fitted a simple Cox model and computed the associated *C*^*H*^ score. The significance was assessed through non-parametric permutation tests (*n* = 1000)resamples). All the procedures described here were carried out after discarding the samples with missing data.

### 2.8. Logistic regression modeling of survival and death predictions

We fitted logistic regression models to assess the contribution of each feature to survival at a given time point (1 died, 0 survived). Mirroring the Cox analyses, we tested four models to evaluate the tract density-based markers, recording each covariate’s significance, confidence interval, and model log-likelihood. The different models were compared using log-likelihood ratio tests. We fitted models to each cohort independently and after merging the survival data.

To predict whether a patient would die before a specified time point, we trained a logistic classifier using repeated stratified cross-validation. A grid search varied the number of splits (2–12) and repeats (5–25), with performance evaluated using both AUC and balanced accuracy (bACC). We selected the parameter set that minimized the average of the two scores to balance class separability and true positive rates. The final model was retrained on the full training cohort and tested on the independent cohort. This process was repeated for 6, 12, and 18-month predictions, alternating training and testing cohorts. Survival times were pre-corrected for site effects as described earlier. All the procedures described here were carried out after discarding the samples with missing data.

## 3. Results

### 3.1. The Lesion-tract density index (L-TDI) is robustly associated with survival in GBM

We investigated whether the set of streamlines intersecting the tumor was associated with overall survival (OS). Prior work postulated that the normative white matter tract density index (TDI) within the tumor significantly modulated OS when stratifying the population into two groups based on the 1^st^ and 3^rd^ quartiles of the density distribution (Salvalaggio et al., 2023). Building on this approach, we instead computed the average normative white matter density of the entire lesion-intersecting tractogram rather than limiting the analysis to the tumor mask alone (see Methods). In both the discovery (i.e., UCSF) and replicative (i.e., UPENN) cohorts, higher values of lesion-tract density index (L-TDI) were significantly associated with worse prognosis (Fig. 2a-b). To measure the robustness of this marker, we also evaluated multiple stratification thresholds (Fig. 2c). In the UCSF cohort, Kaplan–Meier curves consistently showed significant survival differences between low and high L-TDI groups across all thresholds. Similarly, in the UPENN cohort, patients with high L-TDI exhibited significantly worse survival in 5 out of 7 stratifications, with the remaining two thresholds approaching statistical significance. As a result, median survival times were also significantly reduced in patients with high L-TDIs in both the discovery (Low L-TDI 9.58 [4.15, 13.74] and High L-TDI: 17.88 [11.14, 21.06] months after diagnosis; stratified at 25/75-th percentiles) and replicative (Low L-TDI 15.74 [4.96, 19.00] and High L-TDI: 19.89 [11.21, 24.39] months after surgery; stratified at 25/75-th percentiles) cohorts (Fig. 2d-e; Fig. S4b-5b).

**Figure 2.**
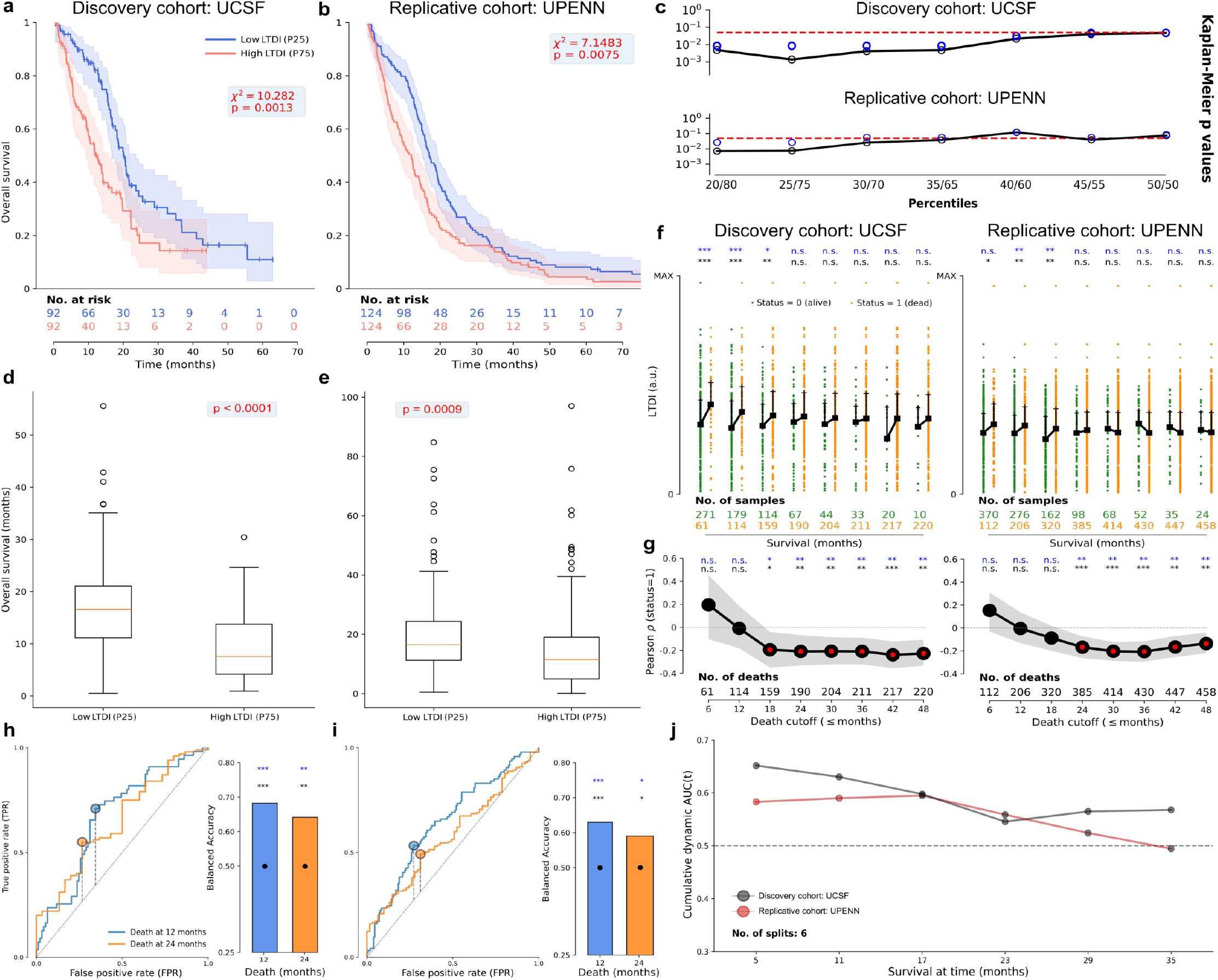
Lesion-tract density index (L-TDI) and overall survival in GBM patients. **(a-b)** Kaplan-Meier curves for the two strata at the *p* = 25-th percentile of the L-TDI distribution for **(a)** the UCSF and **(b)** the UPENN cohorts. The group with low L-TDI (blue) is significantly associated with longer survival times than the high L-TDI (red) group (*χ*^2^ = 10.2820, *p* = 0.0012, two-sided log-rank test). The shaded areas correspond to the 95% confidence interval. Small vertical dashes indicate right-censored entries. **(c)** Natural logarithm of the two-sided log-rank *p*-value (black solid lines) as a function of the stratification thresholds in the UCSF (top) and UPENN (bottom) cohorts. The blue circles are the FDR significance values. Values below the significance level (red dashed horizontal lines; *α* = 0.05) correspond to strata with significant differences in survival rates. The Kaplan-Meier curves for the lowest *p*-value in the discovery cohort (i.e., *p* = 25/75-th percentiles) are plotted in **(a-b). (d-e)** In patients known to have died, the median survival times for the two cohorts and strata shown in **(a-b)** were significantly higher in the low L-TDI group (*p* < 0.001, two-sided Mann-Whitney U-tests); both in the UCSF **(d)** and UPENN **(e)** cohorts. **(f)** L-TDI distributions of dead (orange) and alive (green) patients across multiple survival thresholds in the UCSF **(left)** and UPENN **(right)** cohorts. Black squares and lines show the median and 3^rd^ quartiles, respectively. In black text, the *p*-values of each comparison (‘***’ *p* ≤ 0.001, ‘**’ *p* ≤ 0.01, ‘*’ *p* ≤ 0.05, ‘n.s.’ *p* ≥ 0.05, two-sided Mann-Whitney U-tests). In blue text the FDR corrected significance values. **(g)** Pearson correlation between survival times and L-TDIs in patients known to have died in the UCSF **(left)** and UPENN **(right)** cohorts (see Methods). The correlation stabilized around *ρ* = −0.2 after 18 months. **(h-i)** Receiver operating characteristic (ROC) curves and balanced accuracies (bACC) obtained from the same strata as **(a-b)** in the **(h)** UCSF and **(i)** UPENN cohorts. Separability was assessed at 12 (light blue) and 24 (orange) months. The corresponding bACCs were obtained using Youden’s J index (gray dashed vertical lines and colored circles). **(j)** For both discovery (gray solid line) and replicative (dark red line) cohorts, the time-dependent AUC was stable up to 18 months, after which it started to decay. The months in which the dynamic AUC was calculated were automatically selected depending on the cross-validation scheme (see Methods).

To move beyond the use of arbitrary thresholds, we assessed whether patients who had died by specific time points had lower L-TDIs (Fig. 2f). Noteworthy, alive patients at 6, 12, and 18 months consistently exhibited lower indices. In fact, the Pearson correlation between the OS and L-TDI progressively decreased and settled around *ρ* = −0.2 when considering patients surviving 1.5-2 years from the date of diagnosis or surgery (Fig. 2g). These two findings revealed a natural survival cutoff situated close to the median survival times where the L-TDI showed stable behavior. Specifically, patients with low-L-TDI had increased probabilities of living more than 18 months compared to patients whose GBMs resulted in higher indices (Fig. S4d-S5d). To test this cutoff in a more quantitative way, we measured the separability of the two strata defined from the 1^st^ and 3^rd^ quartiles (Fig. 2h-i). Similar results were observed when using an alternative 50/50 stratification, though the balanced accuracy for predicting survival at 12 and 24 months showed a slight decline (Fig. S6d-h; Fig. S7d-h). Lastly, the cumulative dynamic area under the curve (see Methods), was stable until the 18-month point, after which it started to decrease in both the UCSF and UPENN cohorts. (Fig. 2j).

In summary, the proposed L-TDI consistently stratifies survival times in patients with GBM. To better contextualize this finding, we also computed the corresponding TDI and conducted parallel analyses in both cohorts (Fig. S4-7). Most notably, higher TDIs were associated with reduced survival times only in the replicative cohort (i.e., UPENN), suggesting its potential, despite limited, prognostic role. However, our results did not favor this local tumor description over the distributed model provided by the L-TDI. Indeed, in the discovery cohort, TDI showed no significant association with survival across multiple stratification thresholds (Fig. S6a–c), failing to distinguish survival outcomes at 6, 12, or 18 months (Fig. S4c; Fig. S6d-i). Nonetheless, we do not consider TDI and L-TDI as competing metrics, but rather as complementary descriptors of GBM white matter involvement. We therefore sought to further investigate potential redundancies and synergies between the two density-based indices.

### 3.2. The L-TDI provides a morpho-anatomical model of patient survival

A key concern when introducing new biomarkers is determining whether they offer genuinely novel insights or merely reflect existing information, particularly in clinical settings, where interpretability is essential for informing therapies and targets. To this end, we examined the relationship between tumor volume, the proposed L-TDI, and the TDI. While both tract density-based markers were moderately correlated with each other, their association appeared limited to lower index values, indicating that they capture similar information only in a limited subset of cases (Fig. 3a-b). Additionally, the L-TDI showed a strong positive correlation with tumor volume (UCSF: *ρ* = 0.85, *p* < 0.001; UPENN: *ρ* = 0.81, *p* < 0.001; two-sided exact tests), whereas TDI was only weakly related to the tumor volume (UCSF: *ρ* = 0.06, *p* < 0.155; UPENN: *ρ* = 0.10, *p* < 0.048; two-sided exact tests). Since the same normative tractogram was used for both cohorts, we pooled the data into a unified set of markers. Interestingly, while tumor volume and TDI differed slightly between cohorts, the L-TDI exhibited highly consistent distributions, hinting at an increased generalizability across independent samples (Fig. S8).

**Figure 3.**
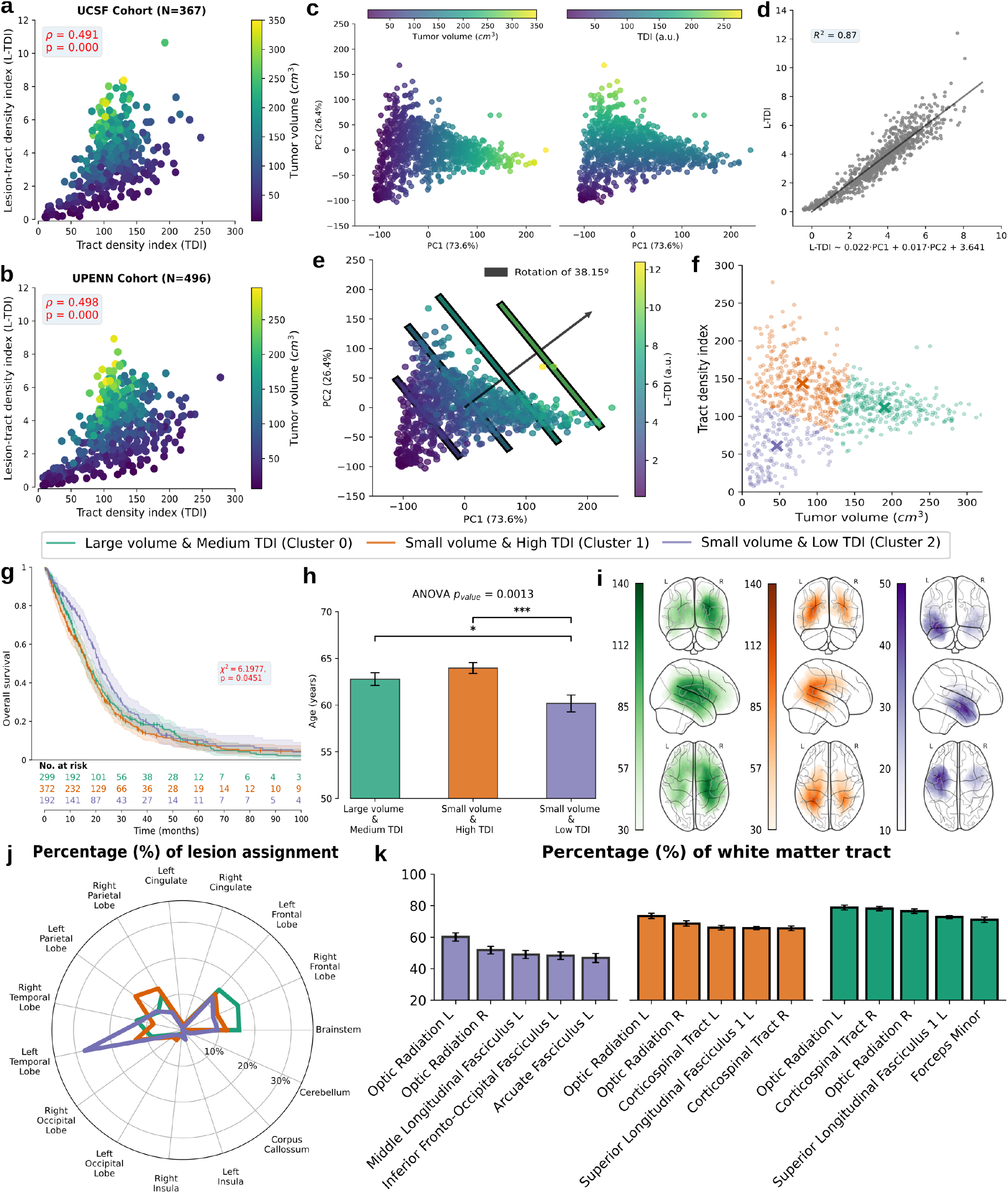
Tract density markers, morphology, and anatomy of GBM. **(a-b)** Scatter plots of the tract-density markers and the corresponding tumor volumes in **(a)** UCSF and **(b)** UPENN cohorts. Tumor volume increased somewhat linearly with the L-TDI but not with the TDI. The red text shows the Pearson correlation and the corresponding two-sided exact tests. **(c)** Principal component (PC) analysis of the 3 features in (a-b). More than 99.9% of the explained variance can be attributed to the 1^st^ and 2^nd^ PCs. The 1^st^ and 2^nd^ PCs largely account for the variance in the tumor volumes (left) and TDIs (right). **(d)** Observed and predicted L-TDI values obtained from the linear combination of the first two PCs. The *R*^2^ depicts the determination coefficient of the linear fit. **(e)** Scatter plot of the 1^st^ and 2^nd^ PCs together with the corresponding L-TDIs. The diagonal lines depict areas of constant L-TDI, and the black arrow indicates the direction along which the L-TDI increases. Because of the orthogonality of the first two PCs in **(d-e)**, the L-TDI contains unique and non-redundant information of both tumor volume and indirect white matter infiltration. **(f)** Automatic K-means clustering from the tumor volumes and TDIs reveal 3 distinct groups. **(g)** Kaplan-Meier curves of each group in **(f)** revealed decreased hazard rates for small tumors with reduced infiltration (purple) when compared with small tumors with increased infiltration as well as large tumors (*χ*^2^ = 6.1977, *p* = 0.0451, two-sided log-rank test). Survival rates were adjusted for site effects (see Methods). **(h)** Average [mean ± SEM] age in years for each cluster in **(f)**. At least one cluster had a significantly difference age (*F* = 6.6746, *p* = 0.0013; one-way ANOVA). Follow-up analyses revealed that small tumors with low TDIs occurred in significantly younger patients (post hoc Tukey’s HSD). **(i)** Maximum intensity projection of the lesion group maps for the 3 distinct clusters in **(f)**. The colorbars show the number of subjects with a GBM in each point of the brain. **(j)** Average percentage of the lesion that overlaps with each brain lobe defined in the USCLobes atlas for each cluster in **(f)**. Percentages were calculated by averaging the overlap in each patient belonging to every distinct cluster. **(k)** Average percentage of overlap between the major white matter tracts defined in XTRACT atlas and the L-TDM of every subject belonging to each cluster in **(f)**. The white matter tracts appear sorted in decreasing order and only the first 5 are shown (Fig. S11c-e).

To disentangle the contributions of these three features, we performed linear dimensionality reduction and observed that more than 99% of the variance was captured by the first two principal components. Further inspections revealed that the 1^st^ and 2^nd^ PCs corresponded to the volume and TDI, respectively (Fig. 3c), confirming the linear independence observed before. Crucially, this implied that the L-TDI should be linearly predictable from the linear combination of the first two PCs (Fig. 3d; *R*^2^ = 0.87). In other words, the L-TDI contained both information about the tumor volume and the normative local average white matter density, linearly and monotonously increasing with both size and density of white matter (Fig. 3e).

It is worth noting that the computation of the L-TDI does not explicitly consider the size of the tumor, but, given that larger tumors intersect more normative tracts, higher L-TDI values were naturally expected for larger lesions. In contrast, the TDI presented a more complex, non-linear relationship with the morphology of the tumor. However, linear independence need not imply the absence of any relationship. Indeed, when applying unsupervised clustering to the two-dimensional space defined by tumor volume and TDI, three distinct GBM subtypes emerged (Fig. 3f): 1) small tumors with low TDI, 2) small tumors with high TDI, and 3) large tumors with intermediate TDI. These three groups had significantly different survival rates (*χ*^’^ = 6.1977, *p* = 0.0451, two-sided log-rank test; Fig. 3g). Post-hoc comparisons revealed that patients with small tumors and low TDI had significantly longer survival compared to those with small tumors and high TDI (*χ*^’^ = 6.3676, *p* = 0.0116, two-sided log-rank test, *p* = 0.0349, FDR corrected). This subset of patients was also significantly younger than the rest (Fig. 3f). While both TDI and L-TDI were only weakly correlated with age at diagnosis (*ρ* = 0.10, *ρ* = 0.15, *p* < 0.01; two-sided exact test), age is a known modulator of OS, and this pattern would suggest that distinct mechanisms may underline GBM development across different age groups. Similar results were observed when clustering into four groups instead of three (Fig. S8f-h; Table S1) and were replicated in both the UCSF and UPENN cohorts independently (Fig. S9-10).

These three tumor subgroups also showed distinct anatomical distribution within the brain (Fig. 3i-j), aligning with prior findings (Mandal et al., 2020; Mandal et al., 2021; Salvalaggio et al., 2023), and offered a white-matter description of the non-uniform anatomical distribution of human gliomas. While two of the three clusters had statistically similar survival rates, their anatomical and morphological configuration differed markedly.

Specifically, small tumors with high TDIs were predominantly located in the left parietal lobe, while large tumors with medium TDIs were primarily found in the right frontal lobe. The percentage of affected brain lobes was significantly lower for the small tumor and high TDI, yet this anatomical sparing did not translate into improved prognosis (Fig. S11a). However, the corresponding L-TDMs of the two groups were largely equivalent (Kendall’s τ = 0.2049, *p* = 0.0592, two-sided exact test), showing substantial overlap with key eloquent white matter pathways such as the corticospinal tracts (Fig. 4k; Fig. S11b-c).

**Figure 4.**
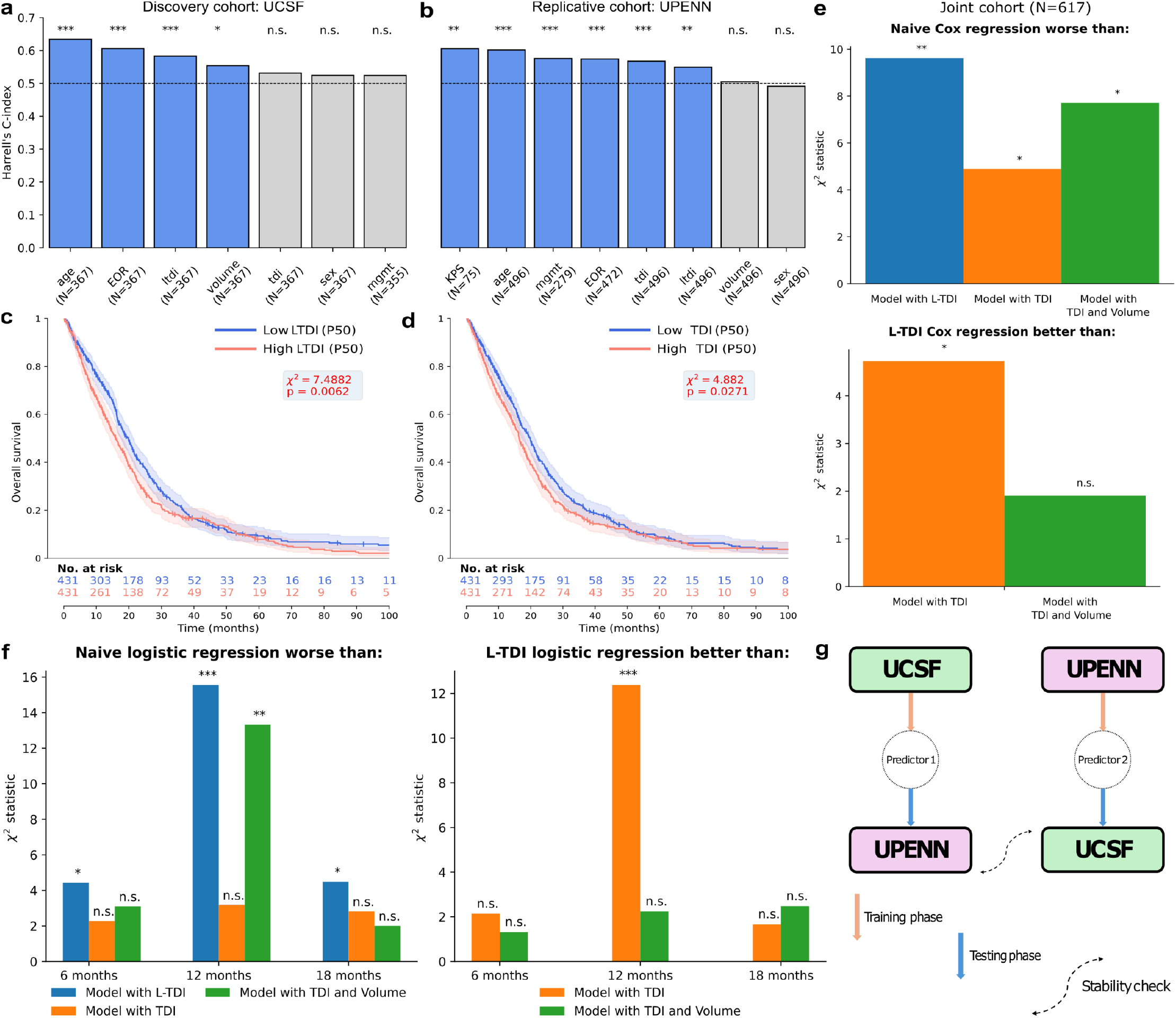
Predictive models of GBM survival using tract density-based biomarkers. **(a-b)** Harrell concordance index (*C*^*H*^) associated with every clinical covariate for the **(a)** UCSF and **(b)** UPENN cohorts. The dotted black lines show no discriminative power (i.e., expected at random). Blue and gray bars correspond to covariates with significant and non-significant indices, respectively (‘***’ *p* ≤ 0.001, ‘**’ *p* ≤ 0.01, ‘*’ *p* ≤ 0.05, ‘n.s.’ *p* ≥ 0.05; one-sided permutation tests; *n* = 1000 resamples). Features were sorted in descending order of importance (Table S2). **(c-d)** Kaplan-Meier curves for the two strata at the *p* = 50-th percentile of the **(c)** L-TDI and **(d)** TDI distributions. The group with low L-TDI (blue) is significantly more associated with longer survival times than the low TDI (red) group (*χ*^2^ = 7.4882, *p* = 0.0062; *χ*^2^ = 4.8820, *p* = 0.0271, two-sided log-rank test). The shaded areas correspond to the 95% confidence interval. Small vertical dashes indicate right-censored entries. **(e, top)** One-sided log-likelihood ratio tests comparing the Cox models with added covariates with respect to the *naive*. **(e, bottom)** *L-TDI* Cox model compared to the model with *TDI* and *TDI+volume* as added covariates of interest. All tests were done with 1 degree of freedom except when comparing the *TDI+Volume* and *Naive* models (*df* = 2). **(f, left)** One-sided log-likelihood ratio tests comparing the logistic regression with added covariates with respect to the *naive*. **(f, right)** *L-TDI* logistic regression compared to the model with TDI and *TDI+volume* as added covariates of interest. The positive class corresponded to the number of dead patients before 6, 12, and 18 months, respectively. **(g)** Schematic representation of the training and testing procedure used to validate a binary logistic classifier of deaths occurring 12 months after the initial diagnosis. All the survival curves and models were fit using the joint cohorts after accounting for site-related effects (see Methods).

In summary, the L-TDI and its spatial counterpart, the L-TDM, integrate information about tumor morphology, white matter density, and anatomical localization into a unified marker. This integrative approach also explained why tumors of different sizes resulted in similar survival outcomes, particularly when they involve similar white matter pathways despite differing in local tract densities (i.e., TDIs). These findings challenge conventional lesion-symptom mapping models and underscore the need to conceptualize GBM as a disorder of distributed white matter network involvement.

### 3.3. Multivariate Cox proportional hazard models of survival rates in human GBMs

We wanted to assess whether tract density-based markers significantly improved the prediction of survival rates compared to the habitual clinical variables, i.e., sex, age, MGMT status, EOR, and KPS. Initially, we computed Harrell’s concordance index (*C*^*H*^) of each clinical covariate, as well as tract-density indices and the tumor volume (Fig. 4a-b; Table S2). The L-TDI was associated with significant discriminative power in the UCSF (*C*^*H*^ = 0.5829, *p* < 0.0001, one-sided permutation tests; *n* = 1000 resamples) and UPENN (*C*^*H*^ = 0.5492, *p* = 0.002, one-sided permutation tests; *n* = 1000 resamples) cohorts. In contrast, the TDI retained statistical significance only in the UPENN cohort (*C*^*H*^ = 0.5677, *p* < 0.0001, one-sided permutation tests; *n* = 1000 resamples). Upon joining the two cohorts and stratifying the combined sample according to the *p* = 50-th percentile (e.g., the most conservative stratification threshold) of the L-TDI and TDI distributions, respectively, we confirmed the superior robustness of the L-TDI in (Fig. 4c-d).

To further quantify the prognostic value of tract density-based markers, we also fitted multiple Cox models to measure the respective hazard ratios (*HR*) in a multivariate context (Table 2). After pooling together the two cohorts and correcting for site-related differences (see Methods), both markers retained statistical significance, with the L-TDI showing a considerably higher contribution to survival (*HR* = 1.0742, *p* = 0.0050, two-sided Wald’s t-test) compared to the TDI (*HR* = 1.0026, *p* = 0.0250, two-sided Wald’s t-test). These results were confirmed after fitting the same Cox models to each cohort independently (Table S3). Amongst the classical clinical predictors, both the EOR and MGMT promoter status were associated with the largest *HRs*. Age, despite having relatively weaker *HRs*, proved to be the most robust indicator of decreased survival rates (Table S3-S7), i.e., it significantly participated in all the models and cohorts. Log-likelihood ratio (*LLR*) tests emphasized the added prognostic value of tract density-based imaging biomarkers (Fig. 4e; Fig. S12). Crucially, the *L-TDI* model was more plausible than the *TDI* one (*p* = 0.0296, one-sided *LLR* test), but not than the model that contained both TDI and volume as added features (*p* = 0.1674, one-sided *LLR* test). This could be understood because the L-TDI implicitly accounts for both local white matter density and lesion morphology (Fig. 3). These findings remained consistent after discarding 1) subjects without proper surgical resections, and 2) the EOR as a primary covariate (Fig. S13a-c; Tables S4-6).

**Table 2.** Multivariate Cox survival models. For the joint cohort (*N* = 617), after correcting for site effects and discarding missing values, hazard ratios (*e*^*βi*^) and the corresponding *p*-values (two-sided Wald’s t-test; *df* = *N* −*#covariates*). Bold numbers indicate significant hazard ratios (*p* < 0.05). The asterisks mark whether the *i*-th feature statistically contributes to 0, at least 1, or all the models (‘n.s.’, ‘*’, and ‘**’ respectively). *N*.*A*. stands for ‘not applicable’ due to the specifics of the Cox model fitted (see Methods). Table S3 includes the 95% confidence intervals and the results of each independent cohort.

| Model | Hazard Ratio ($e^{\beta_i}$) | | | | p-value | | | |
| --- | --- | --- | --- | --- | --- | --- | --- | --- |
|  | L-TDI | TDI | TDI+Volume | Naive | L-TDI | TDI | TDI+Volume | Naive |
| sex<sup>n.s.</sup> | 0.9501 | 0.9614 | 0.9570 | 0.9770 | 0.595 | 0.682 | 0.648 | 0.8084 |
| age<sup>**</sup> | **1.0276** | **1.0271** | **1.0273** | **1.0281** | **<0.001** | **<0.001** | **<0.001** | **<0.001** |
| mgmt<sup>**</sup> | **0.7270** | **0.7224** | **0.7241** | **0.7262** | **<0.001** | **<0.001** | **<0.001** | **<0.001** |
| EOR<sup>**</sup> | **0.5709** | **0.5644** | **0.5652** | **0.5472** | **<0.001** | **<0.001** | **<0.001** | **<0.001** |
| ltdi<sup>**</sup> | **1.0742** | N.A. | N.A. | N.A. | **0.005** | N.A. | N.A. | N.A. |
| tdi<sup>**</sup> | N.A. | **1.0026** | **1.0025** | N.A. | N.A. | **0.025** | **0.039** | N.A. |
| volume<sup>n.s.</sup> | N.A. | N.A. | 1.0009 | N.A. | N.A. | N.A. | 0.188 | N.A. |

We fitted a last multivariate Cox survival model on the subset of patients with available KPS in the UPENN cohort (*N* = 53; Fig. S13d; Table S7). Across all four models, age and MGMT covariates retained statistical significance in all 4 models. In contrast, the EOR remained borderline significant. In this subset, the L-TDI emerged as the strongest predictor of survival, showing the highest hazard ratio (*HR* = 1.3519, *p* = 0.005, two-sided Wald’s t-test). The KPS score was only significant in the 3 models where the L-TDI was not used. The TDI index did not provide significant predictive value in the two models where it was included, and the tumor volume’s hazard ratio was also not significant. Consistent with previous analyses, log-likelihood ratio tests showed that the *L-TDI* model significantly outperformed the *Naive* (*p* = 0.0050, one-sided *LLR* test), and *TDI* (*p* = 0.0292, one-sided *LLR* test), but not the *TDI+Volume* (*p* = 1.000, one-sided *LLR* test) models.

### 3.4. The L-TDI improves the prediction of survival probability

To estimate the probability of death within a specific timeframe, we used a logistic model suited for this binary classification task. First, using the entire joint cohort, we fitted a logistic model to predict the probability of a patient dying before 12 months from the date of diagnosis (Table 3). This approach provided insight into the predictive value of each feature. In line with the results obtained from the multivariate Cox models, the L-TDI significantly contributed to the prediction (*coef* = 0.2185, *p* < 0.001, two-sided Wald’s z-test), along with the traditional clinical features (i.e., age, MGMT status, and EOR). In contrast, the TDI remained borderline non-significant (*coef* = 0.0044, *p* = 0.077, two-sided Wald’s z-test). Tumor volume significantly added to the fit (*coef* = 0.0044, *p* < 0.003, two-sided Wald’s z-test). These results were consistent when considering each cohort independently (Table S8). Precisely, the L-TDI retained statistical significance in both cohorts, while the TDI only appeared significant in the UPENN cohort.

**Table 3.** Multivariate logistic regressions to estimate the probability of dying before 12 months. The logistic regressions were fitted to predict the probability of dying (status=1) before 12 months. Patients who were lost to follow-up (status=0) before 12 months were discarded. For the joint cohort (*N* = 546), after correcting for site eMects and discarding missing values, the coeMicients and the corresponding *p*-values (two-sided Wald’s z-test; *df* = *N* −*#covariates*). Bold numbers indicate significant coeMicients (*p* < 0.05). The asterisks mark whether the *i*-th feature statistically contributes to 0, at least 1, or all the models (‘n.s.’, ‘*’, and ‘**’ respectively). *N*.*A*. stands for ‘not applicable’ due to the specifics of the logistic regressions fitted (see Methods). Table S8 includes the 95% confidence intervals and the results of each independent cohort and multiple time horizons.

| Model | Coefficient for 12 months |  |  |  | p-value |  |  |  |
| --- | --- | --- | --- | --- | --- | --- | --- | --- |
|  | L-TDI | TDI | TDI+Volume | Naive | L-TDI | TDI | TDI+Volume | Naive |
| constant * | **-1.5277** | -1.1045 | **-1.6310** | -0.6189 | **0.048** | 0.156 | **0.043** | 0.392 |
| sex <sup>n.s.</sup> | -0.1230 | -0.1225 | -0.1088 | -0.0948 | 0.550 | 0.546 | 0.597 | 0.638 |
| age ** | **0.0546** | **0.0545** | **0.0548** | **0.0563** | **<0.001** | **<0.001** | **<0.001** | **<0.001** |
| mgmt ** | **-0.2976** | **-0.2905** | **-0.2997** | **-0.2926** | **0.004** | **0.004** | **0.003** | **0.004** |
| EOR ** | **-0.967** | **-1.0188** | **-1.027** | **-1.0696** | **<0.001** | **<0.001** | **<0.001** | **<0.001** |
| ltdi ** | **0.2185** | N.A. | N.A. | N.A. | **<0.001** | N.A. | N.A. | N.A. |
| tdi <sup>n.s.</sup> | N.A. | 0.0044 | 0.0043 | N.A. | N.A. | 0.077 | 0.093 | N.A. |
| volume <sup>n.s.</sup> | N.A. | N.A. | **0.0044** | N.A. | N.A. | N.A. | **0.003** | N.A. |

We extended this framework to account for potential differences in the time courses of survival rates by predicting the probability of dying before 6 and 18 months, respectively (Table S8). The corresponding *LLR*s tests confirmed that logistic fits were statistically more plausible when incorporating explicitly the L-TDI jointly with the habitual clinical markers (Fig. 4f). Importantly, models based on the TDI did not outperform the naïve baseline, whereas those including the L-TDI consistently showed superior predictive performance across all time windows. Additionally, it offered a considerably better fit than the *TDI* model at 12 months (*p* = 0.0004, one-sided *LLR* test). As before, the combination of TDI and lesion volume approximated the performance of the *L-TDI* model (Fig. 4f right), further reinforcing that the L-TDI captures both features in a compact representation.

Finally, to assess the real-world applicability and generalizability of the L-TDI, we leveraged the two independent cohorts to construct cross-site predictive models. Among several modeling options, we intentionally opted for a simple logistic classifier to test the robustness of the proposed marker explicitly. We trained and tested models across the UCSF and UPENN cohorts, then repeated the procedure by swapping their roles (Fig. 4g), automatically readjusting cross-validation parameters to optimize training (Methods; Fig. S9). Since survival prediction is inherently imbalanced (i.e., the number of deceased subjects increases over time), we used the area under the curve (AUC) and balanced accuracy (bACC) as evaluation metrics. The performance of the models was stable when considering the different combinations of training and testing cohorts, with the L-TDI model consistently outperforming the *TDI* and *Naive* models, albeit modestly (Table 4). Results were comparable when predicting death at 6 and 18 months (Fig. S9; Table S10), suggesting that including L-TDI enhances model performance as a relevant imaging feature.

**Table 4.** Classification metrics for the logistic classifier trained to predict mortality within 12 months. Area under the curve (AUC) and balanced accuracy (bACC) of the models tested on each independent cohort. *N*.*A*. stands for not applicable, since we did not perform testing on the cohort used for training. Bold numbers indicate the best model on the testing cohort.

| TRAIN (12 months) |  |  |  |  |  |  |  |
| --- | --- | --- | --- | --- | --- | --- | --- |
| Area under the curve (AUC)<br>*Model* |  | UCSF |  |  | UPENN |  |  |
|  |  | *L-TDI* | *TDI* | *Naive* | *L-TDI* | *TDI* | *Naive* |
| TEST (12 months) | UCSF | N.A. |  |  | **0.7433** | 0.7088 | 0.7220 |
|  | UPENN | **0.7327** | 0.7039 | 0.7077 | N.A. |  |  |
| Balanced accuracy (bACC)<br>*Model* |  | UCSF |  |  | UPENN |  |  |
|  |  | *L-TDI* | *TDI* | *Naive* | *L-TDI* | *TDI* | *Naive* |
| TEST (12 months) | UCSF | N.A. |  |  | **0.6498** | 0.6463 | 0.6418 |
|  | UPENN | **0.6753** | 0.656 | 0.6661 | N.A. |  |  |

## 4. Discussion

In this work, we provide a robust and reproducible stratification of GBM survival outcomes by leveraging a novel Lesion-Tract Density Index (L-TDI) marker, thus presenting compelling evidence that GBM prognosis is fundamentally influenced by the underlying architecture of the brain’s white matter. Crucially, a newly emerging paradigm suggests that GBM interferes with the brain’s circuitry (Tetzlaff et al., 2025), consequently altering various critical functions (Sadanand, 2019; Duffau and Filippi, 2024). In agreement with it, our findings reinforce the view that GBM is not solely a focal pathology, but a disease of large-scale structural involvement, whose clinical impact is governed by its interaction with the brain’s network infrastructure. Prior studies have shown that tumor volume alone does not reliably predict survival (Bette et al., 2017; Henker et al., 2019; Johnstad et al., 2024). Our findings challenge the conventional focus on tumor size or gray matter location and instead highlight the critical importance of the underlying white matter network. As such, GBMs should not be viewed merely as a localized mass disconnecting distant regions, but rather as a network-level disease, involving large-scale white matter organization and, consequently, impacting global brain communication (Krishna et al., 2023; Sun et al., 2025).

A major contribution of this study lies, therefore, in establishing the L-TDI as a marker that simultaneously captures multiple critical GBM dimensions, its volume, spatial distribution, and potential infiltration, therefore offering a white matter-informed perspective on glioma localization. In line with prior work, our findings reinforce that GBMs are not randomly distributed across the brain but preferentially arise in regions of distinct white matter density (Mandal et al., 2021). Yet, unlike traditional volumetric or anatomical metrics, the L-TDI encodes how the tumor is embedded within the broader scaffold of white matter tracts, fundamentally moving beyond lesion-symptom associations. The morpho-anatomical model of the current study reinforces the idea that tumor topography is an emergent property of the structural connectivity landscape of the brain. Indeed, although small tumors with high TDI affected a smaller portion of the brain in terms of lobe involvement compared to large GBMs with an average TDI (Fig. 3; Fig. S11a), this anatomical sparing did not translate into improved prognosis. This underscores a critical point: the clinical impact of a GBM is not solely determined by its size or anatomical topography but by the interaction between the tumor and surrounding white matter pathways. Accordingly, the spatial patterns of white matter involvement of these two subgroups were remarkably similar, indicating that both tumor groups converged in key white matter structures. Hence, our findings suggest that clinical evolution might benefit from augmenting therapeutic strategies with non-local targets.

In addition to clarifying the non-trivial relationship between the tract density markers, morphology, and anatomy in GBM, we validated the prognostic value of L-TDI using multivariate Cox regression models, demonstrating that its contribution to OS prediction remains significant even after accounting for well-established clinical and molecular variables, including MGMT promoter methylation status. The additive power of L-TDI in these models underscores its utility as a non-redundant biomarker that complements, rather than replaces, current prognostic tools. Moreover, L-TDI offers a mechanistic bridge between macroscopic lesion burden and microscale tumor invasiveness, reflecting the involvement of long-range communication pathways critical to brain function and resilience. Ultimately, radiotherapy planning might benefit from targeting this glioma-neuron scaffold (Duffau and Filippi, 2024).

Importantly, the L-TDI possessed predictive value. A simple logistic regression achieved high accuracy in predicting 12-month mortality, both within the original sample and when applied to an independent validation cohort. This cross-site generalizability is essential for clinical translation and suggests that the L-TDI framework could be readily deployed within existing neuroimaging pipelines. Furthermore, this retrospective study draws on one of the largest cohorts reported in recent literature, strengthening the statistical power and generalizability of the results. Such large-scale efforts are key to fostering open clinical collaboration and establishing multi-center networks that can improve the reliability and translational potential of findings in GBM management (Cho et al., 2023).

Lastly, we acknowledge that using normative templates may be perceived as a limitation. Given the known tendency of gliomas to infiltrate, displace, and degrade surrounding tissue, normative atlases might under-or overestimate local white matter involvement. However, this approach offers important advantages, particularly in the context of standardized analysis, reproducibility, and computational tractability. In clinical environments, where patient-specific diffusion imaging is often noisy or unavailable, normative L-TDI provides a viable and informative proxy. That said, a crucial direction for future research will be validating L-TDI within a patient-specific tractography framework. This would allow us to assess how accurately the normative-based L-TDI reflects individual white matter anatomy in the presence of tumor-induced distortion. Nevertheless, performing diffusion tractography in the presence of brain tumors is a technically challenging task (Yeh et al., 2021; Fekonja et al., 2022; Falcó-Roget et al., 2024), and any potential gains must be weighed against the cost and complexity of acquiring high-quality diffusion data in a clinical context.

## 5. Conclusions

Our findings establish a new lesion-tract density index (L-TDI) as a robust and clinically meaningful biomarker that captures the complex interplay between tumor morphology and white matter architecture. By integrating tumor volume, spatial distribution, and normative tract density into a single index, this L-TDI offers a biologically grounded explanation for survival differences that are not accounted for by traditional metrics. Unlike purely local features, the L-TDI reflects the extent to which a tumor interacts with critical white matter circuitry, providing insights into disease prognosis. It consistently outperformed or matched alternative models across multiple statistical frameworks and independent cohorts, while maintaining interpretability and feasibility using normative connectomes. As such, this work represents a step forward toward connectomics-guided neuro-oncology, with the potential to inform individualized risk stratification, clinical decision-making, therapeutic planning, and ultimately, patient outcomes.

## Data availability

The discovery dataset was retrieved from The University of California San Francisco Preoperative Diffuse Glioma MRI repository in The Cancer Imaging Archive (Calabrese et al., 2022; Calabrese et al., 2022). The replicative dataset was retrieved from the Multi-parametric magnetic resonance imaging (mpMRI) scans for de novo Glioblastoma (GBM) patients from the University of Pennsylvania Health System collection in The Cancer Imaging Archive (Bakas et al., 2022; Bakas et al., 2021). The normative connectome employed in the present work is also freely available (Elias et al., 2024; Lozano et al., 2024). The USCLobes brain atlas can be downloaded from: https://brainsuite.org/usclobesatlas/ (Joshi et al., 2022). The XTRACT white matter atlas is openly accessible in FSL (Warrington et al., 2020). We previously registered both parcellations to the same MNI template used in this study, following the same steps outlined in the Methods.

## Code availability

The codes to perform the normalization to MNI, computation of the tract density indices, and the statistical analyses will be made openly available and, in the meantime, can be requested from the corresponding authors. A combination of custom codes and software packages was used. Specifically, survival analyses were carried out within the statsmodels (Seabold and Perktold, 2010), Scikit-survival (Pölsterl, 2020), and lifelines (Davidson-Pilon, 2024) Python packages. All the figures in the main and supplementary text were generated with custom Python codes, saved as scalable vector graphics (SVG) files, and assembled in Inkscape.

## Author contributions

*Conceptualization:* J.F.-R. and A.C. *Methodology:* J.F.-R. and A.C. *Software:* J.F.-R. *Validation:* J.F.-R., G.A.B., A.J., S. L., L.S.P., and A.C. *Formal analysis:* J.F.-R. and A.C. *Investigation:* J.F.-R. *Resources:* J.F.-R., J.K.A., and A.C. *Data Curation:* J.F.-R. and A.C. *Writing – Original Draft:* J.F.-R. and A.C. *Writing – Review & Editing:* J.F.-R., G.A.P, A.J., S.L., L.S.P, J.K.A., and A.C. *Visualization:* J.F.-R., G.A.P., A.J., and A.C. *Supervision:* A.C. *Project administration:* J.F.-R., J.K.A., and A.C. *Funding acquisition:* J.F.-R., J.K.A., and A.C.

## Acknowledgments

J.F-.R., A.J., and J.K.A. thank Cemal Koba, PhD for insightful discussions.

