## Supplementary Materials for "A non-local diffusion magnetic resonance imaging tract density biomarker to stratify, predict, and interpret survival rates in human glioblastoma"

[1]Computational Neuroscience Group, Sano Centre for Computational Medicine, Kraków, Poland. [2]Brain Mapping Lab, Department of Biomedical, Dental Sciences and Morphological and Functional Imaging, University of Messina, Messina, Italy. [3]Department of Neurophysiology and Chronobiology, Institute of Zoology and Biomedical Research, Faculty of Biology, Jagiellonian University, Krakow, Poland. [4]Radiation Oncology Unit, Clinical Department, National Center for Oncological Hadrontherapy (CNAO), 27100, Pavia, Italy. [5]Department of Internal Medicine and Medical Therapy, University of Pavia, 27100 Pavia, Italy. [6]Department of Biomedical Sciences, Humanitas University, Via Rita Levi Montalcini 4, Pieve Emanuele, 20072 Milan, Italy. [7]IRCCS Humanitas Research Hospital, Via Alessandro Manzoni 56, Rozzano, 20089 Milan, Italy. [8]Faculty of Physics, Astronomy and Applied Computer Science, Jagiellonian University, Krakow, Poland.

[†]Equal contribution: Gianpaolo Antonio Basile, Anna Janus

**Abstract:** This supplement contains an extended version of the Methods. We present all the steps of the pre- and post-processing of the imaging and survival data used in this study. The structure of this document closely follows the one in the main document.

### 1. Image normalization to the MNI template

For both cohorts, the structural and multi-label lesion segmentations were already co-registered (i.e., aligned in subject space). Since our study relied on normative models of white matter, we nonlinearly normalized all the necessary images to the Montreal Neurological Institute (MNI ICBM 2009b NLIN Asymmetric; hereafter referred to as "MNI template") standard space (Elias et al., 2024). The structural modalities T1, T1 contrast enhanced, T2, and FLAIR images were corrected for bias field homogeneities to facilitate and improve the normalization in the UPENN cohort (Tustison et al., 2010). This same correction step was already available for the UCSF patients.

Before normalizing, we extracted the brain tissue of the MNI template (Smith, 2002). Additionally, for both cohorts, we binarized and inverted the multi-label lesion segmentations with FSL (FMRIB Software Library; (Jenkinson et al., 2012)) and ANTs (Advanced Normalization Tools[1]; (Avants et al., 2009)). These steps ensured that the registration workflow used only brain tissue of the target space and non-lesioned brain tissue in the subject image. Then, we nonlinearly mapped the T2 images to the T2 MNI template (Avants et al., 2008). This consisted of a rigid, followed by an affine, and a symmetric normalization (SyN) registration with histogram matching (Fig. 1a). The choice of T2 as the reference modality was motivated by a subject-wise higher image

[1] https://stnava.github.io/ANTs/

quality, though the registration results were independent of this choice. The resulting transformation was then sequentially applied to the remaining structural modalities and the multi-label lesion segmentations. As a last step, the normalized images were overlaid onto the MNI template to visually assess the alignment of anatomical landmarks (e.g., sulci and gyri). Automated contour lines were also drawn to highlight the correspondence of the ventricular structures. The volume of the lesion was computed in cubic centimeters ($\mathrm{cm}^3$) by multiplying the number of voxels of the binary masks in MNI space and the factor $0.5^3 \cdot 10^{-3}$ ($\mathrm{cm}^3/\mathrm{voxel}$).

### 2. Normative tractograms

To evaluate the white matter pathways interacting with the tumor lesion, we sought to employ a publicly available normative structural connectome, which is a group-level tractogram capturing the common white matter anatomy of a representative population. In that sense, it can be taken as a proxy for "normal" white matter structures. Specifically, we used an average tractogram of 985 subjects from the Human Connectome Project (Elias et al., 2024). Briefly, for every subject, deterministic diffusion fiber tractography was performed using generalized Q-sampling imaging in DSI Studio, thus resolving crossing fibers (Yeh et al., 2013). The resulting tractogram was nonlinearly transformed to the same MNI template previously described. Then, 12,000 streamlines were randomly sampled from the 250,000 originally reconstructed. The final tractogram was obtained by aggregating the subsampled tractograms, thus creating a normative model of 11,820,000 streamlines.

To compute a patient-specific lesion-tract density map (L-TDM) and index (L-TDI), for every patient, we selected the streamlines that intersected the lesion mask using the "include" option in the "tckedit" command in MRtrix3 (Tournier et al., 2019). From this lesion-tractogram encoding the normative pathways that were likely interacting with the tumor (Fig. 1b-c), we obtained the L-TDM by mapping the number of lesion-streamlines traversing each voxel (Calamante et al., 2010). To obtain the corresponding L-TDI, we masked all voxels of the L-TDM with a value higher than 0 (i.e., voxels with at least one streamline) and computed the average value within this mask (Fig. 1d). In summary, the L-TDI considered the average white matter density of the whole brain-lesion normative circuit, thus explicitly modeling GBM as a distributed white-matter lesion rather than a focal disruption.

For completeness, we also obtained the tract density map (TDM) and index (TDI) of every patient (Salvalaggio et al., 2023). From the full normative tractogram with 11,820,000 streamlines, we created a density map capturing the average number of streamlines traversing every voxel (Calamante et al., 2010). Then we overlayed the resulting map and every tumor mask to create a patient-specific TDM. Lastly, the TDI was obtained by averaging the TDM within the tumor mask. Therefore, the TDI considered the local (within the tumor) density of potentially infiltrated white matter

### 3. Survival analysis and statistical procedures

We treated the UCSF cohort as a discovery dataset and the UPENN cohort as a replicative dataset. This ensured that our results were not tailored to a specific data distribution. Initially, for the discovery cohort, we obtained the L-TDI distribution and stratified the sample into two groups based on an arbitrary percentile. Specifically, patients with an L-TDI smaller than and higher than the $p$-th and $(100 - p)$-th percentiles were assigned to the "Low L-TDI" and "High L-TDI" groups, respectively. To prove that our approach was robust and threshold-independent, the stratification threshold (i.e., percentile) was uniformly changed from 20 to 50 in steps of 5, and we compared the Kaplan-Meier curves for the two groups via two-sided log-rank tests (Kaplan and Meier, 1958; Fleming and Harrington, 1981). The *p*-values were later corrected for multiple comparisons (Benjamini and Hochberg, 1995). Based on the previous stratification thresholds (defined from tract density-based markers) we evaluated the median OS for patients known to die (discarding right-censored entries). Statistical significance was assessed through two-sided Mann-Whitney U-tests. The resulting *p*-values, one for each stratification threshold, were later controlled for multiple hypothesis inflation using the False Discovery Rate (FDR) procedure. This proved that the differences in survival rates in the Kaplan-Meier curves were truly observable.

Then, we computed the average L-TDI and TDI for patients who died and survived beyond a given temporal threshold. The subgroup average indices were systematically compared via two-sided Mann-Whitney U-tests and corrected for multiple comparisons using the FDR correction. For these analyses, we evaluated the separability of the two classes at $6, 12, 18, 24, 30, 36, 42$, and $48$ months. Additionally, for the same survival thresholds, we computed the Pearson correlation ($\rho$) between survival times and (L-)TDIs. More specifically, we selected the patients with survival times lower than a threshold (e.g., patients that died before the first 6 months) and correlated this subset of survival times with the corresponding (L-)TDIs. Significance was assessed via two-sided permutation tests ($n = 2500$ resamples). Given that multiple survival thresholds were tested, we also corrected for multiple comparisons using the FDR correction.

Furthermore, we computed the receiver operating curves (ROCs), using only the L-TDI values without stratifying the patients, to discriminate between dead from alive patients both at $12$ and $24$ months. We used Youden's J index to obtain the corresponding balanced accuracy (bACC) (Youden, 1950). To conclude this part of the analysis, we also evaluated the discriminative power of the tract density markers as a function of time with a cumulative and dynamic time-dependent AUC (Hung and Chiang, 2009). This metric was able to distinguish between patients who experienced failure (i.e., death) before and after a given time. We used a repeated K-fold cross-validation scheme with $50$ instances to estimate the censoring distribution and computed the time-dependent AUC on the corresponding test samples. The time was sampled from $6$ to $36$ months for a different number of cross-validation splits, namely $2, 6$, and $8$. For example, 6-fold cross-validation means that the model was trained jointly on 5 out of 6 splits of the data and validated on the remaining one.

All the previous analyses were then repeated in the same way for the replicative cohort

### 4. Dimensionality reduction and clustering of macroscopic features in GBMs

To understand the relationship between tract density indices and tumor volume, we performed linear principal component (PC) analyses with the TDI, L-TDI, and volume as features of interest. We then obtained the corresponding PC loadings and performed a linear fit between the first two principal components and the L-TDI. This was motivated by the observation that the 1st and 2nd PCs jointly accounted for more than $99\%$ of the explained variance. The goodness of the fit was assessed with the determination coefficient.

Based on the previous findings, we performed automatic clustering of patients based on the TDI and tumor volume with an iterative *K*-means algorithm (Lloyd, 1982). The maximum number of iterations was 300. In what follows, each of the 3 found clusters was analyzed separately. First, we computed the corresponding Kaplan-Meier curves. Second, we computed the tumor maps and visualized them through maximum intensity projection plots. Third, we computed the overlap of every brain lobe delineated in the USCLobes brain atlas (Joshi et al., 2022) with the tumor mask and obtained the averages across subjects. This allowed us to refine the anatomical labeling of the clusters that were automatically discovered by assessing the percentage of every brain lobe inside the tumor, i.e., the percentage of the tumor mask that corresponded to every brain lobe. Lastly, we computed the overlap of the corresponding L-TDMs and every white matter tract delineated in the XTRACT atlas (Warrington et al., 2020). Once again, we obtained the average across subjects to obtain a description of major white matter tracts intersected by the L-TDI maps in every cluster. Importantly, this overlap was not computed with the tumor masks, but rather the underlying white matter profiles of the lesions, thus moving beyond traditional lesion-symptom mappings. The resulting overlaps were then sorted in descending order and compared between clusters using both Spearman correlation and Kendall's $\tau$ coefficients.

### 5. Joining survival data for improved hazard ratio estimates in multivariate Cox models

#### 5.1. Cox models for each independent cohort

To further assess the added value of tract density-based markers in the clinical context, we used several Cox proportional hazard models (Cox, 1972; Breslow, 1975). For each cohort separately, we fitted 1) a *naïve model* containing only the classical covariates of interest (i.e., sex, age, MGMT promoter status, and EOR); 2) an *L-TDI model* containing the classical covariates of interest and the L-TDI; 3) a *TDI model* containing the classical covariates of interest and the TDI; and 4) a *TDI+Volume model* containing the classical covariates of interests, the TDI, and the lesion volume in cubic centimeters in the template space. Model comparisons were carried out through log-likelihood ratio tests. The significance of the hazard ratio for each covariate was assessed with a two-sided Wald test and the associated T distribution. The corresponding 95% confidence intervals were also calculated.

### 5.2. Method to join survival data

We fitted the hazard ratios using the maximum likelihood estimator, which crucially depends on the total sample size $N$; the corresponding biases and confidence intervals decrease with $\sim 1/\sqrt{N}$, and the variance with $\sim 1/N$. In the case of survival data, this dependency is even more restrictive due to the presence of right censoring (Harrell, 2015), justifying joining the cohorts used in this work. This effectively doubled the sample size (from 367 and 496 to 863). However, since the survival was measured from days of diagnosis and from days of surgery in the UCSF and UPENN cohorts, respectively, we accounted for site-related differences in survival rates.

In a Cox proportional hazards model, for every patient $p$, the hazard ratio $\lambda(t)$ is parametrized with an exponential linear combination of all the selected covariates of interest,

$$\lambda\left(t, S_p, \boldsymbol{x}_p, \boldsymbol{\beta}\right) = \lambda_0 \cdot \exp\left(\beta_s \cdot S_p + \sum\nolimits_{i=1}^{M} \beta_i \cdot x_{p,i}\right), \tag{S5.1}$$

where $S_p = \{0,1\}$ for the UCSF and UPENN cohorts, respectively, $\boldsymbol{X} \doteq (\boldsymbol{x}_1, \ldots, \boldsymbol{x}_N)$ is a matrix of $N \times M$ covariates, $\boldsymbol{\beta}$ is the $1 \times M$ vector of log-hazard ratios, and $\lambda_0$ is the baseline hazard rate. For each patient, we then obtained the site-corrected OS

$$\tilde{t}_p = t_p \cdot e^{\beta_s \cdot S_p}, \tag{S5.2}$$

where and $\beta_s$ is the covariate estimated from Eq. (S5.1). Then, we fitted a second Cox model

$$\tilde{\lambda}\left(t, \tilde{\boldsymbol{x}}_p, \tilde{\boldsymbol{\beta}}\right) = \tilde{\lambda}_0 \cdot \exp\left(\sum\nolimits_{i=1}^{M} \tilde{\beta}_i \cdot \tilde{x}_{p,i}\right), \tag{S5.3}$$

where $\tilde{\boldsymbol{X}} = (\tilde{\boldsymbol{x}}_1, \ldots, \tilde{\boldsymbol{x}}_N)$, and $\tilde{\boldsymbol{\beta}}$ are the site-corrected matrix of covariates and the corresponding site-corrected log-hazard ratios. The site-corrected hazard rates $\tilde{\lambda}\left(t, \tilde{\boldsymbol{X}}, \tilde{\boldsymbol{\beta}}\right)$ and $\tilde{\lambda}_0$ were estimated from the site-corrected survival times $\{\tilde{t}_p\}_{p=1,\ldots,N}$ from Eq. (S5.2), contained in $\tilde{\boldsymbol{X}}$. Similarly, the Kaplan-Meier estimators of the survival rates $\hat{S}(t)$ were calculated from the site-corrected times for the joint cohort.

Lastly, Harrell's concordance $(C^H)$ index has the same interpretation as the AUC (Harrell, Lee and Mark, 1996), with values $\leq 0.5$ indicating a worse or equal than random discriminative power. Values significantly higher than $0.5$ indicated that the feature could be used to discriminate differences in survival rates. Although the $C^H$ index can be positively biased for large percentages of right censoring, (Uno et al., 2011), this bias is negligible for percentages below 50-60% of censored samples[2].

### 5.3. Effect of the transformation $\tilde{t}$ in the hazard rate

Let's suppose that we scale the time measurements by the hazard ratio $(HR)$ associated with the covariate of site:

$$\tilde{t} = t \cdot e^{\beta_s \cdot S_p}, \tag{S5.4}$$

whose Jacobian is constant,

---

[2] See also: https://scikit-survival.readthedocs.io/en/stable/user_guide/evaluating-survival-models.html

$$J \doteq \left|\frac{d\tilde{t}}{dt}\right| = e^{\beta_s \cdot S_p}. \quad \text{(S5.5)}$$

From standard probability theory (Bishop and Nasrabadi, 2006), the transformed hazard ratio is scaled by the inverse of the Jacobian,

$$\lambda\left(\tilde{t}, S_p, \boldsymbol{x}_p, \boldsymbol{\beta}\right) = J^{-1} \cdot \lambda\left(t, S_p, \boldsymbol{x}_p, \boldsymbol{\beta}\right). \quad \text{(S5.6)}$$

Substituting Eqs. (S5.1), (S5.4), and (S5.5) to Eq. (S5.6), we obtain the transformed hazard ratio, which turns out to be independent of the site and equal to Eq. (S5.3).

$$\lambda\left(\tilde{t}, \boldsymbol{x}_p, \boldsymbol{\beta}\right) = \lambda_0(\tilde{t}) \ \cdot \exp\left(\sum_{i=1}^{M} \beta_i \cdot x_{p,i}\right),$$

where $\lambda_0(\tilde{t})$ is the transformed baseline hazard ratio. Therefore, we have unified the survival times of both cohorts.

Noteworthy, applying the transformation in Eq. (S5.2) requires estimating the log-$HR$ of the site covariate. Fortunately, Cox proportional models, like the one in Eq. (S5.1), can be readily used provided that the site complies with the proportional hazard assumption (Fig. S3). From a practical point of view, we observed that estimating $\beta_s$, transforming the survival times using Eq. (S5.2), and then fitting a second Cox model to estimate the new log- $HRs$ of the rest of the covariates, returned lower log-likelihoods ratios compared to applying Eq. (S5.6) out-of-box. This resembled established methodologies in multi-site neuroimaging studies (Fortin et al., 2017). However, the procedure above should be applied (for now) in controlled settings where an a *priori* hypothesis exists to explain systematic differences in survival rates. Of utmost importance is to thoroughly verify whether these differences can be accounted for within a constant proportional hazard assumption. In our cohorts, survival was measured from different dates. In one case, counting from the date of diagnosis, and in the other, counting from the date of surgery. This systematic difference could be described perfectly with a proportional hazard assumption (Fig. S3), thus allowing us to combine both cohorts for improved estimation routines.

A useful (fake) scenario to understand why we could join the survival times of the two cohorts we studied is to imagine that overall survival had been measured in days (cohort 1) and months (cohort 2). Obviously, the survival curves of these two cohorts could not be compared without first transforming the survival times of one of these cohorts into the units of the other one, e.g., recording survival in days for the samples in cohort 2. This can be trivially obtained by simply using the equivalence factor between months and days:

$$\text{days} = \frac{365}{12} \cdot \text{months} \quad \text{(S5.7)}$$

Notice how the transformation above is formally identical to Eq. (S5.2), thus implying that changing the measurement units translates into proportional rescaling of the survival rates.

#### 5.4. Effect of the transformation $\tilde{t}$ in the Kaplan-Meier survival estimator

The effect of the transformation in Eq. (S5.2) on the Kaplan-Meier estimator can also be described and is like what was described above. To be precise,

$$\hat{S}(t) = \prod_{t_i \leq t} [1 - h(t_i)] = \prod_{t_i \leq t} \left[1 - \frac{d(t_i)}{n(t_i)}\right], \quad \text{(S5.8)}$$

with $h(t_i)$ being the instantaneous hazard rate, $d(t_i)$ being the number of failures (i.e., deaths) at time $t_i$ and $n(t_i)$ being the number of subjects that did not experience and even at time $t_i$. Both the instantaneous hazard rate and the Kaplan-Meier estimators are functions of the available survival data $\{t_i\}_{i=1,\dots,N}$ where $N$ is the samples size.

Straightforwardly applying the same transformation in Eq. (S5.2), estimated with the corresponding Cox model, implies modifying the data as follows:

$$\left\{\tilde{t}_i = t_i \cdot e^{\beta_s \cdot S_p}\right\}_{i=1,\ldots,N}, \tag{S5.9}$$

This transformation propagates onto the estimator in a controllable manner:

$$\begin{aligned} \hat{S}(t,\{\tilde{t}_i\}) &= \prod_{t_i \cdot e^{\beta_s \cdot S_p} \le t} \left[1 - h(t,\{\tilde{t}_i\})\right] \\ &= \prod_{t_i \le t \cdot e^{-\beta_s \cdot S_p}} \left[1 - h\left(t \cdot e^{-\beta_s \cdot S_p},\{\tilde{t}_i\}\right)\right] \\ &= \hat{S}\left(t \cdot e^{-\beta_s \cdot S_p},\{\tilde{t}_i\}\right) \\ &= \hat{S}(\tilde{t},\{\tilde{t}_i\}) \end{aligned} \tag{S5.10}$$

Thus, transforming the individual measurement times is equivalent to evaluating the transformed estimator at a time rescaled by the transformation factor. In summary, the transformation we applied effectively rescales the Kaplan-Meier estimators without modifying the instantaneous probabilities of failure within the rescaled cohort.

### 6. Logistic regression modeling of patient survival and death predictions

We fitted a logistic regression to evaluate the contribution of each feature to the outcome at a given time (i.e., 1 died and 0 survived). As per the Cox proportional modelling, we fitted the same four logistic models to inspect the contribution of tract density-based markers. Then, we recorded the significance of each covariate, the confidence interval, as well as the model log-likelihood. Models were then compared via log-likelihood ratio tests. We trained these models for each separate cohort, but given that the logistic regression was fit using maximum likelihood estimation, we also joined the two cohorts to improve the stability of the final estimates.

Additionally, to predict whether a given patient would die before a given number of months, we trained a logistic classifier. The training was performed using repeated stratified cross-validation using a grid-search scheme varying both the number of splits (from 2 to 12) and repeats (from 5 to 25). We recorded the AUC and the bACC at every combination and selected the parameters with the minimum cost,

$$-\lambda \cdot (\mathrm{AUC} + \mathrm{bACC}). \tag{4}$$

We opted for $\lambda = 0.5$ to consider both class separability and true positive rates. Finally, we trained the logistic classifier with the selected parameters from scratch for the same cohort and tested it on the independent one. This procedure was repeated for 6, 12, and 18 months using each cohort as training and testing sets. Importantly, survival times were previously corrected for site dependencies as described earlier.

All the procedures described here were carried out after discarding the samples with missing data.
