## Supplementary Tables for "A non-local diffusion magnetic resonance imaging tract density biomarker to stratify, predict, and interpret survival rates in human glioblastoma"

[1]Computational Neuroscience Group, Sano Centre for Computational Medicine, Kraków, Poland. [2]Brain Mapping Lab, Department of Biomedical, Dental Sciences and Morphological and Functional Imaging, University of Messina, Messina, Italy. [3]Department of Neurophysiology and Chronobiology, Institute of Zoology and Biomedical Research, Faculty of Biology, Jagiellonian University, Krakow, Poland. [4]Radiation Oncology Unit, Clinical Department, National Center for Oncological Hadrontherapy (CNAO), 27100, Pavia, Italy. [5]Department of Internal Medicine and Medical Therapy, University of Pavia, 27100 Pavia, Italy. [6]Department of Biomedical Sciences, Humanitas University, Via Rita Levi Montalcini 4, Pieve Emanuele, 20072 Milan, Italy. [7]IRCCS Humanitas Research Hospital, Via Alessandro Manzoni 56, Rozzano, 20089 Milan, Italy. [8]Faculty of Physics, Astronomy and Applied Computer Science, Jagiellonian University, Krakow, Poland.

[†]Equal contribution: Gianpaolo Antonio Basile, Anna Janus

| ***(Volume, TDI)*** | (Small, High) | Mixture group | (Small, Small) | (Large, Medium) |
|---|---|---|---|---|
| (Small, High) | *N.A.* | **(3.9989, 0.0455, 0.1366)** | **(10.8932, 0.001, 0.0058)*** | (1.8803, 0.1703, 0.2044) |
| Mixture group | *N.A.* | *N.A.* | (1.922, 0.1581, 0.2044) | 0.2277,0.6333, 0.6333) |
| (Small, Small) | *N.A.* | *N.A.* | *N.A.* | (2.6595, 0.1029, 0.2044) |
| (Large, Medium) | *N.A.* | *N.A.* | *N.A.* | |

**Table S1. Post-hoc comparisons between survival across the different 4 clusters.** Results of the pairwise comparisons between Kaplan-Meier curves ($\chi^2$, $p$, $p_{FDR}$, two-sided log-rank test, FDR correction). Each cluster was automatically detected in the 2-dimensional space formed by the TDI and volume of the tumor. The details on the clustering algorithm can be found in the methods section of the main text. Bold numbers indicate significant differences in survival rates, and * depicts comparisons that survived the false discovery rate correction. This table is a companion to Fig. S8f-h.

| ***Feature*** | **Joint** | | | | **UCSF** | | | | **UPENN** | | | |
|---|---|---|---|---|---|---|---|---|---|---|---|---|
| | C-index | P value | Rank | N | C-index | P value | Rank | N | C-index | P value | Rank | N |
| sex | 0.5017 | 0.762 | 7 | 863 | 0.5246 | 0.200 | 6 | 367 | 0.4914 | 0.989 | 8 | 496 |
| age*** | **0.6139** | **<0.001** | 1 | 863 | **0.6343** | **<0.001** | 1 | 367 | **0.6018** | **<0.001** | 2 | 496 |
| mgmt* | **0.5537** | **<0.001** | 5 | 634 | 0.5143 | 0.170 | 7 | 355 | **0.5756** | **<0.001** | 3 | 279 |
| EOR** | **0.5881** | **<0.001** | 2 | 839 | **0.6058** | **<0.001** | 2 | 367 | **0.5740** | **<0.001** | 4 | 472 |
| KPS* | *N.A.* | *N.A.* | *N.A.* | *N.A.* | *N.A.* | *N.A.* | *N.A.* | *N.A.* | **0.6055** | **0.004** | 1 | 75 |
| ltdi** | **0.5608** | **<0.001** | 3 | 863 | **0.5829** | **<0.001** | 3 | 367 | **0.5492** | **0.002** | 6 | 496 |
| tdi | **0.5549** | **<0.001** | 4 | 863 | 0.5313 | 0.150 | 5 | 367 | **0.5677** | **<0.001** | 5 | 496 |
| volume | 0.5218 | 0.081 | 6 | 863 | 0.5538 | 0.017 | 4 | 36 | 0.5048 | 0.698 | 7 | 496 |

**Table S2. Concordance Hazard indices.** The C-index of every feature was computer by fitting a univariate Cox model and its corresponding *p*-value. Features were ranked according to the C-index. Each univariate Cox model was fitted using the number of samples ($N$) with available information. The effect of site in the joint cohort was removed by fitting a previous model that included the feature of interest and the site. This allowed us to estimate the effect of the site in a feature-specific manner. The significance was assessed by randomly permuting the time of an event and censoring 1000 times. Bold numbers indicate significant C-indices ($p < 0.05$). The asterisks mark whether a given feature appears 1, 2, or 3 times in the top 3 ranked features ('*', '**', and '***' respectively). This table corresponds to Figure 4 (a-b) in the main text. *N.A.* Stands for 'not applicable' due to missing information.

**Table S3 (below). Multivariate Cox survival models.** Hazard ratios ($e^{\beta_i}$), the corresponding *p*-values (two-sided Wald's t-test; $df = N - \#covariates$), and 95% confidence intervals. Bold numbers indicate significant hazard ratios ($p < 0.05$). All the models were fitted after discarding missing values. For the joint cohort ($N = 617$), we corrected for site effects. The asterisks mark whether the $i$-th feature statistically contributes to 0, at least 1, or all the models ('n.s.', '*', and '**' respectively). *N.A.* stands for 'not applicable' due to the specifics of the Cox model fitted (see Methods). The log-likelihood ratio tests are shown in Fig. S12.

### **Joint** ($N = 617$)

| | **Hazard Ratio ($e^{\beta_i}$)** | | | | ***p*-value** | | | | **95% confidence interval** | | | |
|---|---|---|---|---|---|---|---|---|---|---|---|---|
| *Model* | *L-TDI* | *TDI* | *TDI+Volume* | *Naive* | *L-TDI* | *TDI* | *TDI+Volume* | *Naive* | *L-TDI* | *TDI* | *TDI+Volume* | *Naive* |
| sex [n.s.] | 0.9501 | 0.9614 | 0.9570 | 0.9770 | 0.595 | 0.682 | 0.648 | 0.8084 | [0.7865,1.1477] | [0.7960,1.1611] | [0.7925,1.1558] | [0.8095,1.1791] |
| age [**] | **1.0276** | **1.0271** | **1.0273** | **1.0281** | **<0.001** | **<0.001** | **<0.001** | **<0.001** | **[1.0187,1.0367]** | **[1.0182,1.0361]** | **[1.0183,1.0363]** | **[1.0192,1.0371]** |
| mgmt [**] | **0.7270** | **0.7224** | **0.7241** | **0.7262** | **<0.001** | **<0.001** | **<0.001** | **<0.001** | **[0.6603,0.8004]** | **[0.6559,0.7957]** | **[0.6575,0.7974]** | **[0.6594,0.7996]** |
| EOR [**] | **0.5709** | **0.5644** | **0.5652** | **0.5472** | **<0.001** | **<0.001** | **<0.001** | **<0.001** | **[0.4843,0.6730]** | **[0.4792,0.6648]** | **[0.4797,0.6660]** | **[0.4655,0.6433]** |
| ltdi [**] | **1.0742** | *N.A.* | *N.A.* | *N.A.* | **0.005** | *N.A.* | *N.A.* | *N.A.* | **[1.0216,1.1295]** | *N.A.* | *N.A.* | *N.A.* |
| tdi [**] | *N.A.* | **1.0026** | **1.0025** | *N.A.* | *N.A.* | **0.025** | **0.039** | *N.A.* | *N.A.* | **[1.0003,1.0049]** | **[1.0001,1.0048]** | *N.A.* |
| volume [n.s.] | *N.A.* | *N.A.* | 1.0009 | *N.A.* | *N.A.* | *N.A.* | 0.188 | *N.A.* | *N.A.* | *N.A.* | [0.9996,1.0022] | *N.A.* |

### **UCSF** ($N = 355$)

| | **Hazard Ratio ($e^{\beta_i}$)** | | | | ***p*-value** | | | | **95% confidence interval** | | | |
|---|---|---|---|---|---|---|---|---|---|---|---|---|
| *Model* | *L-TDI* | *TDI* | *TDI+Volume* | *Naive* | *L-TDI* | *TDI* | *TDI+Volume* | *Naive* | *L-TDI* | *TDI* | *TDI+Volume* | *Naive* |
| sex [n.s.] | 0.7630 | 0.7768 | 0.7799 | 0.7843 | 0.063 | 0.0883 | 0.093 | 0.092 | [0.5740,1.0144] | [0.5810,1.0386] | [0.5836,1.0422] | [0.5912,1.0405] |
| age [**] | **1.0355** | **1.0362** | **1.0361** | **1.0364** | **<0.001** | **<0.001** | **<0.001** | **<0.001** | **[1.0224,1.0488]** | **[1.0231,1.0495]** | **[1.0230,1.0494]** | **[1.0233,1.0497]** |
| mgmt | 0.8806 | 0.8804 | 0.8804 | 0.8820 | 0.092 | 0.0935 | 0.093 | 0.097 | [0.7595,1.0211] | [0.7586,1.0217] | [0.7589,1.0213] | [0.7605,1.0230] |
| EOR [**] | **0.4497** | **0.4373** | **0.4378** | **0.4343** | **<0.001** | **<0.001** | **<0.001** | **<0.001** | **[0.3616,0.5593]** | **[0.3517,0.5436]** | **[0.3517,0.5450]** | **[0.3510,0.5373]** |
| ltdi [n.s.] | 1.0608 | *N.A.* | *N.A.* | *N.A.* | 0.112 | *N.A.* | *N.A.* | *N.A.* | **[0.9863,1.1409]** | *N.A.* | *N.A.* | *N.A.* |
| tdi [n.s.] | *N.A.* | 1.0005 | 1.0002 | *N.A.* | *N.A.* | 0.7733 | 0.910 | *N.A.* | *N.A.* | [0.9969,1.0042] | [0.9965,1.0040] | *N.A.* |
| volume [n.s.] | *N.A.* | *N.A.* | 1.0012 | *N.A.* | *N.A.* | *N.A.* | 0.1889 | *N.A.* | *N.A.* | *N.A.* | [0.9994,1.0031] | *N.A.* |

### **UPENN** ($N = 262$)

| | **Hazard Ratio ($e^{\beta_i}$)** | | | | ***p*-value** | | | | **95% confidence interval** | | | |
|---|---|---|---|---|---|---|---|---|---|---|---|---|
| *Model* | *L-TDI* | *TDI* | *TDI+Volume* | *Naive* | *L-TDI* | *TDI* | *TDI+Volume* | *Naive* | *L-TDI* | *TDI* | *TDI+Volume* | *Naive* |
| sex [n.s.] | 1.0503 | 1.0851 | 1.0831 | 1.0629 | 0.707 | 0.534 | 0.543 | 0.641 | [0.8131,1.3567] | [0.8387,1.4039] | [0.8372,1.4013] | [0.8224,1.3737] |
| age [**] | **1.0274** | **1.0259** | **1.0260** | **1.0275** | **<0.001** | **<0.001** | **<0.001** | **<0.001** | **[1.0146,1.0403]** | **[1.0131,1.0388]** | **[1.0132,1.0389]** | **[1.0148,1.0403]** |
| mgmt [**] | **0.6266** | **0.6266** | **0.6269** | **0.6268** | **<0.001** | **<0.001** | **<0.001** | **<0.001** | **[0.5439,0.7220]** | **[0.5438,0.7220]** | **[0.5441,0.7223]** | **[0.5437,0.7226]** |
| EOR [**] | **0.7058** | **0.7056** | **0.7062** | **0.6731** | **0.008** | **0.008** | **0.009** | **0.003** | **[0.5451,0.9138]** | **[0.5444,0.9144]** | **[0.5451,0.9149]** | **[0.5208,0.8699]** |
| ltdi [**] | **1.0828** | *N.A.* | *N.A.* | *N.A.* | **0.028** | *N.A.* | *N.A.* | *N.A.* | **[1.0089,1.1622]** | *N.A.* | *N.A.* | *N.A.* |
| tdi [**] | *N.A.* | **1.0041** | **1.0040** | *N.A.* | *N.A.* | **0.011** | **0.013** | *N.A.* | *N.A.* | **[1.0009,1.0072]** | **[1.0009,1.0072]** | *N.A.* |
| volume [n.s.] | *N.A.* | *N.A.* | 1.0004 | *N.A.* | *N.A.* | *N.A.* | 0.673 | *N.A.* | *N.A.* | *N.A.* | [0.9985,1.0024] | *N.A.* |

**UCSF** ($N = 355$)

| | **Hazard Ratio ($e^{\beta_i}$)** | | | | ***p*-value** | | | | **95% confidence interval** | | | |
|---|---|---|---|---|---|---|---|---|---|---|---|---|
| *Model* | *L-TDI* | *TDI* | *TDI+Volume* | *Naive* | *L-TDI* | *TDI* | *TDI+Volume* | *Naive* | *L-TDI* | *TDI* | *TDI+Volume* | *Naive* |
| sex n.s. | 0.7997 | 0.7985 | 0.8022 | 0.8365 | 0.120 | 0.124 | 0.131 | 0.212 | [0.6035,1.0597] | [0.5994,1.0639] | [0.6026,1.0679] | [0.6321,1.1070] |
| age ** | **1.0316** | **1.0315** | **1.0318** | **1.0327** | **<0.001** | **<0.001** | **<0.001** | **<0.001** | **[1.0185,1.0449]** | **[1.0184,1.0447]** | **[1.0187,1.0451]** | **[1.0195,1.0459]** |
| mgmt * | 0.8629 | 0.8634 | **0.8614** | 0.8679 | 0.050 | 0.053 | **0.048** | 0.061 | **[0.7444,1.0003]** | [0.7442,1.0016] | **[0.7428,0.9990]** | [0.7484,1.0065] |
| ltdi ** | **1.1177** | *N.A.* | *N.A.* | *N.A.* | **0.003** | *N.A.* | *N.A.* | *N.A.* | **[1.0389,1.2024]** | *N.A.* | *N.A.* | *N.A.* |
| tdi n.s. | *N.A.* | 1.0028 | 1.0025 | *N.A.* | *N.A.* | 0.113 | 0.162 | *N.A.* | *N.A.* | [0.9993,1.0062] | [0.9990,1.0061] | *N.A.* |
| volume n.s. | *N.A.* | *N.A.* | 1.0015 | *N.A.* | *N.A.* | *N.A.* | 0.117 | *N.A.* | *N.A.* | *N.A.* | [0.9996,1.0034] | *N.A.* |

**Table S4. Multivariate Cox survival models without including the extent of resection (EOR-less).** Hazard ratios ($e^{\beta_i}$), the corresponding $p$-values (two-sided Wald's t-test; $df = N - \#covariates$), and 95% confidence intervals for the UCSF cohort ($N = 355$). Bold numbers indicate significant hazard ratios ($p < 0.05$). All the models were fitted after discarding missing values. The asterisks mark whether the $i$-th feature statistically contributes to 0, at least 1, or all the models ('n.s.', '*', and '**' respectively). *N.A.* stands for 'not applicable' due to the specifics of the Cox model fitted (see Methods). The log-likelihood ratio tests are shown in Fig. S13a.

**UCSF** ($N = 318$)

| | **Hazard Ratio ($e^{\beta_i}$)** | | | | ***p*-value** | | | | **95% confidence interval** | | | |
|---|---|---|---|---|---|---|---|---|---|---|---|---|
| *Model* | *L-TDI* | *TDI* | *TDI+Volume* | *Naive* | *L-TDI* | *TDI* | *TDI+Volume* | *Naive* | *L-TDI* | *TDI* | *TDI+Volume* | *Naive* |
| sex n.s. | 0.8426 | 0.8494 | 0.8531 | 0.8617 | 0.267 | 0.300 | 0.311 | 0.333 | [0.6227,1.1402] | [0.6240,1.1562] | [0.6273,1.1602] | [0.6375,1.1647] |
| age ** | **1.0340** | **1.0344** | **1.0344** | **1.0347** | **<0.001** | **<0.001** | **<0.001** | **<0.001** | **[1.0197,1.0485]** | **[1.0201,1.0490]** | **[1.0201,1.0490]** | **[1.0204,1.0492]** |
| mgmt n.s. | 0.8780 | 0.8802 | 0.8781 | 0.8833 | 0.112 | 0.121 | 0.113 | 0.130 | [0.7479,1.0306] | [0.7491,1.0343] | [0.7476,1.0314] | [0.7524,1.0371] |
| EOR ** | **0.5283** | **0.5108** | **0.5204** | **0.5072** | **<0.001** | **<0.001** | **<0.001** | **<0.001** | **[0.3889,0.7178]** | **[0.3774,0.6914]** | **[0.3839,0.7054]** | **[0.3754,0.6852]** |
| ltdi n.s. | 1.0606 | *N.A.* | *N.A.* | *N.A.* | 0.153 | *N.A.* | *N.A.* | *N.A.* | [0.9784,1.1497] | *N.A.* | *N.A.* | *N.A.* |
| tdi n.s. | *N.A.* | 1.0009 | 1.0006 | *N.A.* | *N.A.* | 0.665 | 0.783 | *N.A.* | *N.A.* | [0.9970,1.0048] | [0.9965,1.0046] | *N.A.* |
| volume n.s. | *N.A.* | *N.A.* | 1.0013 | *N.A.* | *N.A.* | *N.A.* | 0.214 | *N.A.* | *N.A.* | *N.A.* | [0.9993,1.0033] | *N.A.* |

**Table S5. Multivariate Cox survival models after excluding subjects who did not undergo proper resection (only biopsy) in the UCSF cohort.** Hazard ratios ($e^{\beta_i}$), the corresponding $p$-values (two-sided Wald's t-test; $df = N - \#covariates$), and 95% confidence intervals for the UCSF cohort ($N = 318$). Bold numbers indicate significant hazard ratios ($p < 0.05$). All the models were fitted after discarding missing values. The asterisks mark whether the $i$-th feature statistically contributes to 0, at least 1, or all the models ('n.s.', '*', and '**' respectively). *N.A.* stands for 'not applicable' due to the specifics of the Cox model fitted (see Methods). The log-likelihood ratio tests are shown in Fig. S13b.

### Joint ($N = 580$)

| | **Hazard Ratio ($e^{\beta_i}$)** | | | | ***p*-value** | | | | **95% confidence interval** | | | |
|---|---|---|---|---|---|---|---|---|---|---|---|---|
| *Model* | *L-TDI* | *TDI* | *TDI+Volume* | *Naive* | *L-TDI* | *TDI* | *TDI+Volume* | *Naive* | *L-TDI* | *TDI* | *TDI+Volume* | *Naive* |
| sex $^{\text{n.s.}}$ | 0.9801 | 0.9898 | 0.9852 | 1.0054 | 0.839 | 0.918 | 0.881 | 0.957 | [0.8066,1.1908] | [0.8146,1.2027] | [0.8109,1.1970] | [0.8280,1.2209] |
| age $^{**}$ | **1.0286** | **1.0277** | **1.0280** | **1.0288** | **<0.001** | **<0.001** | **<0.001** | **<0.001** | **[1.0193,1.0380]** | **[1.0184,1.0371]** | **[1.0187,1.0375]** | **[1.0195,1.0382]** |
| mgmt $^{**}$ | **0.7056** | **0.7040** | **0.7036** | **0.7083** | **<0.001** | **<0.001** | **<0.001** | **<0.001** | **[0.6389,0.7793]** | **[0.6372,0.7778]** | **[0.6369,0.7772]** | **[0.6413,0.7824]** |
| EOR $^{**}$ | **0.6367** | **0.6291** | **0.6311** | **0.6112** | **<0.001** | **<0.001** | **<0.001** | **<0.001** | **[0.5250,0.7723]** | **[0.5190,0.7625]** | **[0.5207,0.7649]** | **[0.5050,0.7399]** |
| ltdi $^{**}$ | **1.0743** | *N.A.* | *N.A.* | *N.A.* | **0.008** | *N.A.* | *N.A.* | *N.A.* | **[1.0192,1.1324]** | *N.A.* | *N.A.* | *N.A.* |
| tdi $^{**}$ | *N.A.* | **1.0030** | **1.0029** | *N.A.* | *N.A.* | **0.014** | **0.021** | *N.A.* | *N.A.* | **[1.0006,1.0054]** | **[1.0004,1.0053]** | *N.A.* |
| volume $^{\text{n.s.}}$ | *N.A.* | *N.A.* | 1.0008 | *N.A.* | *N.A.* | *N.A.* | 0.252 | *N.A.* | *N.A.* | *N.A.* | [0.9994,1.0022] | *N.A.* |

**Table S6. Multivariate Cox survival models after excluding subjects who did not undergo proper resection (only biopsy) in the joint cohort.** Hazard ratios ($e^{\beta_i}$), the corresponding *p*-values (two-sided Wald's t-test; $df = N - \#covariates$), and 95% confidence intervals for the joint cohort ($N = 580$). Bold numbers indicate significant hazard ratios ($p < 0.05$). The effect of site was removed by fitting a previous model that included the features of interest and the site. All the models were fitted after discarding missing values. The asterisks mark whether the $i$-th feature statistically contributes to 0, at least 1, or all the models ('n.s.', '*', and '**' respectively). *N.A.* stands for 'not applicable' due to the specifics of the Cox model fitted (see Methods). The log-likelihood ratio tests are shown in Fig. S13c.

### UPENN ($N = 53$)

| | **Hazard Ratio ($e^{\beta_i}$)** | | | | ***p*-value** | | | | **95% confidence interval** | | | |
|---|---|---|---|---|---|---|---|---|---|---|---|---|
| *Model* | *L-TDI* | *TDI* | *TDI+Volume* | *Naive* | *L-TDI* | *TDI* | *TDI+Volume* | *Naive* | *L-TDI* | *TDI* | *TDI+Volume* | *Naive* |
| sex $^{\text{n.s.}}$ | 1.1372 | 1.2799 | 1.2430 | 1.2394 | 0.676 | 0.431 | 0.487 | 0.487 | [0.6222,2.0784] | [0.6929,2.3641] | [0.6734, 2.2944] | [0.6766,2.2704] |
| age $^{**}$ | **1.0624** | **1.0622** | **1.0602** | **1.0688** | **<0.001** | **<0.001** | **<0.001** | **<0.001** | **[1.0313,1.0944]** | **[1.0298,1.0956]** | **[1.0280,1.0933]** | **[1.0373,1.1012]** |
| mgmt $^{**}$ | **0.3503** | **0.4034** | **0.3906** | **0.3990** | **<0.001** | **<0.001** | **<0.001** | **<0.001** | **[0.2291,0.5357]** | **[0.2712,0.6000]** | **[0.2606,0.5854]** | **[0.2669,0.5964]** |
| EOR $^{**}$ | 0.5396 | 0.5375 | 0.5295 | 0.6351 | 0.077 | 0.074 | 0.069 | 0.166 | [0.2725,1.0682] | [0.2720,1.0619] | [0.2669,1.0507] | [0.3339,1.2078] |
| KPS $^{*}$ | 0.9772 | **0.9670** | **0.9722** | **0.9639** | 0.059 | **0.004** | **0.027** | **0.002** | [0.9540,1.0009] | **[0.9452,0.9894]** | **[0.9483,0.9968]** | **[0.9423,0.9861]** |
| ltdi $^{**}$ | **1.3519** | *N.A.* | *N.A.* | *N.A.* | **0.005** | *N.A.* | *N.A.* | *N.A.* | **[1.0937,1.6710]** | *N.A.* | *N.A.* | *N.A.* |
| tdi | *N.A.* | 1.0060 | 1.0064 | *N.A.* | *N.A.* | 0.076 | 0.066 | *N.A.* | *N.A.* | [0.9994,1.0126] | [0.9996, 1.0133] | *N.A.* |
| volume $^{\text{n.s.}}$ | *N.A.* | *N.A.* | 1.0033 | *N.A.* | *N.A.* | *N.A.* | 0.323 | *N.A.* | *N.A.* | *N.A.* | [0.9968, 1.0098] | *N.A.* |

**Table S7. Multivariate Cox survival models after excluding subjects with available Karnofsky performance status (KPS) score in the UPENN cohort.** Hazard ratios ($e^{\beta_i}$), the corresponding *p*-values (two-sided Wald's t-test; $df = N - \#covariates$), and 95% confidence intervals for the joint cohort ($N = 53$). Bold numbers indicate significant hazard ratios ($p < 0.05$). All the models were fitted after discarding missing values. The asterisks mark whether the $i$-th feature statistically contributes to 0, at least 1, or all the models ('n.s.', '*', and '**' respectively). *N.A.* stands for 'not applicable' due to the specifics of the Cox model fitted (see Methods). The log-likelihood ratio tests are shown in Fig. S13d.

| **JOINT** | **Coefficient at 6 months $N = 584$** | | | | ***p*-value** | | | | **95% confidence interval** | | | |
|---|---|---|---|---|---|---|---|---|---|---|---|---|
| *Model* | *L-TDI* | *TDI* | *TDI+Volume* | *Naive* | *L-TDI* | *TDI* | *TDI+Volume* | *Naive* | *L-TDI* | *TDI* | *TDI+Volume* | *Naive* |
| constant $^{**}$ | **-3.5769** | **-3.4993** | **-3.6954** | **-3.0058** | **<0.001** | **<0.001** | **<0.001** | **<0.001** | **[-5.397,-1.757]** | **[-5.346,-1.652]** | **[-5.602,-1.788]** | **[-4.727,-1.284]** |
| sex $^{n.s.}$ | -0.2107 | -0.2082 | -0.2121 | -0.1636 | 0.373 | 0.379 | 0.371 | 0.484 | [-0.674,0.253] | [-0.672,0.256] | [-0.677,0.252] | [-0.622,0.294] |
| age $^{**}$ | **0.0675** | **0.0671** | **0.0670** | **0.0684** | **<0.001** | **<0.001** | **<0.001** | **<0.001** | **[0.046,0.089]** | **[0.045,0.089]** | **[0.045,0.089]** | **[0.047,0.090]** |
| mgmt $^{n.s.}$ | -0.1995 | -0.1985 | -0.1967 | -0.2095 | 0.087 | 0.089 | 0.092 | 0.071 | [-0.428,0.029] | [-0.427,0.030] | [-0.426,0.032] | [-0.437,0.018] |
| EOR $^{**}$ | **-0.8248** | **-0.8468** | **-0.8397** | **-0.8929** | **<0.001** | **<0.001** | **<0.001** | **<0.001** | **[-1.180,-0.470]** | **[-1.200,-0.493]** | **[-1.194,-0.486]** | **[-1.241,-0.545]** |
| ltdi $^{**}$ | **0.1335** | *N.A.* | *N.A.* | *N.A.* | **0.037** | *N.A.* | *N.A.* | *N.A.* | **[0.008,0.259]** | *N.A.* | *N.A.* | *N.A.* |
| tdi $^{n.s.}$ | *N.A.* | 0.0043 | 0.0044 | *N.A.* | *N.A.* | 0.133 | 0.130 | *N.A.* | *N.A.* | [-0.001,0.010] | [-0.001,0.010] | *N.A.* |
| volume $^{n.s.}$ | *N.A.* | *N.A.* | 0.0015 | *N.A.* | *N.A.* | *N.A.* | 0.363 | *N.A.* | *N.A.* | *N.A.* | [-0.002,0.005] | *N.A.* |
| | **Coefficient at 12 months $N = 546$** | | | | ***p*-value** | | | | **95% confidence interval** | | | |
| *Model* | *L-TDI* | *TDI* | *TDI+Volume* | *Naive* | *L-TDI* | *TDI* | *TDI+Volume* | *Naive* | *L-TDI* | *TDI* | *TDI+Volume* | *Naive* |
| constant $^{*}$ | **-1.5277** | -1.1045 | **-1.6310** | -0.6189 | **0.048** | 0.156 | **0.043** | 0.392 | **[-3.043,-0.013]** | [-2.630,0.421] | **[-3.214,-0.048]** | [-2.035,0.797] |
| sex $^{n.s.}$ | -0.1230 | -0.1225 | -0.1088 | -0.0948 | 0.550 | 0.546 | 0.597 | 0.638 | [-0.526,0.820] | [-0.520,0.275] | [-0.512,0.294] | [-0.489,0.300] |
| age $^{**}$ | **0.0546** | **0.0545** | **0.0548** | **0.0563** | **<0.001** | **<0.001** | **<0.001** | **<0.001** | **[0.037,0.072]** | **[0.037,0.072]** | **[0.037,0.073]** | **[0.039,0.074]** |
| mgmt $^{**}$ | **-0.2976** | **-0.2905** | **-0.2997** | **-0.2926** | **0.004** | **0.004** | **0.003** | **0.004** | **[-0.498,-0.097]** | **[-0.489,-0.092]** | **[-0.500,-0.099]** | **[-0.490,-0.950]** |
| EOR $^{**}$ | **-0.967** | **-1.0188** | **-1.027** | **-1.0696** | **<0.001** | **<0.001** | **<0.001** | **<0.001** | **[1.302,-0.632]** | **[-1.350,-0.688]** | **[-1.348,-0.677]** | **[-1.397,-0.743]** |
| ltdi $^{**}$ | **0.2185** | *N.A.* | *N.A.* | *N.A.* | **<0.001** | *N.A.* | *N.A.* | *N.A.* | **[0.108,0.329]** | *N.A.* | *N.A.* | *N.A.* |
| tdi $^{n.s.}$ | *N.A.* | 0.0044 | 0.0043 | *N.A.* | *N.A.* | 0.077 | 0.093 | *N.A.* | *N.A.* | [0.000,0.009] | [-0.001,0.009] | *N.A.* |
| volume $^{**}$ | *N.A.* | *N.A.* | **0.0044** | *N.A.* | *N.A.* | *N.A.* | **0.003** | *N.A.* | *N.A.* | *N.A.* | **[0.002,0.007]** | *N.A.* |
| | **Coefficient at 18 months $N = 526$** | | | | ***p*-value** | | | | **95% confidence interval** | | | |
| *Model* | *L-TDI* | *TDI* | *TDI+Volume* | *Naive* | *L-TDI* | *TDI* | *TDI+Volume* | *Naive* | *L-TDI* | *TDI* | *TDI+Volume* | *Naive* |
| constant $^{*}$ | 1.0012 | 1.2224 | 1.0688 | **1.6596** | 0.191 | 0.120 | 0.177 | **0.025** | [-0.501,2.503] | [-0.317,2.762] | [-0.483,2.621] | **[0.209,3.110]** |
| sex $^{n.s.}$ | -0.2025 | -0.2255 | -0.1939 | -0.2020 | 0.314 | 0.262 | 0.336 | 0.312 | [-0.597,0.192] | [-0.620,0.169] | [-0.589,0.201] | [-0.594,0.190] |
| age $^{**}$ | **0.0459** | **0.0455** | **0.0445** | **0.0474** | **<0.001** | **<0.001** | **<0.001** | **<0.001** | **[0.029,0.063]** | **[0.029,0.062]** | **[0.028,0.061]** | **[0.031,0.064]** |
| mgmt $^{**}$ | **-0.4969** | **-0.4672** | **-0.4989** | **-0.4656** | **<0.001** | **<0.001** | **<0.001** | **<0.001** | **[-0.702,-0.292]** | **[-0.671,-0.263]** | **[-0.703,-0.294]** | **[-0.669,-0.263]** |
| EOR $^{**}$ | **-1.0477** | **-1.1001** | **-1.0760** | **-1.1509** | **<0.001** | **<0.001** | **<0.001** | **<0.001** | **[-1.410,-0.685]** | **[-1.466,-0.735]** | **[-1.439,-0.713]** | **[-1.512,-0.790]** |
| ltdi $^{**}$ | **0.1434** | *N.A.* | *N.A.* | *N.A.* | **0.010** | *N.A.* | *N.A.* | *N.A.* | **[0.034,0.253]** | *N.A.* | *N.A.* | *N.A.* |
| tdi $^{n.s.}$ | *N.A.* | 0.0041 | 0.0034 | *N.A.* | *N.A.* | 0.094 | 0.166 | *N.A.* | *N.A.* | [-0.001,0.009] | [-0.001,0.008] | *N.A.* |
| volume $^{n.s.}$ | *N.A.* | *N.A.* | 0.0021 | *N.A.* | *N.A.* | *N.A.* | 0.144 | *N.A.* | *N.A.* | *N.A.* | [-0.001,0.005] | *N.A.* |

*Continues in the next page*

| **UCSF** | **Coefficient at 6 months $N = 322$** | | | | ***p*-value** | | | | **95% confidence interval** | | | |
|---|---|---|---|---|---|---|---|---|---|---|---|---|
| *Model* | *L-TDI* | *TDI* | *TDI+Volume* | *Naive* | *L-TDI* | *TDI* | *TDI+Volume* | *Naive* | *L-TDI* | *TDI* | *TDI+Volume* | *Naive* |
| constant $^{**}$ | **-4.6943** | **-3.6383** | **-4.3492** | **-3.7732** | **<0.001** | **0.005** | **0.002** | **0.002** | **[-7.248,-2.141]** | **[-6.184,-1.092]** | **[-7.051,-1.648]** | **[-6.178,-1.369]** |
| sex $^{**}$ | **-0.8517** | **-0.7724** | **-0.7623** | **-0.7909** | **0.018** | **0.031** | **0.035** | **0.025** | **[-1.558,-0.145]** | **[-1.475,-0.070]** | **[-1.471,-0.053]** | **[-1.485,-0.097]** |
| age $^{**}$ | **0.0811** | **0.0841** | **0.0843** | **0.0834** | **<0.001** | **<0.001** | **<0.001** | **<0.001** | **[0.049,0.113]** | **[0.052,0.116]** | **[0.052,0.117]** | **[0.052,0.115]** |
| mgmt $^{n.s.}$ | 0.0271 | 0.0135 | 0.0109 | 0.0171 | 0.880 | 0.940 | 0.952 | 0.924 | [-0.326,0.380] | [-0.337,0.364] | [-0.342,0.364] | [-0.332,0.367] |
| EOR $^{**}$ | **-0.7761** | **-0.9079** | **-0.8947** | **-0.8914** | **0.001** | **<0.001** | **<0.001** | **<0.001** | **[-1.221,-0.331]** | **[-1.354,-0.462]** | **[-1.348,-0.441]** | **[-1.325,-0.458]** |
| ltdi $^{**}$ | **0.2132** | *N.A.* | *N.A.* | *N.A.* | **0.019** | *N.A.* | *N.A.* | *N.A.* | **[0.035,0.391]** | *N.A.* | *N.A.* | *N.A.* |
| tdi $^{n.s.}$ | *N.A.* | -0.0014 | -0.0016 | *N.A.* | *N.A.* | 0.748 | 0.733 | *N.A.* | *N.A.* | [-0.010,.007] | [-0.011,0.008] | *N.A.* |
| volume $^{**}$ | *N.A.* | *N.A.* | **0.0054** | *N.A.* | *N.A.* | *N.A.* | **0.012** | *N.A.* | *N.A.* | *N.A.* | **[0.001,0.010]** | *N.A.* |
| | **Coefficient at 12 months $N = 284$** | | | | ***p*-value** | | | | **95% confidence interval** | | | |
| *Model* | *L-TDI* | *TDI* | *TDI+Volume* | *Naive* | *L-TDI* | *TDI* | *TDI+Volume* | *Naive* | *L-TDI* | *TDI* | *TDI+Volume* | *Naive* |
| constant $^{n.s.}$ | -2.0003 | -1.1035 | -1.6546 | -1.2035 | 0.051 | 0.288 | 0.124 | 0.210 | [-4.011,0.010] | [-3.138,0.931] | [-3.765,0.456] | [-3.084,0.677] |
| sex $^{n.s.}$ | -0.4449 | -0.4402 | -0.4135 | -0.4456 | 0.130 | 0.129 | 0.161 | 0.124 | [-1.021,0.131] | [-1.009,0.128] | [-0.992,0.165] | [-1.013,0.121] |
| age $^{**}$ | **0.0667** | **0.0696** | **0.0691** | **0.0691** | **<0.001** | **<0.001** | **<0.001** | **<0.001** | **[0.041,0.092]** | **[0.044,0.095]** | **[0.043,0.095]** | **[0.044,0.095]** |
| mgmt $^{n.s.}$ | -0.975 | -0.0886 | -0.0983 | -0.0890 | 0.521 | 0.557 | 0.519 | 0.555 | [-0.396,0.201] | [-0.384,0.207] | [-0.397,0.200] | [-0.384,0.206] |
| EOR $^{**}$ | **-0.9888** | **-1.1135** | **-1.0916** | **-1.1006** | **<0.001** | **<0.001** | **<0.001** | **<0.001** | **[-1.428,-0.550]** | **[-1.553,-0.674]** | **[-1.536,-0.648]** | **[-1.528,-0.673]** |
| ltdi $^{**}$ | **0.1817** | *N.A.* | *N.A.* | *N.A.* | **0.022** | *N.A.* | *N.A.* | *N.A.* | **[0.026,0.337]** | *N.A.* | *N.A.* | *N.A.* |
| tdi $^{n.s.}$ | *N.A.* | -0.001 | -0.0014 | *N.A.* | *N.A.* | 0.801 | 0.718 | *N.A.* | *N.A.* | [-0.008,0.007] | [-0.009,0.006] | *N.A.* |
| volume $^{**}$ | *N.A.* | *N.A.* | **0.0047** | *N.A.* | *N.A.* | *N.A.* | **0.014** | *N.A.* | *N.A.* | *N.A.* | **[0.001,0.008]** | *N.A.* |
| | **Coefficient at 18 months $N = 264$** | | | | ***p*-value** | | | | **95% confidence interval** | | | |
| *Model* | *L-TDI* | *TDI* | *TDI+Volume* | *Naive* | *L-TDI* | *TDI* | *TDI+Volume* | *Naive* | *L-TDI* | *TDI* | *TDI+Volume* | *Naive* |
| constant $^{n.s.}$ | 0.6994 | 1.3645 | 1.1705 | 1.1222 | 0.51 | 0.214 | 0.291 | 0.267 | [-1.380,2.779] | [-0.787,3.516] | [-1.003,3.344] | [-0.858,3.102] |
| sex $^{n.s.}$ | -0.3162 | -0.3163 | -0.2966 | -0.3270 | 0.275 | 0.274 | 0.308 | 0.257 | [-0.884,0.252] | [-0.883,0.251] | [-0.867,0.273] | [-0.893,0.239] |
| age $^{**}$ | **0.0576** | **0.0606** | **0.0598** | **0.0594** | **<0.001** | **<0.001** | **<0.001** | **<0.001** | **[0.032,0.083]** | **[0.035,0.0086]** | **[0.034,0.085]** | **[0.034,0.085]** |
| mgmt $^{n.s.}$ | -0.2523 | -0.2444 | -0.2516 | -0.2453 | 0.114 | 0.125 | 0.116 | 0.123 | [-0.565,0.061] | [-0.557,0.068] | [-0.565,0.062] | [-0.557,0.067] |
| EOR $^{**}$ | **-1.2301** | **-1.3338** | **-1.3172** | **-1.2996** | **<0.001** | **<0.001** | **<0.001** | **<0.001** | **[-1.750,-0.710]** | **[-1.859,-0.809]** | **[-1.842,-0.793]** | **[-1.810,-0.790]** |
| ltdi $^{n.s.}$ | 0.0091 | *N.A.* | *N.A.* | *N.A.* | 0.218 | *N.A.* | *N.A.* | *N.A.* | [-0.059,0.257] | *N.A.* | *N.A.* | *N.A.* |
| tdi $^{n.s.}$ | *N.A.* | -0.0022 | -0.0026 | *N.A.* | *N.A.* | 0.566 | 0.503 | *N.A.* | *N.A.* | [-0.010,0.005] | [-0.010,0.005] | *N.A.* |
| volume $^{n.s.}$ | *N.A.* | *N.A.* | 0.0020 | *N.A.* | *N.A.* | *N.A.* | 0.315 | *N.A.* | *N.A.* | *N.A.* | [-0.002,0.006] | *N.A.* |

*Continues in the next page*

| **UPENN** | **Coefficient at 6 months $N = 262$** | | | | ***p*-value** | | | | **95% confidence interval** | | | |
|---|---|---|---|---|---|---|---|---|---|---|---|---|
| *Model* | *L-TDI* | *TDI* | *TDI+Volume* | *Naive* | *L-TDI* | *TDI* | *TDI+Volume* | *Naive* | *L-TDI* | *TDI* | *TDI+Volume* | *Naive* |
| constant ** | **-3.0823** | **-3.6198** | **-3.4722** | **-2.7064** | **0.012** | **0.005** | **0.009** | **0.02** | **[-5.487,-0.677]** | **[-6.160,-1.079]** | **[-6.058,-0.886]** | **[-4.991,-0.422]** |
| sex n.s. | 0.3025 | 0.3026 | 0.3139 | 0.3477 | 0.345 | 0.346 | 0.330 | 0.272 | [-0.325,0.930] | [-0.327,0.932] | [-0.317,0.945] | [-0.273,0.968] |
| age ** | **0.0500** | **0.0491** | **0.0493** | **0.0508** | **0.001** | **0.001** | **0.001** | **0.001** | **[0.020,0.080]** | **[0.019,0.079]** | **[0.019,0.079]** | **[0.021,0.080]** |
| mgmt ** | **-0.4345** | **-0.4505** | **-0.4605** | **-0.4499** | **0.012** | **0.010** | **0.009** | **0.01** | **[-0.775,-0.094]** | **[-0.793,-0.108]** | **[-0.806,-0.115]** | **[-0.790,-0.109]** |
| EOR ** | **-0.9301** | **-0.877** | **-0.8834** | **-0.9781** | **0.004** | **0.006** | **0.006** | **0.002** | **[-1.555,-0.306]** | **[-1.508,-0.246]** | **[-1.517,-0.250]** | **[-1.598,-0.358]** |
| ltdi n.s. | 0.108 | *N.A.* | *N.A.* | *N.A.* | 0.213 | *N.A.* | *N.A.* | *N.A.* | [-0.069,0.285] | *N.A.* | *N.A.* | *N.A.* |
| tdi ** | *N.A.* | **0.0077** | **0.0076** | *N.A.* | *N.A.* | **0.046** | **0.046** | *N.A.* | *N.A.* | **[0.000,0.015]** | **[0,.000,0.015]** | *N.A.* |
| volume n.s. | *N.A.* | *N.A.* | -0.0013 | *N.A.* | *N.A.* | *N.A.* | 0.601 | *N.A.* | *N.A.* | *N.A.* | [-0.006,0.004] | *N.A.* |
| | **Coefficient at 12 months $N = 262$** | | | | ***p*-value** | | | | **95% confidence interval** | | | |
| *Model* | *L-TDI* | *TDI* | *TDI+Volume* | *Naive* | *L-TDI* | *TDI* | *TDI+Volume* | *Naive* | *L-TDI* | *TDI* | *TDI+Volume* | *Naive* |
| constant n.s. | -1.2105 | -1.5008 | -1.6561 | -0.6566 | 0.238 | 0.160 | 0.128 | 0.500 | [-3.223,0.801] | [-3.592I0.590] | [-3.791,0.479] | [-2.564,1.250] |
| sex n.s. | 0.0387 | 0.0410 | 0.0393 | 0.0835 | 0.892 | 0.886 | 0.891 | 0.767 | [-0.522,0.600] | [-0.519,0.601] | [-0.522,0.601] | [-0.468,0.635] |
| age ** | **0.0438** | **0.0433** | **0.43** | **0.0462** | **0.001** | **0.001** | **0.001** | **<0.001** | **[0.019,0.069]** | **[0.018,0.068]** | **[0.018,0.068]** | **[0.022,0.070]** |
| mgmt ** | **-0.5902** | **-0.6105** | **-0.6023** | **-0.5986** | **<0.001** | **<0.001** | **<0.001** | **<0.001** | **[0.032,0.340]** | **[-0.913,-0.308]** | **[-0.905,-0.300]** | **[-0.897,-0.300]** |
| EOR ** | **-0.9977** | **-0.9683** | **-0.9712** | **-1.0574** | **<0.001** | **0.001** | **0.001** | **<0.001** | **[-1.550,-0.446]** | **[-1.524,-0.413]** | **[-1.528,-0.415]** | **[-1.602,-0.513]** |
| ltdi ** | **0.1861** | *N.A.* | *N.A.* | *N.A.* | **0.018** | *N.A.* | *N.A.* | *N.A.* | **[0.032,0.340]** | *N.A.* | *N.A.* | *N.A.* |
| tdi ** | *N.A.* | **0.0082** | **0.0081** | *N.A.* | *N.A.* | **0.012** | **0.014** | *N.A.* | *N.A.* | **[0.002,0.015]** | **[0.002,0.015]** | *N.A.* |
| volume n.s. | *N.A.* | *N.A.* | 0.0018 | *N.A.* | *N.A.* | *N.A.* | 0.417 | *N.A.* | *N.A.* | *N.A.* | [-0.002,0.006] | *N.A.* |
| | **Coefficient at 18 months $N = 262$** | | | | ***p*-value** | | | | **95% confidence interval** | | | |
| *Model* | *L-TDI* | *TDI* | *TDI+Volume* | *Naive* | *L-TDI* | *TDI* | *TDI+Volume* | *Naive* | *L-TDI* | *TDI* | *TDI+Volume* | *Naive* |
| constant n.s. | 0.7695 | 0.3929 | 0.3270 | 1.273 | 0.449 | 0.706 | 0.756 | 0.198 | [-1.222,2.761] | [-1.651,2.436] | [-1.740,2.393] | [-0.663,3.209] |
| sex n.s. | -0.1409 | -0.1885 | -0.1811 | -0.1218 | 0.631 | 0.524 | 0.541 | 0.673 | [-0.716,0.434] | [-0.768,0.391] | [-0.762,0.400] | [-0.688,0.444] |
| age ** | **0.0385** | **0.0372** | **0.0371** | **0.0415** | **0.002** | **0.003** | **0.003** | **0.001** | **[0.015,0.062]** | **[0.013,0.061]** | **[0.013,0.061]** | **[0.018,0.065]** |
| mgmt ** | **-0.7865** | **-0.8127** | **-0.8116** | **-0.7732** | **<0.001** | **<0.001** | **<0.001** | **<0.001** | **[-1.103,-0.470]** | **[-1.134,-0.492]** | **[-1.133,-0.490]** | **[-1.084,-0.463]** |
| EOR ** | **-0.9965** | **-0.9599** | **-0.9668** | **-1.0383** | **0.001** | **0.002** | **0.002** | **<0.001** | **[-1.588,-0.405]** | **[-1.557,-0.363]** | **[-1.565,-0.368]** | **[-1.620,-0.456]** |
| ltdi ** | **0.2015** | *N.A.* | *N.A.* | *N.A.* | **0.013** | *N.A.* | *N.A.* | *N.A.* | **[0.042,0.361]** | *N.A.* | *N.A.* | *N.A.* |
| tdi ** | *N.A.* | **0.0105** | **0.0103** | *N.A.* | *N.A.* | **0.001** | **0.002** | *N.A.* | *N.A.* | **[0.004,0.017]** | **[0.004,0.017]** | *N.A.* |
| volume n.s. | *N.A.* | *N.A.* | 0.001 | *N.A.* | *N.A.* | *N.A.* | 0.658 | *N.A.* | *N.A.* | *N.A.* | [-0.003,0.005] | *N.A.* |

*Caption in the next page*

**Table S8. Multivariate logistic regressions.** The logistic regressions were fitted to predict the probability of dying (status=1) before 6, 12, and 18 months. Patients who were lost to follow-up (status=0) before each time horizon month were discarded. The top part of the table corresponds to the results using the joint cohort; the middle part corresponds to the results using the UCSF cohort; and the bottom part corresponds to the results using the UPENN cohort. For the joint cohort, regressions were fit after correcting for site effects. Samples with missing values were discarded in all cases. The coefficients and the corresponding *p*-values (two-sided Wald's z-test; $df = N - \#covariates$). Bold numbers indicate significant hazard ratios ($p < 0.05$). The asterisks mark whether the $i$-th feature statistically contributes to 0, at least 1, or all the models ('n.s.', '*', and '**' respectively). *N.A.* stands for 'not applicable' due to the specifics of the logistic regressions fitted (see Methods). The log-likelihood ratio tests are shown in Fig. 4f and Fig. S15.

| Table S9a | | **TRAIN (6 months)** | | | | | | | | | | | |
|---|---|---|---|---|---|---|---|---|---|---|---|---|---|
| | | UCSF | | | UPENN | | | UCSF | | | UPENN | | |
| *Model* | | *L-TDI* | *TDI* | *Naive* | *L-TDI* | *TDI* | *Naïve* | *L-TDI* | *TDI* | *Naive* | *L-TDI* | *TDI* | *Naive* |
| **TEST (6 months)** | UCSF | *N.A.* | | | 6 | 2 | 6 | *N.A.* | | | 5 | 5 | 5 |
| | UPENN | 6 | 6 | 8 | *N.A.* | | | 7 | 6 | 9 | *N.A.* | | |
| | | *Area under the curve (AUC)* | | | | | | *Balanced accuracy (bACC)* | | | | | |

| Table S9b | | **TRAIN (12 months)** | | | | | | | | | | | |
|---|---|---|---|---|---|---|---|---|---|---|---|---|---|
| | | UCSF | | | UPENN | | | UCSF | | | UPENN | | |
| *Model* | | *L-TDI* | *TDI* | *Naive* | *L-TDI* | *TDI* | *Naïve* | *L-TDI* | *TDI* | *Naive* | *L-TDI* | *TDI* | *Naive* |
| **TEST (6 months)** | UCSF | *N.A.* | | | 2 | 3 | 10 | *N.A.* | | | 12 | 11 | 23 |
| | UPENN | 5 | 9 | 3 | *N.A.* | | | 5 | 12 | 11 | *N.A.* | | |
| | | *Area under the curve (AUC)* | | | | | | *Balanced accuracy (bACC)* | | | | | |

| Table S9c | | **TRAIN (18 months)** | | | | | | | | | | | |
|---|---|---|---|---|---|---|---|---|---|---|---|---|---|
| | | UCSF | | | UPENN | | | UCSF | | | UPENN | | |
| *Model* | | *L-TDI* | *TDI* | *Naive* | *L-TDI* | *TDI* | *Naïve* | *L-TDI* | *TDI* | *Naive* | *L-TDI* | *TDI* | *Naive* |
| **TEST (6 months)** | UCSF | *N.A.* | | | 7 | 3 | 2 | *N.A.* | | | 6 | 5 | 5 |
| | UPENN | 7 | 4 | 4 | *N.A.* | | | 8 | 5 | 14 | *N.A.* | | |
| | | *Area under the curve (AUC)* | | | | | | *Balanced accuracy (bACC)* | | | | | |

**Table S9. Training specifics for the logistic classifiers trained to predict death.** The specifics used to train the different models with the two independent cohorts. The exact values of these parameters were not selected *ad hoc* and were the results of automated and unbiased procedures solely aimed at maximizing the performance of the final model on the corresponding test cohort. See the methods in the main text for the explicit procedures.

| Table S8a | | TRAIN (6 months) | | | | | | | | | | | |
|---|---|---|---|---|---|---|---|---|---|---|---|---|---|
| | | UCSF | | | UPENN | | | UCSF | | | UPENN | | |
| *Model* | | *L-TDI* | *TDI* | *Naive* | *L-TDI* | *TDI* | *Naïve* | *L-TDI* | *TDI* | *Naive* | *L-TDI* | *TDI* | *Naive* |
| **TEST (6 months)** | UCSF | *N.A.* | | | **0.7376** | 0.7237 | 0.7305 | *N.A.* | | | **0.5902** | 0.5357 | 0.5864 |
| | UPENN | **0.7090** | 0.6970 | 0.7018 | *N.A.* | | | 0.5099 | **0.5444** | **0.5444** | *N.A.* | | |

*Area under the curve (AUC)* — *Balanced accuracy (bACC)*

| Table S8b | | TRAIN (12 months) | | | | | | | | | | | |
|---|---|---|---|---|---|---|---|---|---|---|---|---|---|
| | | UCSF | | | UPENN | | | UCSF | | | UPENN | | |
| *Model* | | *L-TDI* | *TDI* | *Naive* | *L-TDI* | *TDI* | *Naïve* | *L-TDI* | *TDI* | *Naive* | *L-TDI* | *TDI* | *Naive* |
| **TEST (6 months)** | UCSF | *N.A.* | | | **0.7433** | 0.7088 | 0.7220 | *N.A.* | | | **0.6498** | 0.6463 | 0.6418 |
| | UPENN | **0.7327** | 0.7039 | 0.7077 | *N.A.* | | | **0.6753** | 0.656 | 0.6661 | *N.A.* | | |

*Area under the curve (AUC)* — *Balanced accuracy (bACC)*

| Table S8c | | TRAIN (18 months) | | | | | | | | | | | |
|---|---|---|---|---|---|---|---|---|---|---|---|---|---|
| | | UCSF | | | UPENN | | | UCSF | | | UPENN | | |
| *Model* | | *L-TDI* | *TDI* | *Naive* | *L-TDI* | *TDI* | *Naïve* | *L-TDI* | *TDI* | *Naive* | *L-TDI* | *TDI* | *Naive* |
| **TEST (6 months)** | UCSF | *N.A.* | | | **0.7387** | 0.7153 | 0.7360 | *N.A.* | | | **0.6799** | 0.6338 | 0.6519 |
| | UPENN | **0.7292** | 0.7090 | 0.7168 | *N.A.* | | | 0.6640 | 0.6475 | **0.6679** | *N.A.* | | |

*Area under the curve (AUC)* — *Balanced accuracy (bACC)*

**Table S10. Classification metrics for the logistic classifiers trained to predict death.** Area under the curve (AUC) and balanced accuracy (bACC) of the models trained and tested on the different combinations of the two independent cohorts. The different time horizons are shown in sub-tables for **(a)** 6, **(b)** 12**,** and **(c)** 18 months. *N.A.* stands for not applicable, since we did not perform testing on the cohort used for training. Bold numbers indicate the best-performing model on the testing cohort.
