## Supplementary Figures for "A non-local diffusion magnetic resonance imaging tract density biomarker to stratify, predict, and interpret survival rates in human glioblastoma"

[1]Computational Neuroscience Group, Sano Centre for Computational Medicine, Kraków, Poland. [2]Brain Mapping Lab, Department of Biomedical, Dental Sciences and Morphological and Functional Imaging, University of Messina, Messina, Italy. [3]Department of Neurophysiology and Chronobiology, Institute of Zoology and Biomedical Research, Faculty of Biology, Jagiellonian University, Krakow, Poland. [4]Radiation Oncology Unit, Clinical Department, National Center for Oncological Hadrontherapy (CNAO), 27100, Pavia, Italy. [5]Department of Internal Medicine and Medical Therapy, University of Pavia, 27100 Pavia, Italy. [6]Department of Biomedical Sciences, Humanitas University, Via Rita Levi Montalcini 4, Pieve Emanuele, 20072 Milan, Italy. [7]IRCCS Humanitas Research Hospital, Via Alessandro Manzoni 56, Rozzano, 20089 Milan, Italy. [8]Faculty of Physics, Astronomy and Applied Computer Science, Jagiellonian University, Krakow, Poland.

[†]Equal contribution: Gianpaolo Antonio Basile, Anna Janus

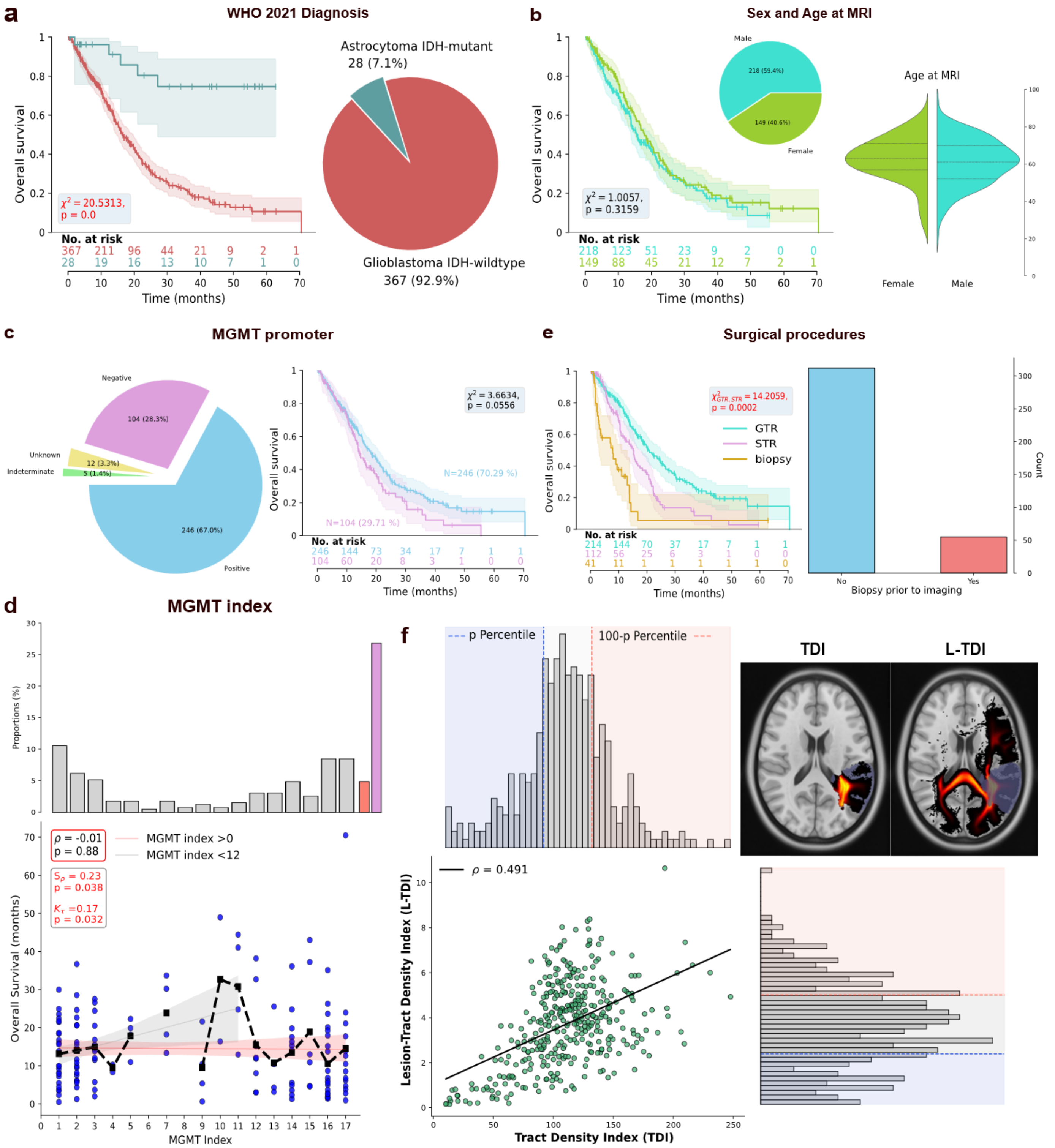

**Figure S1. Description of the UCSF cohort. (a)** Kaplan-Meier estimators including the 95% confidence interval (left) and the percentages of the two groups stratified according to the diagnosis set according to the WHO 2021 guidelines (right). IDH-wildtype glioblastomas (GBM, $n = 367$) carried worse prognosis compared to IDH-mutant astrocytoma ($\chi^2 = 20.513$, $p < 0.001$, two-sided log-rank test). All subsequent analyses were run on the GBM group. **(b)** Survival curves for males (turquoise) and females (light green) depict a slightly worse prognosis for males, although it did not reach statistical significance ($\chi^2 = 1.006$, $p = 0.316$, two-sided log-rank test). Males were slightly more prevalent than females (60/40% ratio; inset) but their mean age did not differ ($U = 14836$, $p = 0.159$, two-sided U-test; $t = -1.248$, $p = 0.213$, $df = 323$, two-sided Welch's t-test; right). **(c)** MGMT promoter distribution (left). Most samples were methylated (cyan), followed by unmethylated (violet), indeterminate (green), and unknown (yellow). Methylation was linked to survival benefits, but they were not significant ($\chi^2 = 3.366$, $p = 0.056$, two-sided log-rank test). **(d)** Distribution of MGMT indices (top): methylated (gray), unavailable (thin red), and unmethylated (thin violet). The black dashed line shows the average survival times as a function of MGMT index (bottom). There was no significant correlation between survival in patients known to be deceased and the MGMT index ($\rho = 0.01$, $p = 0.85$, two-sided exact test), but a strong positive relationship was found for indices below 11 (Spearman $\rho = 0.230$, $p = 0.038$; Kendall $\tau = 0.170$, $p = 0.032$; two-sided permutation tests $n = 5000$). **(e)** Survival curves (left): gross total resection (GTR, turquoise), subtotal resection (STR, violet), and biopsy, respectively (gold). Better survival rates followed GTR as compared to STR ($\chi^2 = 14.206$, $p < 0.001$, two-sided log-rank test). A few patients underwent a biopsy before imaging (right). **(f)** Tract density index (TDI) and lesion-tract density index (L-TDI) showed a nonlinear relationship. Histograms illustrate differing distributions, with L-TDI displaying non-normal behavior.

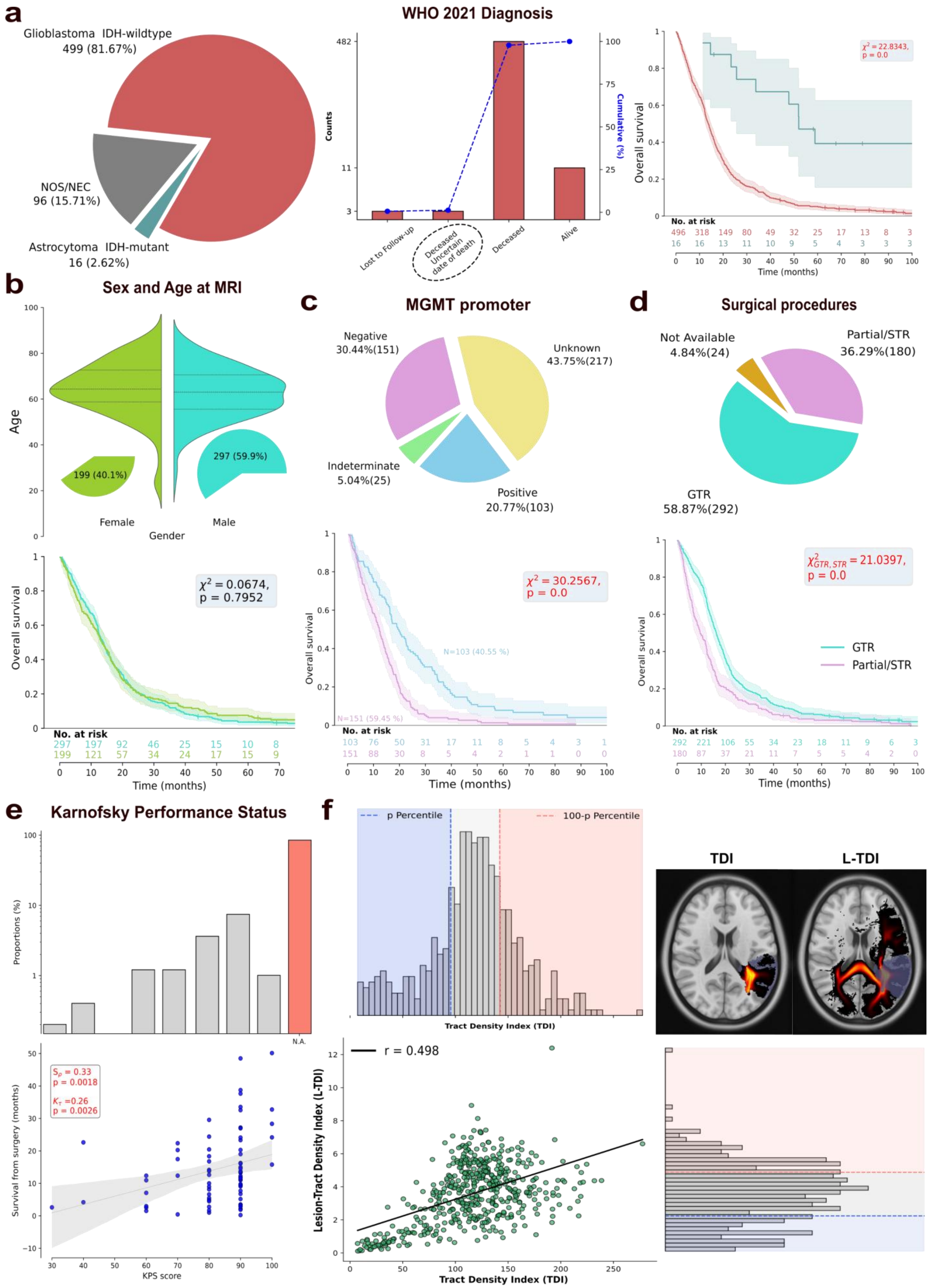

*Caption on the next page*

**Figure S2. Description of the UPENN cohort. (a)** Distribution of patients based on the WHO 2021 guidelines (left), classifying high-grade gliomas based on the mutational status of IDH. The 96 cases (NOS/NEC) were classified according to "The Consortium to Inform Molecular and Practical Approaches to CNS Tumor Taxonomy–Not Official WHO (cIMPACT-NOW)" as IDH-Not-Otherwise-Specified (IDH-NOS). The number of occurrences for each event (middle) in the IDH-wildtype glioblastoma group (GBM). 3 patients did not have available survival data (dotted black circle) and were discarded from the analyses. Kaplan-Meier estimators from the day of surgery, including the 95% confidence interval (right). GBMs ($n = 496$) carried a worse prognosis compared to IDH-mutant astrocytoma ($\chi^2 = 22.834$, $p < 0.001$, two-sided log-rank test). All subsequent analyses were run on the GBM group with recorded survival/censorship times. **(b)** Males were slightly more prevalent than females ($60/40\%$ ratio) and their mean age slightly differed ($U = 26077$, $p = 0.025$, two-sided U-test; $t = -1.768$, $p = 0.079$, $df = 393$, two-sided Welch's t-test; top), albeit rather in an inconclusive manner. Survival curves (bottom) for males and females depict comparable prognosis ($\chi^2 = 0.067$, $p = 0.795$, two-sided log-rank test). **(c)** MGMT promoter distribution (top). Most samples had unknown methylation status (light yellow), followed by unmethylated (violet), methylated (cyan), and indeterminate (green). Methylation was linked to a clear significant survival benefit (bottom; $\chi^2 = 30.2568$, $p < 0.001$, two-sided log-rank test). **(d)** Distribution of surgical procedures (top). Most patients underwent gross total resection (GTR; removal of over $90\%$ of the tumor) and partial tumor resection (STR; removal of less than $90\%$ of the tumor). GTR was associated with increased survival rates ($\chi^2 = 21.257$, $p < 0.001$, two-sided log-rank test). **(e)** Distribution of the Karnofsky performance status (KPS) scores (top). A large percentage of subjects had unavailable data (red bar; $84.88\%$). The y-axis is on a logarithmic scale to enhance visibility. A clear linear trend existed between the KPS scores and survival (Spearman $\rho = 0.330$, $p = 0.002$; Kendall $\tau = 0.260$, $p < 0.003$; two-sided permutation tests $n = 5000$). **(f)** Tract density index (TDI) and lesion-tract density index (L-TDI) showed a nonlinear relationship. Histograms illustrate differing distributions, with L-TDI displaying non-normal behavior.

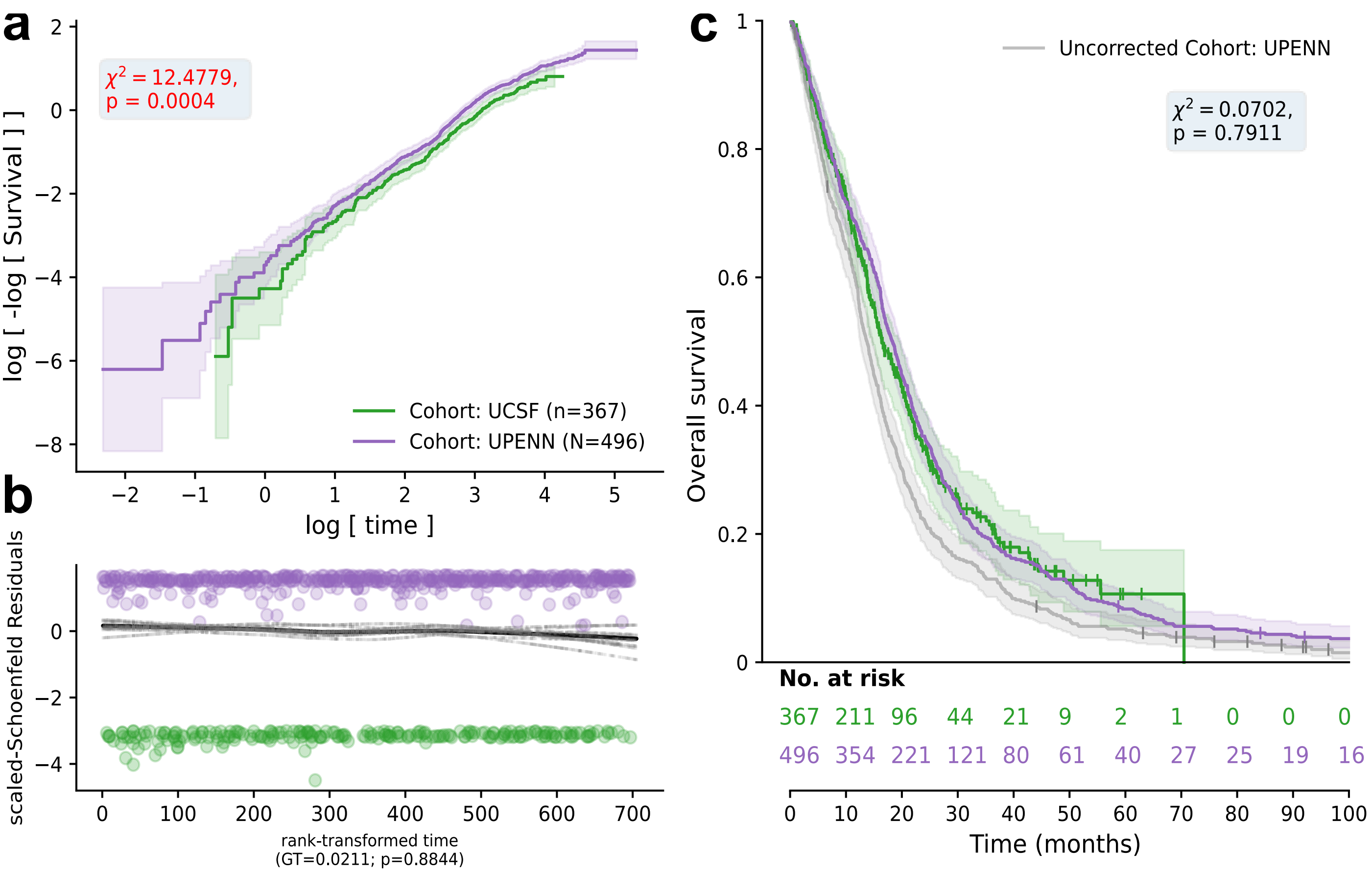

**Figure S3. Unifying the survival times in different cohorts. (a)** Kaplan-Meier log-log plot of the survival time [months] and the overall survival for each cohort. The shaded areas correspond to the 95% confidence interval. Initially, there existed a significant difference in the survival rates ($\chi^2 = 12.4779$, $p = 0.0004$; two-sided log-rank test). Given that both curves are largely parallel, the proportional hazard assumption is likely to hold. **(b)** Scaled-Schoenfeld residuals for each cohort. The absence of any trends confirms the validity of the proportional hazard assumption (GT=0.0211, p=0.884; two-sided Grambsch-Therneau's test). **(c)** Kaplan-Meier curves of corrected survival times compared to the uncorrected ones (gray). The shaded areas correspond to the 95% confidence interval. After transforming the survival times, the differences in survival disappeared ($\chi^2 = 0.0702$, $p = 0.7911$; two-sided log-rank test).

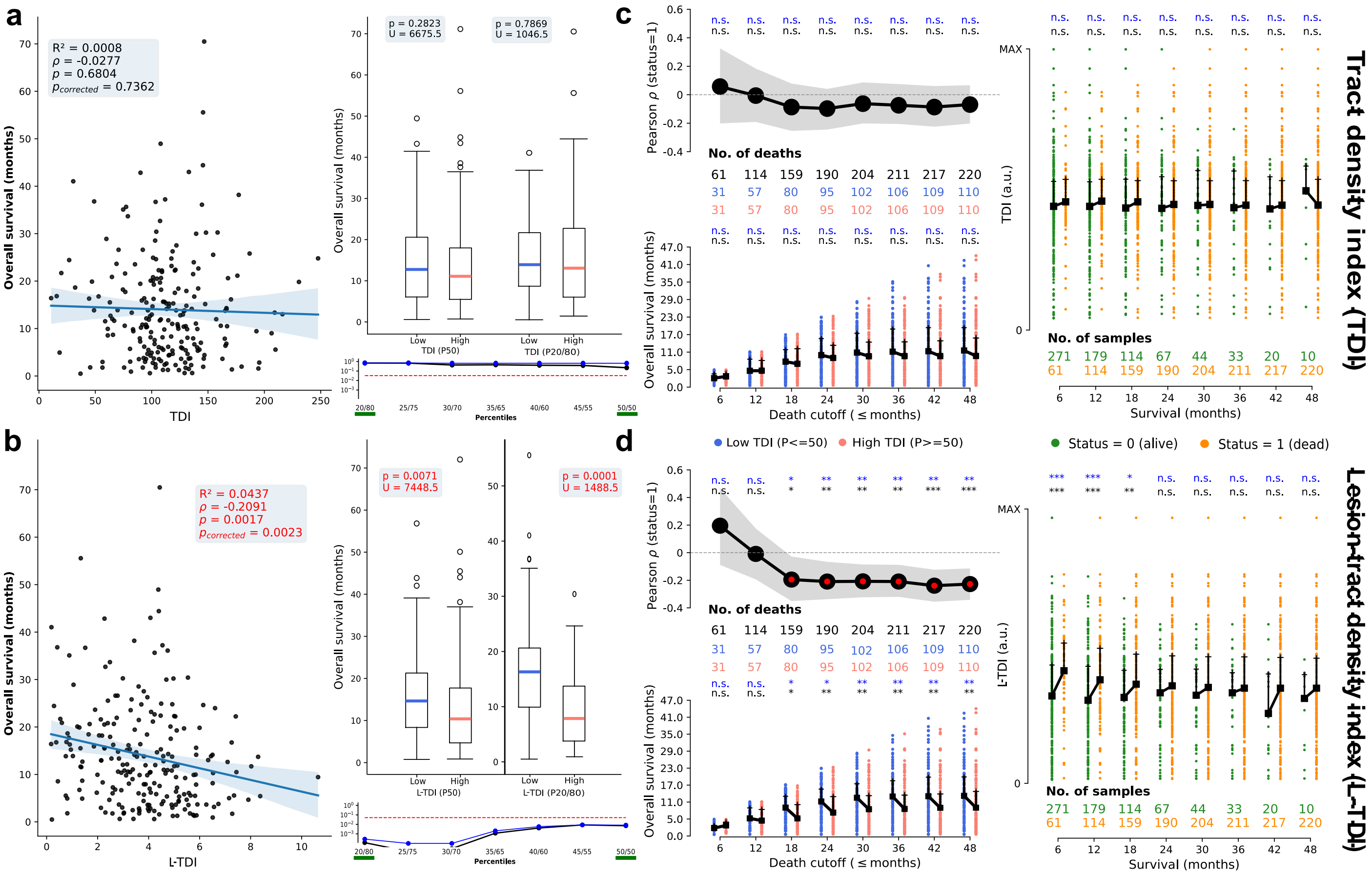

**Figure S4. Association between (lesion-)tract density indices and overall survival in the UCSF cohort. (a)** The correlation (left) between the tract density index (TDI) and the overall survival (OS) in patients known to be deceased was negative but not significant ($\rho = -0.028$, $p = 0.680$, two-sided exact test). Upon splitting these patients into two groups based on the 50$^{th}$ and 20$^{th}$/80$^{th}$ TDI percentiles (right), the median survival times in patients with lower TDIs were higher but without reaching statistical significance ($U = 6675.5$, $p = 0.2823$; $U = 1046.5$, $p = 0.7869$; two-sided U-tests). Boxplots depict the median for the low (blue) and high (salmon) TDI strata, the interquartile range (black box), the 1$^{st}$/4$^{th}$ quartiles (whiskers), and outliers. Differences in median survival times were checked across different stratification thresholds (bottom right), but did not reach statistical significance (black: two-sided U-tests; blue: FDR correction). The red dashed line marks the significance level $\alpha = 0.05$ and the scale of the y-axis is logarithmic. The green bars correspond to the *p*-values shown in the boxplots. **(b)** The correlation (left) between the *lesion*-tract density index (L-TDI) and the OS in patients known to be deceased was strong ($\rho = -0.209$, $p = 0.002$, two-sided exact test). Upon splitting these patients into two groups based on the 50$^{th}$ and 20$^{th}$/80$^{th}$ TDI percentiles (right), the median survival times in patients with smaller L-TDIs were significantly higher ($U = 7448.5$, $p = 0.0071$, $U = 1488.5$, $p = 0.0001$; two-sided U-tests). Differences in median survival times were checked across different stratification thresholds (bottom right), reaching near zero $p$-values for aggressive percentiles (black: two-sided U-tests; blue: FDR correction). The red dashed line marks the significance level $\alpha = 0.05$ and the scale of the y-axis is logarithmic. The green bars correspond to the *p*-values shown in the boxplots. **(c)** The correlation (top, left) between TDIs and OS converged to the negative value in **(a)**, without reaching statistical significance. The 95% confidence intervals (shaded gray region) and the statistical significance (black; '***' $p = 0.001$, '**' $p < 0.01$, '*' $p < 0.05$, 'n.s.' $p > 0.05$) were computed using bootstrapping and permutation procedures with $n = 2500$ resamples respectively. The corresponding $p$ values were further corrected for multiple comparisons (blue; FDR corrected). Within each *death cutoff*, patients were further subdivided according to the 50$^{th}$ TDI percentile (bottom, left). Median survival times were not different between the TDI groups (black: two-sided U-tests; blue: FDR corrected). Thin vertical black lines mark the 2$^{nd}$ and 3$^{rd}$ quartiles. In each death cutoff, the median TDI values did not significantly differ for dead and alive patients (right; black: two-sided U-tests; blue: FDR corrected). The y-axis is min-max scaled for visualization purposes, but the numerical analyses were done with the raw TDI values. **(d)** We repeated the same analyses described in **(c)** for the L-TDI marker. The correlation (top, left) between L-TDIs and OS settled to a negative and significant value, as seen in **(b)**. A clear boundary appeared around 18 months, with lower L-TDIs significantly associated with longer median survival times (bottom, right) and patients surviving at least 18 months exhibiting lower L-TDI values (right). All the $p$-values survived FDR corrections.

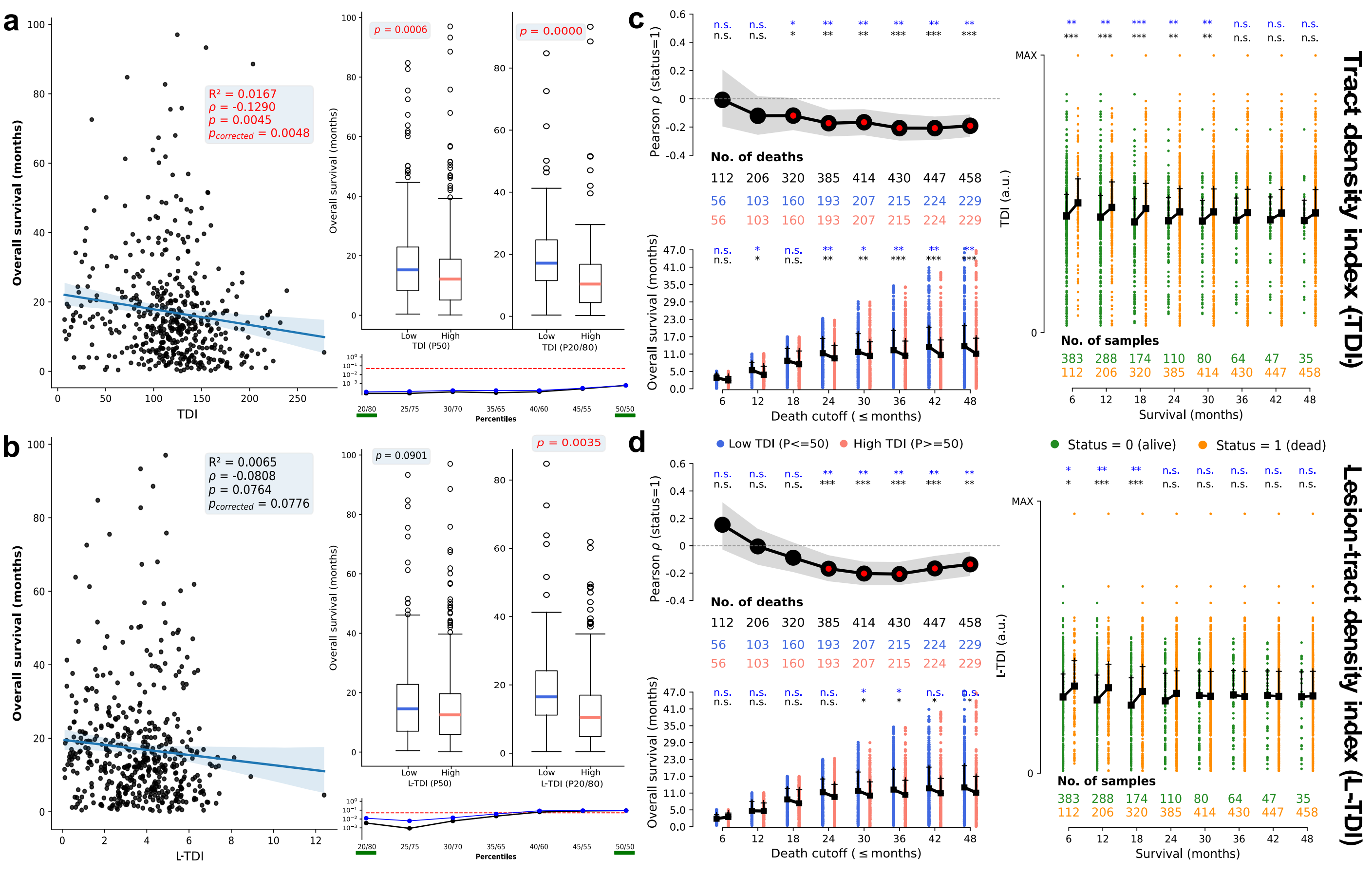

**Figure S5. Association between (lesion-)tract density indices and overall survival in the UPENN cohort.** **(a)** The correlation (left) between the tract density index (TDI) and the overall survival (OS) in patients known to be deceased was negative and significant ($\rho = -0.1290$, $p = 0.0045$, two-sided exact test). Upon splitting these patients into two groups based on the 50th and 20th/80th TDI percentiles (right), the median survival times in patients with lower TDIs were higher ($p = 0.0006$; $p < 0.0001$; two-sided U-tests). Boxplots depict the median for the low (blue) and high (salmon) TDI strata the interquartile range (black box), the 1st/4th quartiles (whiskers), and outliers. Differences in median survival times were checked across different stratification thresholds (bottom right), reaching statistical significance (black: two-sided U-tests; blue: FDR correction). The red dashed line marks the significance level $\alpha = 0.05$ and the scale of the y-axis is logarithmic. The green bars correspond to the *p*-values shown in the boxplots. **(b)** The correlation (left) between the *lesion*-tract density index (L-TDI) and the OS in patients known to be deceased was negative and almost reached statistical significance ($\rho = -0.0808$, $p = 0.0764$, two-sided exact test). Upon splitting these patients into two groups based on the 50th and 20th/80th TDI percentiles (right), the median survival times in patients with smaller L-TDIs were higher ($p = 0.0901$, $p = 0.0035$; two-sided U-tests). Differences in median survival times were checked across different stratification thresholds (bottom right), reaching statistical significance at the 40th/60th percentiles (black: two-sided U-tests; blue: FDR correction). The red dashed line marks the significance level $\alpha = 0.05$ and the scale of the y-axis is logarithmic. The green bars correspond to the *p*-values shown in the boxplots. **(c)** The correlation (top, left) between TDIs and OS converged to the negative value in **(a)**, reaching statistical significance. The 95% confidence intervals (shaded gray region) and the statistical significance (black; '***' $p = 0.001$, '**' $p < 0.01$, '*' $p < 0.05$, 'n.s.' $p > 0.05$) were computed using bootstrapping and permutation procedures with $n = 2500$ resamples respectively. The corresponding $p$ values were further corrected for multiple comparisons (blue; FDR corrected). Within each *death cutoff*, patients were further subdivided according to the 50th TDI percentile (bottom, left). Median survival times were different between the TDI groups beyond the 24 months threshold (black: two-sided U-tests; blue: FDR corrected). Thin vertical black lines mark the 2nd and 3rd quartiles. In each death cutoff, the median TDI values significantly differed for dead and alive patients (right; black: two-sided U-tests; blue: FDR corrected). The y-axis is min-max scaled for visualization purposes, but the numerical analyses were done with the raw TDI values. **(d)** We repeated the same analyses described in **(c)** for the L-TDI marker. The correlation (top, left) between L-TDIs and OS settled to a negative and significant value, as seen in **(b)**. A clear boundary appeared around 24 months, with lower L-TDIs significantly associated with longer median survival times (bottom, right) and patients surviving at least 18 months exhibiting lower L-TDI values (right). All the $p$-values survived FDR corrections.

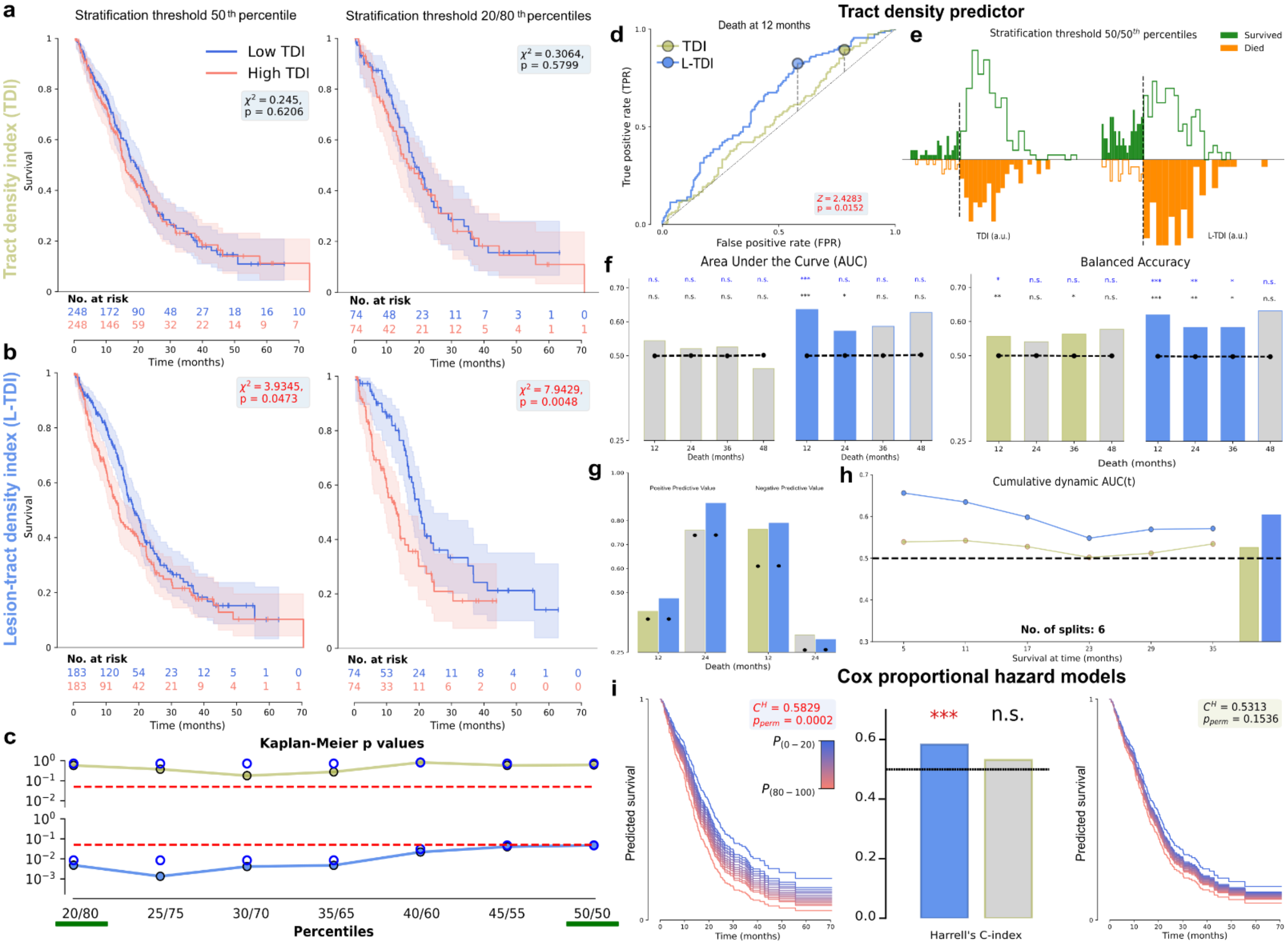

**Figure S6. (Lesion-)tract density indices as predictors of survival in the UCSF cohort. (a)** Kaplan-Meier estimators, including the 95% confidence intervals for the low (blue) and high (red) TDI groups. No differences in survival rates of the two groups were visible for the stratifications based on the 50th percentile (left; $\chi^2 = 0.245$, $p = 0.6206$, two-sided log-rank test), and the 20th/80th percentiles (right; $\chi^2 = 0.3064$, $p = 0.5799$, two-sided log-rank test). **(b)** Kaplan-Meier estimators, including the 95% confidence intervals for the low (blue) and high (red) L-TDI groups. Significant differences in the survival rates of the two groups were visible for the stratifications based on the 50th percentile (left; $\chi^2 = 3.9345$, $p = 0.0473$, two-sided log-rank test), and the 20th/80th percentiles (right; $\chi^2 = 7.9429$, $p = 0.0048$, two-sided log-rank test). **(c)** The $p$-values of the two-sided log-rank tests in **(a)** and **(b)** across different stratification thresholds for the TDI (top) and L-TDI (bottom) markers, respectively. Colored lines and black circles show the original $p$-values, while the blue circles depict the $p$-values after FDR correction. The red dotted line marks the significance level ($p = 0.05$). The y-axis is on a log scale to enhance visualization. The green bars correspond to the $p$-values shown in **(a-b)**. **(d)** Receiver operating curves (ROCs) for the two different tract density-based markers. L-TDI rather than TDI values stratified at the 50th percentile proved to be a better death/survival classifier at 12 months ($Z = 2.4283$, $p = 0.0152$, two-sided DeLong test). The dashed gray lines and colored circles show the decision threshold maximizing the Youden's J statistic for each classifier. **(e)** Distribution of true negative (green bars), false negative (green-contoured), false positives (orange-contoured), and true positives (orange) for the TDI (left) and L-TDI (right) markers. Dashed black lines show the decision thresholds maximizing Youden's J statistic in **(d)**. **(f)** Area under the curve (left) and balanced accuracy (right) of the death/survival classification based on the decision in **(d-e)**. Classification metrics derived from the TDI marker are shown in light green-filled (significant) and -contoured (non-significant) bars, while L-TDI-based metrics are shown in light blue-filled (significant) and -contoured (non-significant) bars. Gray-filled/color-contoured bars depict non-significant results w.r.t. the dashed black lines (two-sided permutation tests in black text, $n = 5000$ resamples; '***' $p = 0.001$, '**' $p < 0.01$, '*' $p < 0.05$, 'n.s.' $p > 0.05$). Blue text for FDR corrected results. **(g)** Positive (left) and negative (left) predictive values of the classification in **(e)**. Colors follow the same convention as in **(f)**. **(h)** Cumulative dynamic area under the curve for the TDI and L-TDI markers as a function of the survival time using the same strata as in **(d-g)**. The colored bars (right) show the temporal average of the scores. **(i)** Summary of the fitted Cox proportional models to the survival as a function of the L-TDI (left) and TDI (right) markers. Center bars show the corresponding hazard ratios and their significance (two-sided permutation tests in black text, $n = 5000$ resamples). The colorbar shows the fitted survival function for different tract density-based markers from the lowest (blue) to the highest (red) values. It is visible how the L-TDI is associated with a higher and significant hazard ratio for the UCSF cohort.

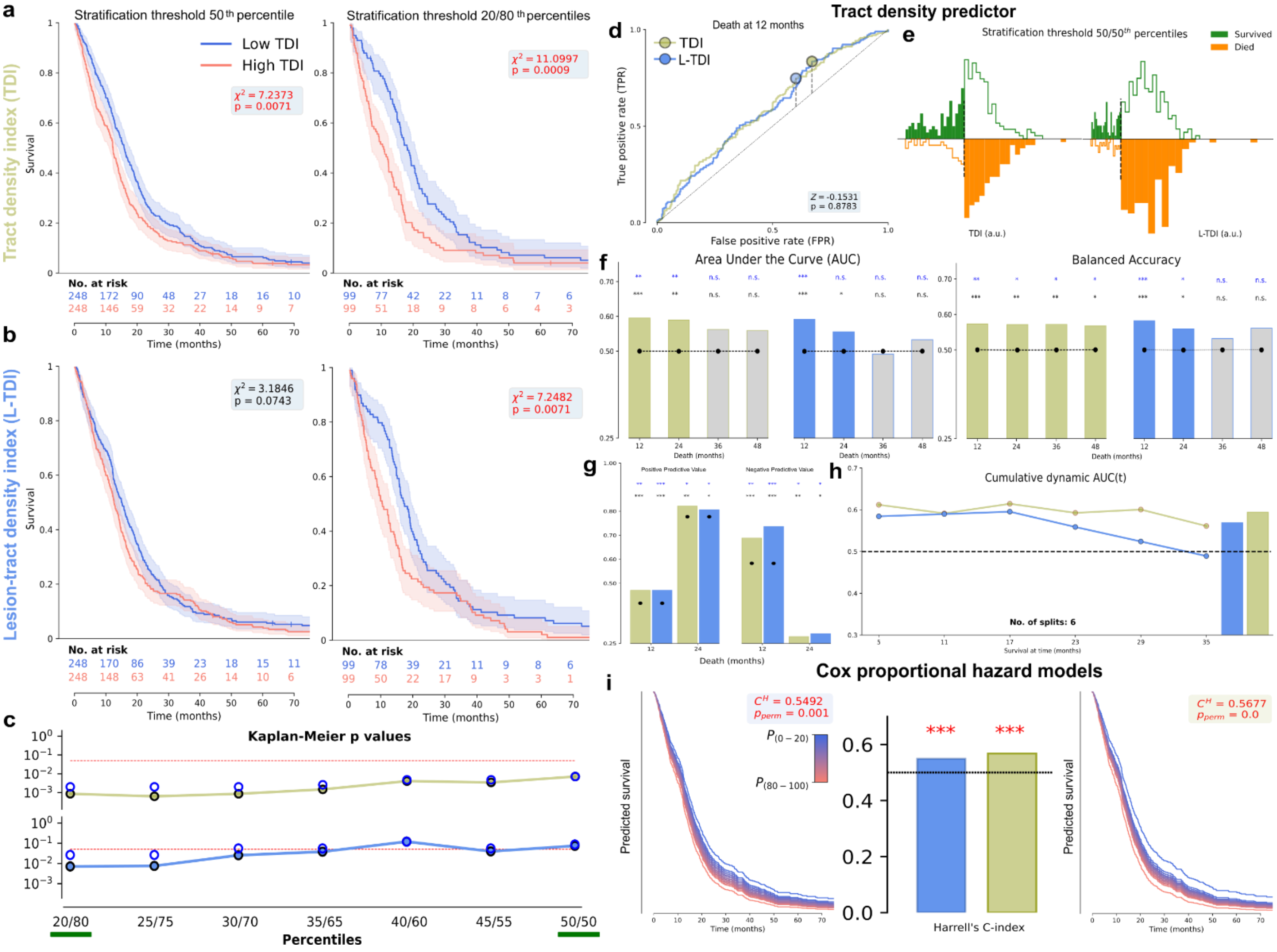

**Figure S7. (Lesion-)tract density indices as predictors of survival in the UPENN cohort. (a)** Kaplan-Meier estimators, including the 95% confidence intervals for the low (blue) and high (red) TDI groups. There were significant differences in survival rates of the two groups the stratifications based on the 50th percentile (left; $\chi^2 = 7.2373$, $p = 0.0071$, two-sided log-rank test), and the 20th/80th percentiles (right; $\chi^2 = 11.0997$, $p = 0.0009$, two-sided log-rank test). **(b)** Kaplan-Meier estimators, including the 95% confidence intervals for the low (blue) and high (red) L-TDI groups. Differences in the survival rates of the two groups were almost significant for the stratifications based on the 50th percentile (left; $\chi^2 = 3.1846$, $p = 0.0743$, two-sided log-rank test), and clearly visible for the 20th/80th percentiles (right; $\chi^2 = 7.2482$, $p = 0.0071$, two-sided log-rank test). **(c)** The $p$-values of the two-sided log-rank tests in **(a)** and **(b)** across different stratification thresholds for the TDI (top) and L-TDI (bottom) markers, respectively. Colored lines and black circles show the original $p$-values, while the blue circles depict the $p$-values after FDR correction. The red dotted line marks the significance level ($p = 0.05$). The y-axis is on a log scale to enhance visualization. The green bars correspond to the $p$-values shown in **(a-b)**. **(d)** Receiver operating curves (ROCs) for the two different tract density-based markers. L-TDI and TDI values stratified at the 50th percentile proved to be an equivalent death/survival classifier at 12 months ($Z = -0.1531$, $p = 0.8783$, two-sided DeLong test). The dashed gray lines and colored circles show the decision threshold maximizing the Youden's J statistic for each classifier. **(e)** Distribution of true negative (green bars), false negative (green-contoured), false positives (orange-contoured), and true positives (orange) for the TDI (left) and L-TDI (right) markers. Dashed black lines show the decision thresholds maximizing Youden's J statistic in **(d)**. **(f)** Area under the curve (left) and balanced accuracy (right) of the death/survival classification based on the decision in **(d-e)**. Classification metrics derived from the TDI marker are shown in light green-filled (significant) and -contoured (non-significant) bars, while L-TDI-based metrics are shown in light blue-filled (significant) and -contoured (non-significant) bars. Gray-filled/color-contoured bars depict non-significant results w.r.t. the dashed black lines (two-sided permutation tests in black text, $n = 5000$ resamples; '***' $p = 0.001$, '**' $p < 0.01$, '*' $p < 0.05$, 'n.s.' $p > 0.05$). Blue text for FDR corrected results. **(g)** Positive (left) and negative (left) predictive values of the classification in **(e)**. Colors follow the same convention as in **(f)**. **(h)** Cumulative dynamic area under the curve for the TDI and L-TDI markers as a function of the survival time using the same strata as in **(d-g)**. The colored bars (right) show the temporal average of the scores. **(i)** Summary of the fitted Cox proportional models to the survival as a function of the L-TDI (left) and TDI (right) markers. Center bars show the corresponding hazard ratios and their significance (two-sided permutation tests in black text, $n = 5000$ resamples). The colorbar shows the fitted survival function for different tract density-based markers from the lowest (blue) to the highest (red) values. It is visible how the L-TDI is associated with a higher and significant hazard ratio for the UCSF cohort.

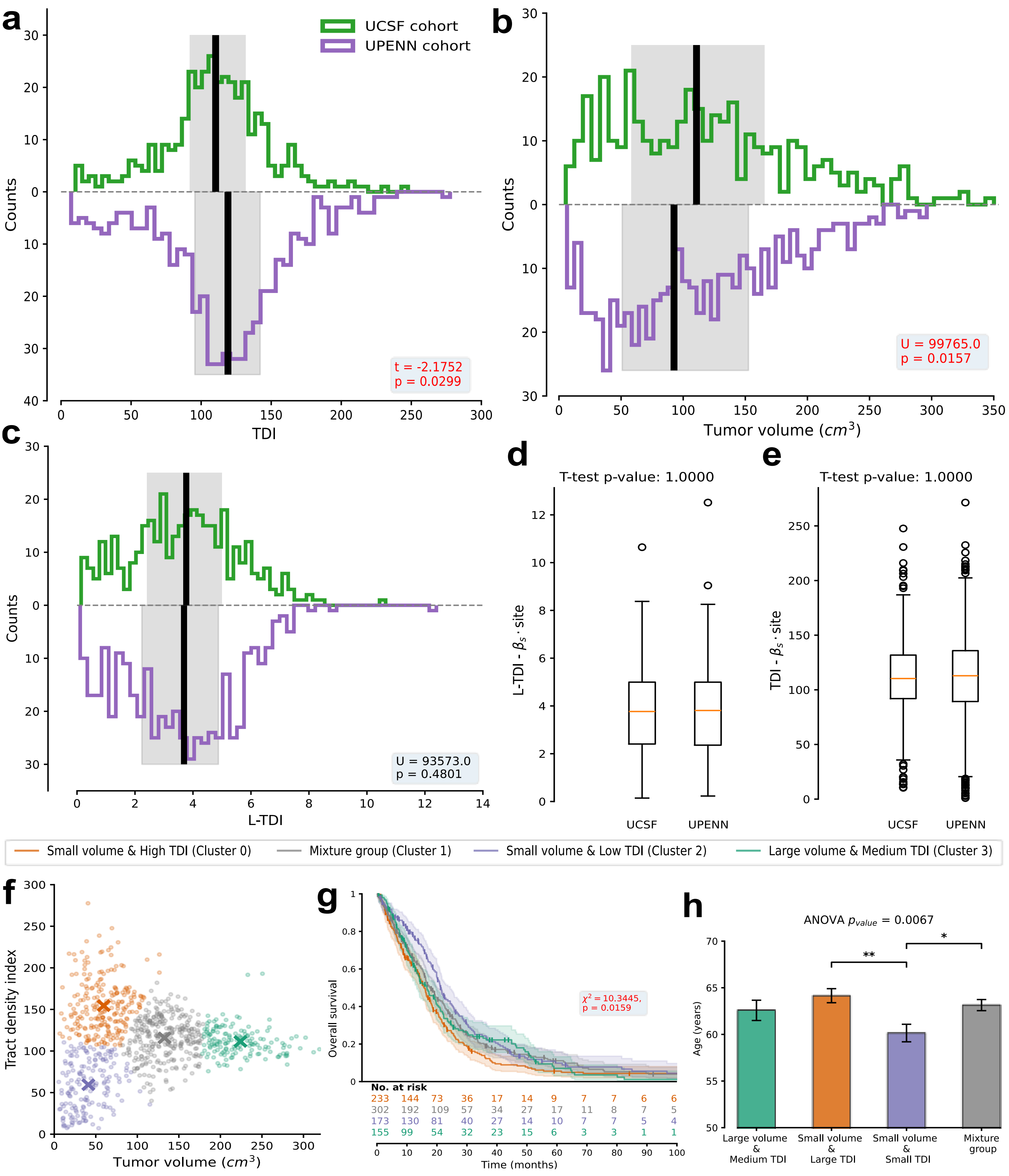

**Figure S8. Cohort differences in tract densities and tumor volumes. (a)** Distribution of the tract density index (TDI) in both cohorts. A small but significant difference in [mean ± SEM] existed (t=-2.1752, p=0.0299; two-sided t-test). **(b)** Distribution of tumor volumes in both cohorts. A small but significant difference in [mean ± SEM] existed (U=99765, p=0.0157; two-sided Mann-Whitney test). **(c)** Distribution of the lesion-tract density index (L-TDI) in both cohorts. No significant difference in [mean ± SEM] existed (U=93573, p=0.4801; two-sided Mann-Whitney test), highlighting the robustness of the L-TDI marker. **(d-e)** Boxplots comparing the distribution of residuals for the **(d)** L-TDI and **(e)** TDI values in each cohort. If necessary, the small site effects observed in (a-c) could be removed by simply computing the residuals of a linear fit. **(f)** Automatic K-means clustering from the tumor volumes and TDIs reveal 4 distinct groups. **(g)** Kaplan-Meier curves of each group in **(f)** revealed decreased hazard rates for small tumors with reduced infiltration (purple) w.r.t. small tumors with increased infiltration as well as large tumors ($\chi^2 = 10.3445$, $p = 0.0159$, two-sided log-rank test). Survival rates were adjusted for site effects (see Methods). **(h)** Average [mean ± SEM] age in years for each cluster in **(f)**. At least one cluster had a significantly different age ($p = 0.0067$; one-way ANOVA). Follow-up analyses revealed that small tumors with low TDIs occurred in significantly younger patients (post hoc Tukey's HSD).

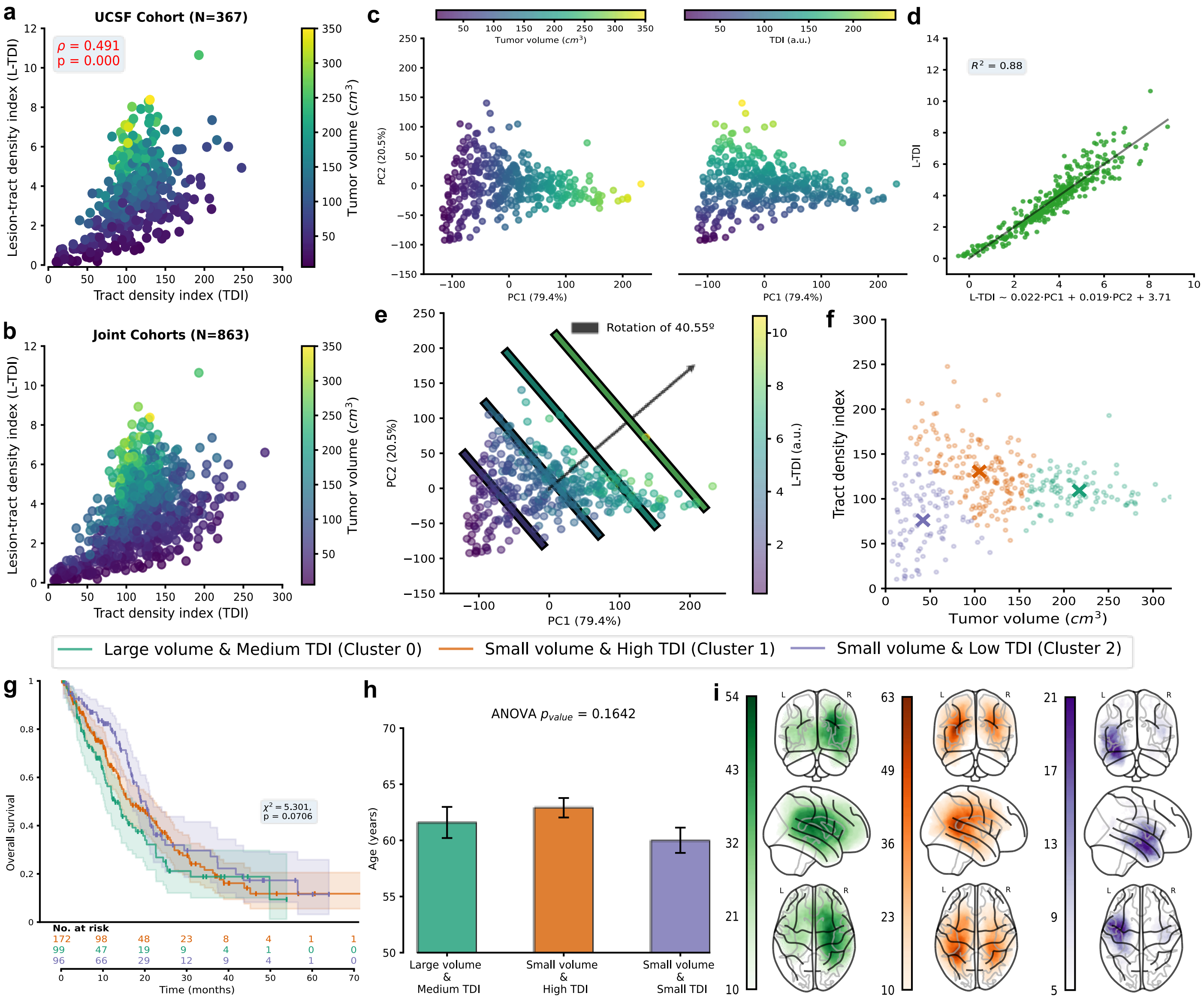

**Figure S9. Tract density markers, morphology, and anatomy of glioblastomas in the UCSF cohort. (a-b)** Scatter plots of the tract-density markers and the corresponding tumor volumes **(a)** UCSF and **(b)** joint cohorts. Tumor volume increased somewhat linearly with the L-TDI but not with the TDI. **(c)** Principal component (PC) analysis of the 3 features in (a-b). More than 99.9% of the explained variance can be attributed to the 1st and 2nd PCs. The 1st and 2nd PCs largely account for the variance in the tumor volumes (left) and TDIs (right). **(d)** Observed and predicted L-TDI values obtained from the linear combination of the first two PCs. The $R^2$ depicts the determination coefficient of the linear fit. **(e)** Scatter plot of the 1st and 2nd PCs together with the corresponding L-TDIs. The diagonal lines depict areas of constant L-TDI, and the black arrow indicates the direction along which the L-TDI increases. Because of the orthogonality of the first two PCs in **(d-e)**, the L-TDI contains unique and non-redundant information of both tumor volume and indirect white matter infiltration. **(f)** Automatic K-means clustering from the tumor volumes and TDIs reveal 3 distinct groups. **(g)** Kaplan-Meier curves of each group in **(f)** revealed decreased hazard rates for small tumors with reduced infiltration (purple) w.r.t. small tumors with increased infiltration, as well as large tumors ($\chi^2 = 6.1977$, $p = 0.0451$, two-sided log-rank test). **(h)** Average [mean ± SEM] age in years for each cluster in **(f)**. At least one cluster had a significantly different age ($F = 6.6746$, $p = 0.0013$; one-way ANOVA). Follow-up analyses revealed that small tumors with low TDIs occurred in significantly younger patients (post hoc Tukey's HSD). **(i)** Maximum intensity projection of the lesion group maps for the 3 distinct clusters in **(f)**. The colorbars show the number of subjects with a GBM in each point of the brain.

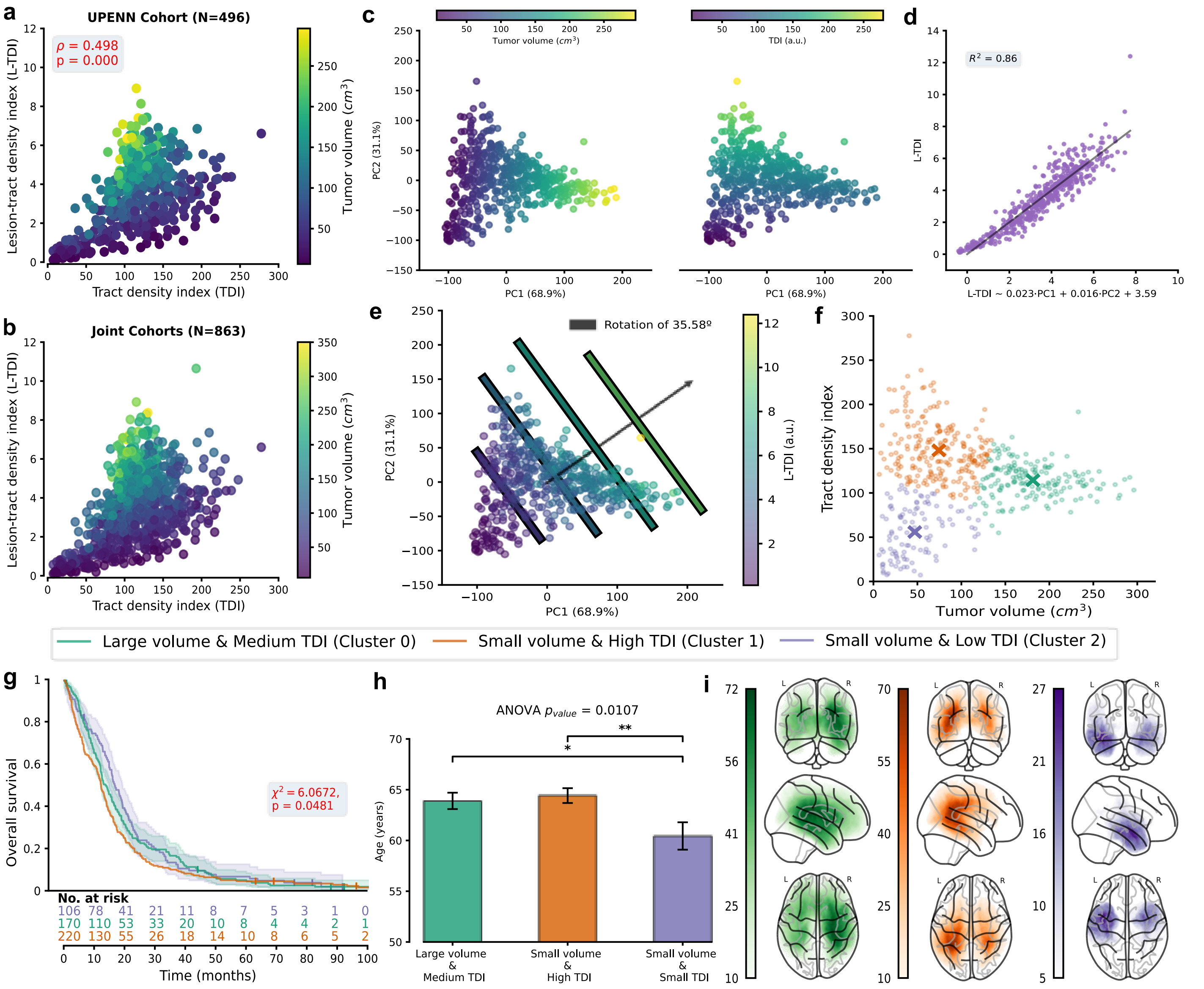

**Figure S10. Tract density markers, morphology, and anatomy of glioblastomas in the UPENN cohort. (a-b)** Scatter plots of the tract-density markers and the corresponding tumor volumes **(a)** UPENN and **(b)** joint cohorts. Tumor volume increased somewhat linearly with the L-TDI but not with the TDI. **(c)** Principal component (PC) analysis of the 3 features in (a-b). More than 99.9% of the explained variance can be attributed to the 1st and 2nd PCs. The 1st and 2nd PCs largely account for the variance in the tumor volumes (left) and TDIs (right). **(d)** Observed and predicted L-TDI values obtained from the linear combination of the first two PCs. The $R^2$ depicts the determination coefficient of the linear fit. **(e)** Scatter plot of the 1st and 2nd PCs together with the corresponding L-TDIs. The diagonal lines depict areas of constant L-TDI, and the black arrow indicates the direction along which the L-TDI increases. Because of the orthogonality of the first two PCs in **(d-e)**, the L-TDI contains unique and non-redundant information of both tumor volume and indirect white matter infiltration. **(f)** Automatic K-means clustering from the tumor volumes and TDIs reveal 3 distinct groups. **(g)** Kaplan-Meier curves of each group in **(f)** revealed decreased hazard rates for small tumors with reduced infiltration (purple) w.r.t. small tumors with increased infiltration, as well as large tumors ($\chi^2 = 6.1977$, $p = 0.0451$, two-sided log-rank test). **(h)** Average [mean ± SEM] age in years for each cluster in **(f)**. At least one cluster had a significantly different age ($F = 6.6746$, $p = 0.0013$; one-way ANOVA). Follow-up analyses revealed that small tumors with low TDIs occurred in significantly younger patients (post hoc Tukey's HSD). **(i)** Maximum intensity projection of the lesion group maps for the 3 distinct clusters in **(f)**. The colorbars show the number of subjects with a GBM in each point of the brain.

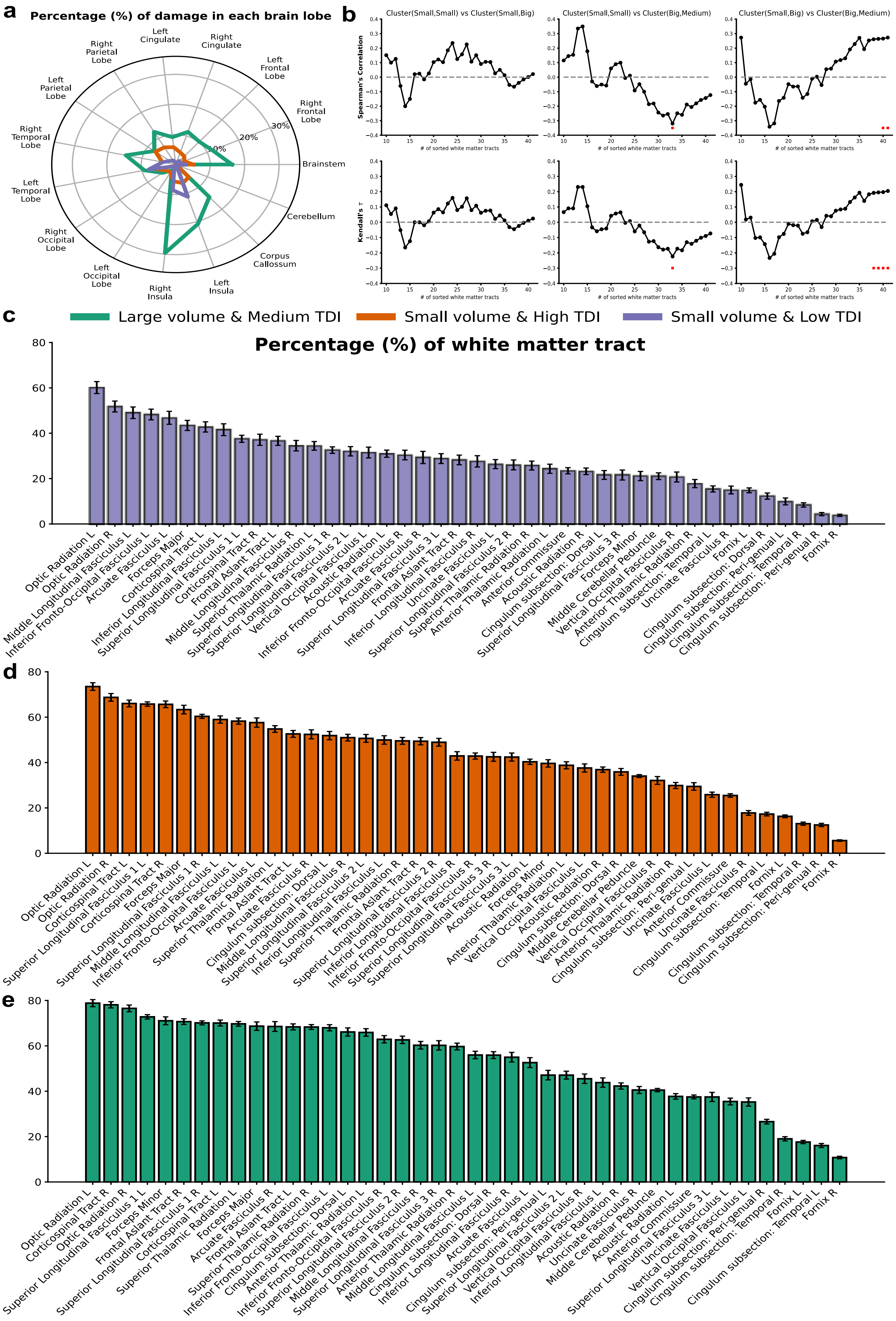

*Caption on the next page*

**Figure S11. Lesion and lesion-tract density maps overlap with brain lobes and white matter. (a)** Average overlap between the brain lobes defined in the USCLobes atlas and the tumor mask. The overlap was measured in terms of the number of voxels of a given brain lobe occupied by the tumor mask. **(b)** Spearman correlation **(top)** and Kendall's $\tau$ **(bottom)** coefficients between the sorted overlap sequences between patient-specific L-TDMs and the white matter tracts in the XTRACT atlas (see Main text; see also Fig. 3). The sequences were truncated in incremental steps from 10 to 42, which corresponded to the number of tracts defined in the XTRACT atlas. Significance was obtained through permutation tests in the case of Spearman's correlation with $N = 10000$ resamples and the exact test for Kendall's $\tau$ due to the absence of ties. Significance values ($p < 0.1$; two-sided) are marked with red squares. **(c-e)** Complete sorted sequence of the average overlap between the major white matter tracts defined in the XTRACT atlas and the patient-wise L-TDMs. The colors obey the legend above.

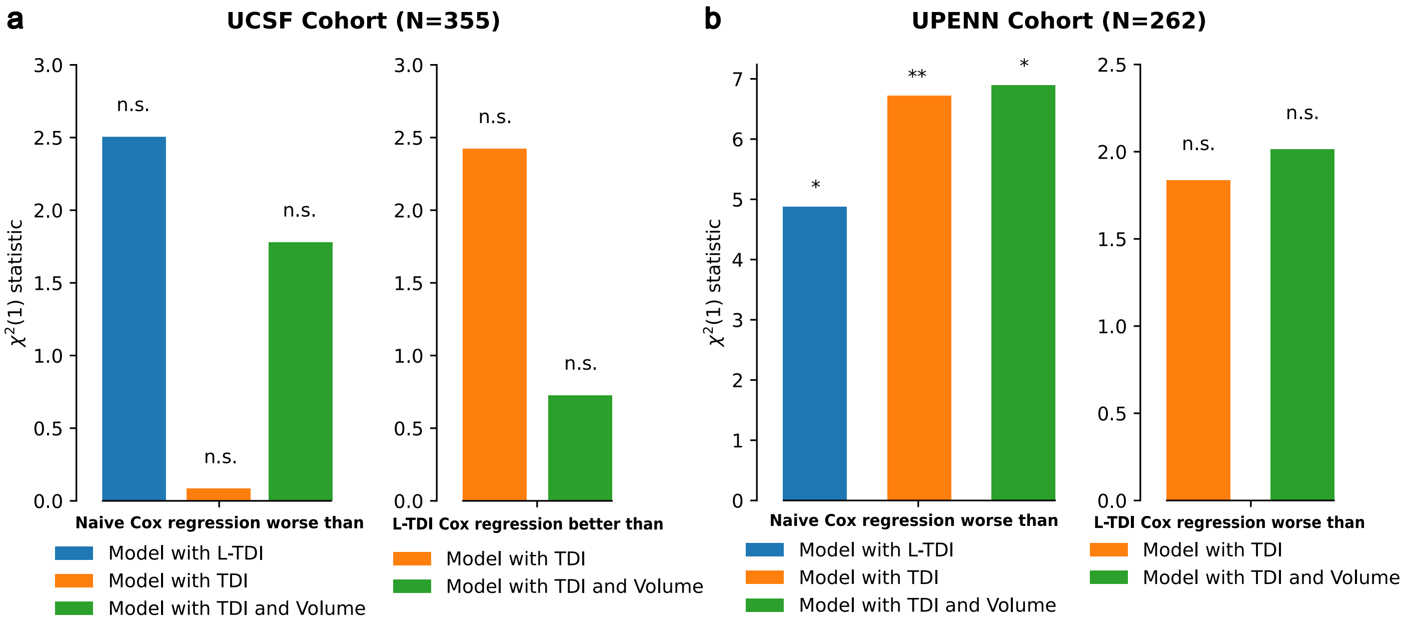

**Figure S12. Log-likelihood ratio tests in multivariate Cox models. (a-b, left)** One-sided log-likelihood ratio tests comparing the Cox models with added covariates w.r.t. the *naïve* in the **(e)** UCSF and **(b)** UPENN cohorts. **(a-b right)** *L-TDI* Cox model compared to the model with *TDI* and *TDI+volume* as added covariates of interest. All tests were done with 1 degree of freedom, except when comparing the *TDI+Volume* and *Naive* models ($df = 2$).

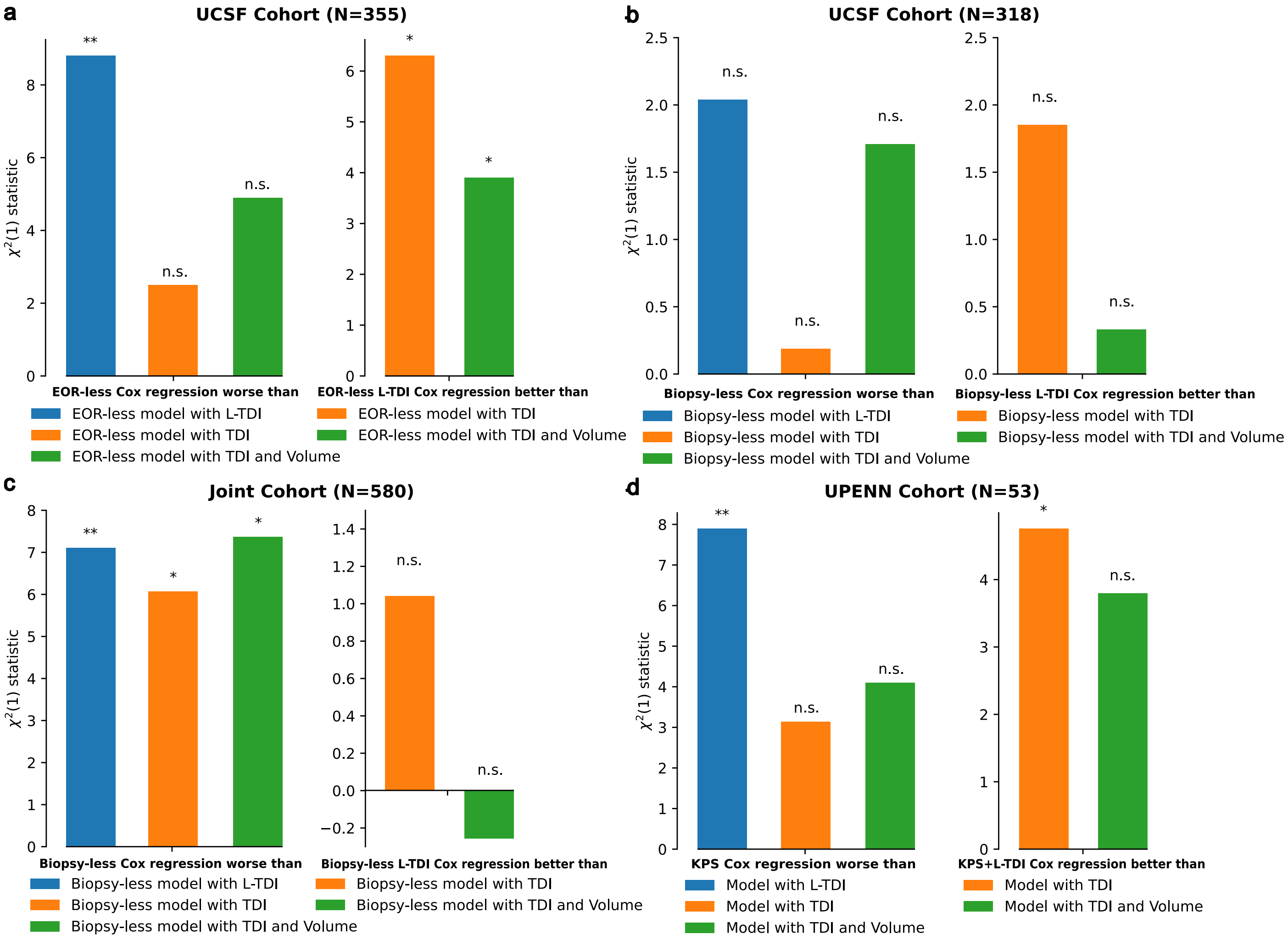

**Figure S13. Log-likelihood ratio tests in multivariate Cox models with different variations of covariates. (a)** Models were fitted without using the extent of resection (EOR) as a covariate of interest. One-sided log-likelihood ratio tests comparing the Cox models with added covariates w.r.t. the *naïve* (left). *L-TDI* Cox model compared to the model with *TDI* and *TDI+volume* as added covariates of interest. All tests were done with 1 degree of freedom, except when comparing the *TDI+Volume* and *Naive* models ($df = 2$; right). **(b)** Models fitted after discarding the patients that did not undergo a proper resection. One-sided log-likelihood ratio tests comparing the Cox models with added covariates w.r.t. the *naïve* (left). *L-TDI* Cox model compared to the model with *TDI* and *TDI+volume* as added covariates of interest. All tests were done with 1 degree of freedom, except when comparing the *TDI+Volume* and *Naive* models ($df = 2$; right). **(c)** Models fit after discarding the patients that did not undergo a proper resection, after correcting for site effects. One-sided log-likelihood ratio tests comparing the Cox models with added covariates w.r.t. the *naïve* (left). *L-TDI* Cox model compared to the model with *TDI* and *TDI+volume* as added covariates of interest. All tests were done with 1 degree of freedom, except when comparing the *TDI+Volume* and *Naive* models ($df = 2$; right). **(d)** Models fitted using only the patients with available Karnofsky performance status (KPS) score in the UPENN cohort and discarding samples with missing information. One-sided log-likelihood ratio tests comparing the Cox models with added covariates w.r.t. the *naïve* (left). *L-TDI* Cox model compared to the model with *TDI* and *TDI+volume* as added covariates of interest. All tests were done with 1 degree of freedom except when comparing the *TDI+Volume* and *Naive* models ($df = 2$; right).

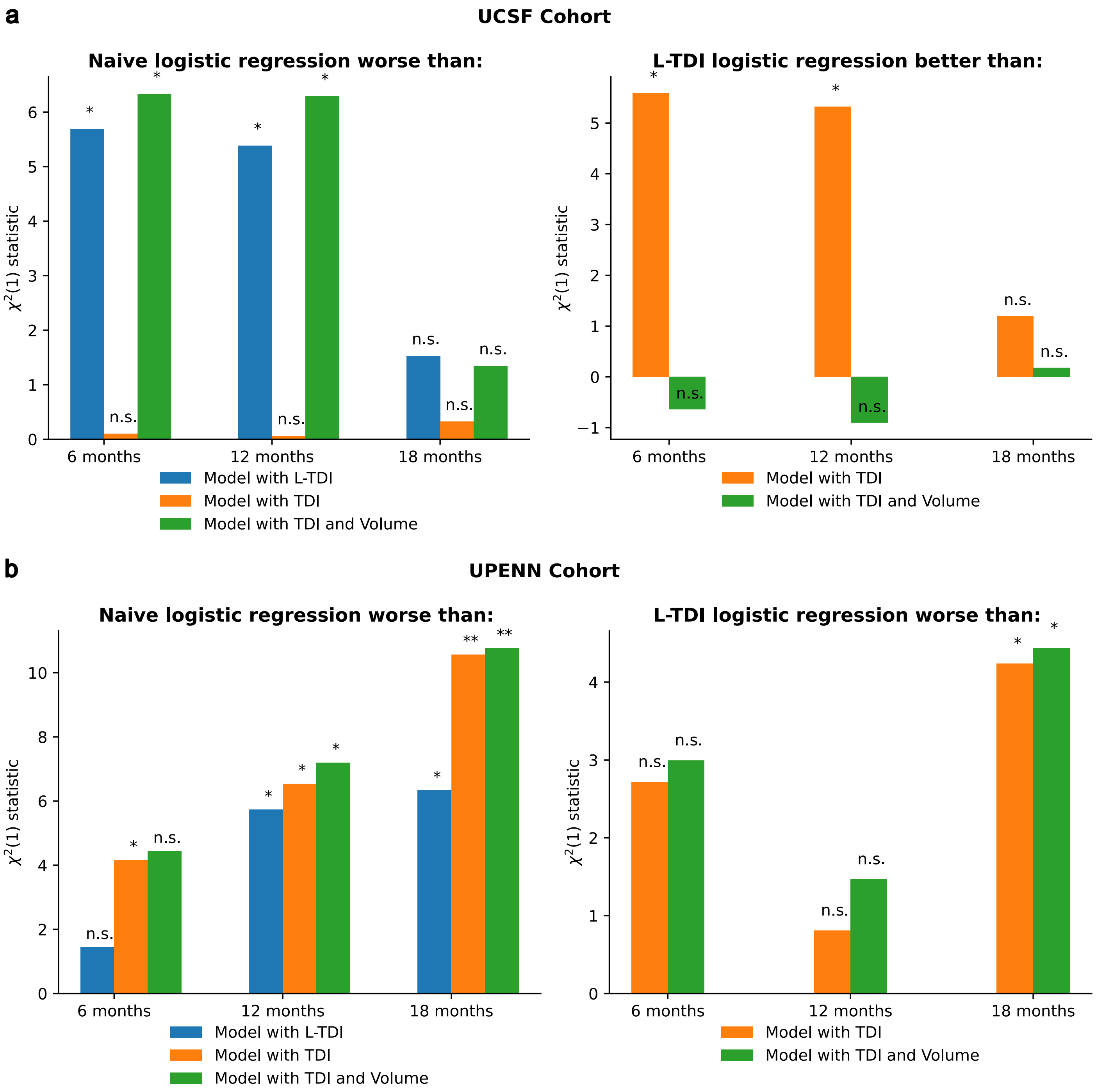

**Figure S14. Log-likelihood ratio tests in multivariate logistic regression.** The logistic regressions were fitted to predict the probability of dying (status=1) before 6, 12, and 18 months. Patients who were lost to follow-up (status=0) before each time horizon were discarded. **(a-b, left)** One-sided log-likelihood ratio tests comparing the logistic regressions with added covariates w.r.t. the *naive* in the **(e)** UCSF and **(b)** UPENN cohorts. **(a-b right)** *L-TDI* logistic regression compared to the regression with *TDI* and *TDI+volume* as added covariates of interest. All tests were done with 1 degree of freedom, except when comparing the *TDI+Volume* and *Naive* models ($df = 2$).
